# Depression and anxiety as causes and consequences of urinary incontinence in women: a population-based study

**DOI:** 10.64898/2026.03.16.26348501

**Authors:** Kimberley Burrows, Kate Tilling, Marcus J Drake, Rochelle Knight, Tom M Palmer, Carol Joinson

## Abstract

**Objective:** To examine the bidirectional relationships between depression, anxiety, neuroticism, and urinary incontinence in women.

**Design:** A prospective time-to-event and two-sample Mendelian randomisation (MR) study.

**Setting:** Individual participant data from the UK Biobank and summary genome-wide association (GWAS) study data from international consortia.

**Participants:** Up to 118 526 UK Biobank women with linked health records and up to 1.6 million participants with GWAS summary data.

**Main outcome measures:** Urinary incontinence (UI) and its subtypes (stress, urge, mixed), urinary urgency (irrespective of leakage), depression, anxiety, and neuroticism.

**Results:** We triangulated evidence to demonstrate bidirectional relationships between depression/anxiety and UI. In prospective analyses adjusted for confounders, depression was associated with a higher rate of new onset UI (any UI: Hazard Ratio (HR) 1.67; 95% Confidence Intervals (CI) 1.55 to 1.81) and its subtypes, with the strongest associations observed for mixed UI (HR 1.91; 95%CI 1.59 to 2.31). Similarly, anxiety and higher neuroticism scores were prospectively associated with UI and its subtypes. In the reverse direction, all UI subtypes were associated with a higher rate of new onset depression (e.g. any UI: HR 1.40; 95%CI 1.27 to 1.54) and anxiety (e.g. any UI: HR 1.28; 95%CI 1.17 to 1.39). Two-sample MR provided evidence for a causal effect of genetic liability to depression and neuroticism on UI and its subtypes (e.g. depression on any UI: OR_ivw_; 1.25 95%CI 1.16 to 1.35). Evidence for a causal effect in the reverse direction was weaker, with modest effects of genetic liability to any UI on depression. Little evidence was found for causal effects of anxiety with UI subtypes in either direction. Results were largely robust to sensitivity analyses.

**Conclusion:** We find evidence of bidirectional relationships between depression/anxiety and UI. Evidence that depression, anxiety and neuroticism are predictors of UI onset has implications for treatment. Research is needed to examine if treatments for depression/anxiety could be effective in alleviating UI.

**KEY MESSAGE:** *What is already known on this topic:*

- Urinary incontinence (UI) co-occurs with depression and anxiety, but the exact nature of the relationship is poorly understood because much of the existing evidence comes from cross-sectional studies.
- Amongst the existing prospective studies, only one used a clinically validated questionnaire to assess UI, few distinguished between UI subtypes (stress, urgency and mixed UI), and some did not adjust for important confounders.
- It is commonly believed that depression and anxiety are consequences of UI; if they are also causes of UI this has important implications for clinical care.

*What the study adds:*

- Our study demonstrates that the relationship between UI and depression/anxiety is bidirectional; We found that depression, anxiety and neuroticism (a personality trait characterised by a disposition to experience depression and anxiety) are predictors of UI onset and that UI is associated with new onset depression and anxiety.
- Depression and anxiety are not routinely assessed in urology clinics, and a continued failure to recognise their contribution to the onset and persistence of UI could be a cause of low success rates of existing treatments for UI.

## BACKGROUND

Urinary incontinence (UI) affects 25-45% of women worldwide, and the prevalence is rising [1]. The economic burden of UI on individuals’ quality of life and work productivity, health-systems, long-term care, and society is well-established. For example, a 2023 study estimated that the EU economic burden of UI was €80 billion including productivity losses and costs relating to health care, caregivers, and waste disposal [2]. UI can be differentiated through distinct but often co-occurring subtypes: stress UI (SUI: urine leakage during exertion, such as exercise, coughing or laughing); urgency UI (UUI: leakage associated with a strong and sudden need to urinate), and mixed UI (MUI: a combination of SUI and UUI) [3]. Genetic, biological, psychological, and environmental factors contribute to UI, but causes remain poorly understood. UI tends to be chronic and treatment-resistant, resulting in high NHS costs [4,5]. It is now recognised that low treatment success rates might arise from clinicians failing to recognise possible causal effects of mental health problems on UI [4,6]. A global multi-disciplinary think tank at the International Consultation on Incontinence Research Society meeting in 2025 concluded that research aimed at advancing understanding of the relationship between mental health problems and UI is crucial to improving clinical care [7].

Comorbidity between UI and mental health problems including depression and anxiety is well established in cross-sectional studies [8]. An unresolved issue, however, is whether depression and anxiety are causes or consequences of UI. It is often assumed that the elevated rates of mental health problems are due to the adverse consequences of UI on the daily lives of affected people [9]. An alternative explanation, that mental health problems are a cause of UI, is also highly plausible. For instance, studies have found that depression and neuroticism (a personality trait characterised by a disposition to experience negative affect) are prospectively associated with subsequent UI (any urinary leakage irrespective of subtype) in women [10–13].

Only a few studies have differentiated between UI subtypes. These include a 10-year longitudinal study reporting bidirectional relationships between UI subtypes (UUI and SUI) and mental health problems (depression and anxiety) [14,15]. Another study found that UUI (but not SUI) at baseline was associated with an increased incidence of anxiety and depression a year later and that incident UUI (but not SUI) was predicted by anxiety but not depression [16]. Previous studies did not use validated questionnaires to assess UI, and some did not adjust for important confounders including lifestyle and health/reproductive factors. More recently, a prospective cohort study, using validated questionnaires to assess UI (including UI subtypes), found that depression was associated with subsequent ‘any UI’, MUI and urinary urgency but only SUI was associated with subsequent depression [17].

Prospective studies provide insights into the direction of the relationship between UI and mental health problems but are limited by unmeasured and residual confounding and, hence, cannot provide conclusive evidence of causal effects. To try to overcome this, a recent study, based on a large UK cohort, examined the association between common genetic variants (polygenic risk scores – PRS) for psychiatric traits and UI [18]. Since genetic variants are randomized and fixed at conception, they should not be associated with environmental confounders at a population level. The study found evidence that PRSs for depression, anxiety, and neuroticism were associated with any UI, MUI and urinary urgency, but there was little evidence of associations with SUI [18]. It is not possible, however, to conclude from this study that psychiatric traits have causal effects on UI. The use of PRS to instrument an exposure is conceptually equivalent to Mendelian randomisation (MR); however, PRSs prioritise predictive accuracy by aggregating many variants, including those with pleiotropic effects, thus violating the exclusion restriction assumption required for causal inference [19,20]. Bidirectional MR applies the same principle to strengthen causal inference by using genetic variants as instrumental variables to examine if a specific trait is a cause or consequence of another trait, or whether there is a true bidirectional causal effect between two traits [21,22].

In the present study we used linked health records on UI in women from UK Biobank to examine relationships between UI (including UI subtypes) and depression, anxiety, and neuroticism. We performed a time-to-event analysis examining the relationship between (i) depression, anxiety and neuroticism at baseline and UI at follow-up and (ii) UI at baseline and depression and anxiety at follow up. We also performed a bidirectional two-sample MR analysis examining bidirectional causal relationships between genetic liability to depression, anxiety and neuroticism, and UI.

## METHODS

The results of this study are presented in accordance with the STROBE (The Strengthening the Reporting of Observational Studies in Epidemiology) and STROBE-MR guidelines. The STROBE [23] and STROBE-MR [24] checklists are available in supplementary materials.

### Study design and population

The UK Biobank (UKB) is an ongoing prospective cohort study of approximately 500 000 participants (55% women) aged 40 to 69 [25,26]. Participants were recruited across twenty-two assessment centres in England, Scotland, and Wales between 2006 and 2010 with a response rate of 5.5% [25]. The study has collected sociodemographic information, medical history, psychosocial and early life exposure history, physical measures at assessment centres, and genomic/omic samples. Medical outcomes have been collected prospectively via data linkage to hospital admission records, death and cancer registries, and primary care records. Our study sample included female participants with data from baseline assessment, linked primary care records, and linked hospital records (see supplementary figure 1). Prior to analyses participants who had withdrawn consent were removed and the sample was restricted to females according to sex (Data-Field 31; 54%; n=273 158; see figure 1 for the study flow chart).

**Figure 1.**
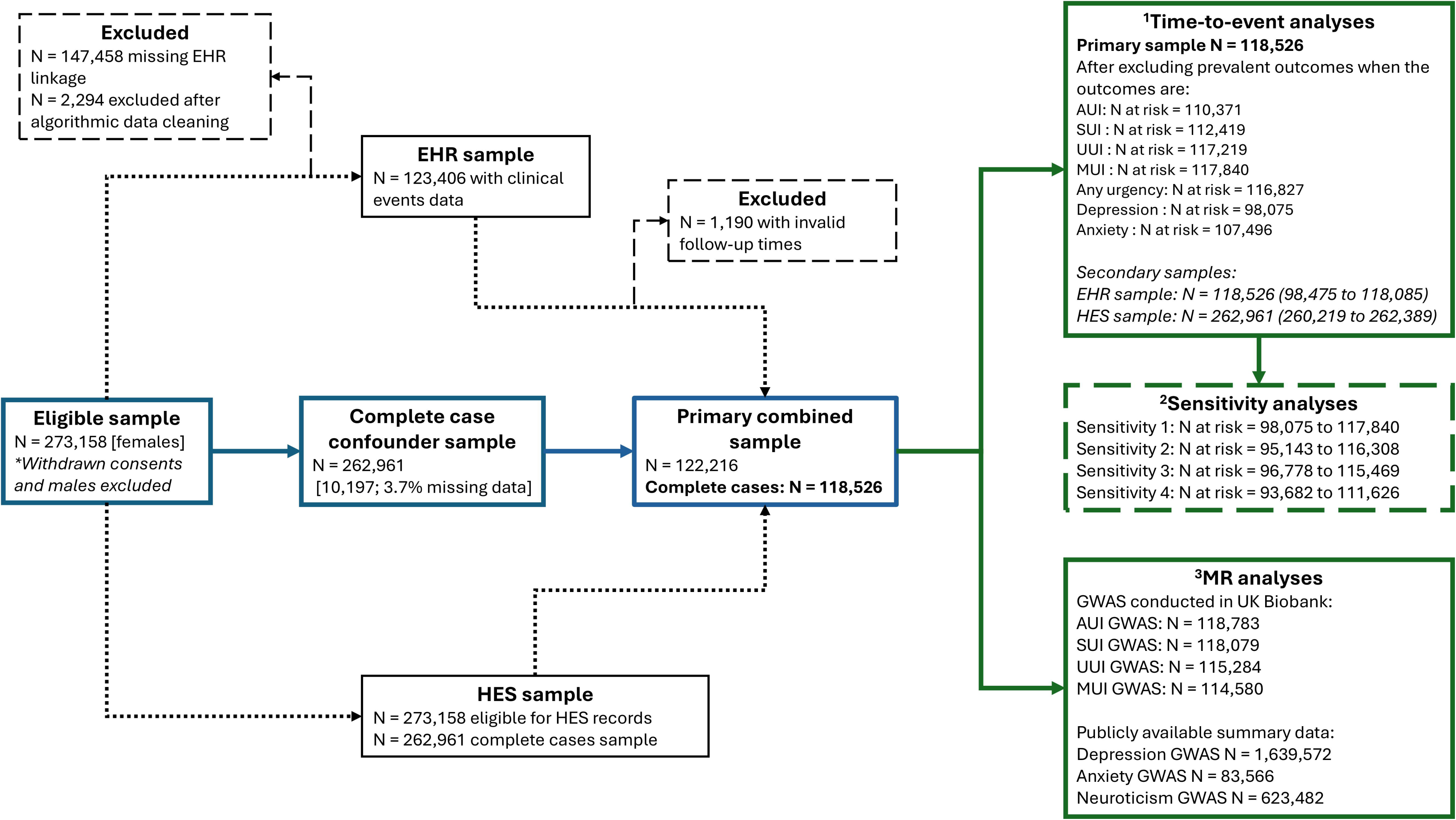
Flow chart of participant numbers. EHR Electronic Health Records; HES Hospital Episode Records; AUI any Urinary Incontinence (UI); SUI stress UI; UUI urgency UI; MUI mixed UI; GWAS genome-wide association study. 1; 2; 3;

### Linked electronic health records

Linked primary care data (hereafter referred to as electronic health record (EHR) data) were available from General Practitioner (GP) practices in England, Scotland and Wales that used either the Vision or TPP management systems as part of routine patient care. EHR data were available for approximately 45% of the cohort and included GP practice registration records along with coded diagnoses, test results, and prescriptions all recorded prior to 2016/2017. As the UKB purposefully carried out minimal data cleaning, we used an adapted set of standards described by Darke *et al* [27]. This algorithm was used to 1) clean the raw data, 2) identify continuous periods of data collection, and 3) establish censoring dates, resulting in accurate follow-up times for participants with fragmented registration records (see Darke *et al.* 2021 and supplementary methods for further details).

In addition to EHR data, all participants were linked to hospital inpatient records (hereafter referred to as Hospital Episodes Statistics (HES) data) with data on primary and secondary diagnoses available across different dates for England, Scotland and Wales with a minimum coverage of eight years prior to baseline assessment (see supplementary methods for further details).

### Defining urinary incontinence

In consultation with clinicians, we developed a comprehensive diagnosis code list for UI in women, comprising Read v2 and Clinical Terms V3 (CTV3) codes (hereafter referred to as Read codes) for EHR data and ICD-10 codes for HES data (supplementary table 1). These codes were used to identify cases of any UI (AUI) and its three subtypes: stress UI (SUI), urgency UI (UUI), and mixed UI (MUI). We also identified diagnoses of any urinary urgency with or without UI (hereafter referred to as any urgency) because urinary urgency has been linked to depression and anxiety irrespective of urinary leakage [9].

Using EHR data, we classified participants as cases if they had subtype-specific Read codes present, and as non-cases if those specific codes were absent (see supplementary table 1 for all Read codes). For MUI, cases required both SUI and UUI codes. For AUI, cases included participants with SUI and/or UUI codes, or codes for non-specific UI (e.g. 1A23. Incontinence of urine). Non-cases for AUI included participants who were non-cases for SUI and UUI and did not have any codes for non-specific UI (see supplementary table 1 for case definition).

Using HES data, we identified cases through primary or secondary ICD-10 diagnoses: N39.3 for SUI and N39.4 for UUI. MUI cases required both SUI and UUI diagnoses. AUI cases included any participant with SUI and/or UUI diagnoses. Non-cases for AUI included participants who were non-cases for SUI and UUI.

To maximise case ascertainment, we combined cases identified in EHR data with those in HES data. For each subtype, participants were classified as cases if they were cases in either data source, or as non-cases only if they were classified as non-cases in both data sources. As EHR data covers approximately 45% of UKB participants, this integration strategy reduced the available non-case sample size (compared to the HES data) but enhanced case identification. This combined EHR and HES sample was used for our primary analyses. However, we additionally report results for each of the EHR and HES samples separately in the supplementary materials. All diagnosis codes and variable definitions are given in supplementary table 1. A matrix of case definitions for SUI, UUI, MUI and AUI is shown in supplementary table 2.

### Defining depression, anxiety and neuroticism

#### Depression

For EHR data we defined cases of any depression as those with a Read Code for depression as previously defined in Fabbri *et al.* [28] (see supplementary table 3). We identified cases of depression in HES data as those with an ICD-10 primary or secondary diagnosis of F32, F33, F34, F38, and/or F39 (supplementary table 4). Non-cases were defined as absence of any depression diagnosis in their respective sample.

We also combined EHR and HES data to enhance case ascertainment. Cases were defined as participants who were cases for either the EHR or HES sample. The non-cases were defined as participants who were a non-case for both EHR and HES samples, thus restricting the sample to the 45% of participants with EHR data.

#### Anxiety

Anxiety cases were defined using a previously published list of Read codes from Fabbri *et al*. [28] for EHR data (see supplementary table 5) and the ICD-10 diagnosis codes F40 and F41 for HES data (see supplementary table 4). For the EHR sample, non-cases were defined as participants who did not have any diagnosis of anxiety Read codes, and for the HES sample, absence of any of the ICD-10 diagnosis codes. In defining anxiety cases, we took the same approach we used to maximise case ascertainment for UI and depression, by combining cases in either data sample or combining non-cases; restricting to those who are non-case for both data samples.

#### Neuroticism

We used a neuroticism score (Field-code 20127; derived variable from Smith *et al.* [29]) based on twelve dichotomous variables from the Eysenck Personality Questionnaire Revised Short Form (EPQ-RS) [30], collected using a touchscreen-based questionnaire at the UKBB assessment centres. This scale has been validated in middle-aged people against two of the most widely used measures of neuroticism (International Personality Item Pool (IPIP) and the NEO-Five Factor Inventory (NEO-FFI)) [31,32]. Each of the twelve dichotomous variables were summed to produce a neuroticism score ranging from 0 to 12, where an increasing score represents a higher level of neuroticism.

### Confounders and additional variables

We considered potential confounders of the bidirectional relationships between UI and mental health problems (depression and anxiety) and neuroticism, grouped into two categories: i) sociodemographic factors including age at recruitment, self-reported ethnicity (White and non-white), Townsend Deprivation Index (TDI), education level (degree vs. no degree), and cohabiting with spouse or partner (cohabiting with partner/spouse vs. other); and ii) lifestyle and health/reproductive factors including BMI (kg/m2; weight (kg) and height (m) measured at assessment centre), smoking status (never, previous, current), alcohol intake (never, infrequent, frequent), multimorbidity (fewer than 2 vs. 2 or more; taken from 33 self-reported non-cancer illnesses), menopause status (no, yes, not sure – had hysterectomy, not sure – other reason), hysterectomy (no vs. yes), hormone replacement therapy (HRT) use (ever; yes vs. no), and parity (nulliparous, one, 2 or more). All confounders were measured at baseline through assessment centre visits or self-completed questionnaires. Further information on the field IDs, coding, and recoding for each of the confounders is given in supplementary table 6.

### Statistical analyses – prospective analyses

We examined bidirectional prospective associations between UI (and its subtypes: SUI, UUI, and MUI, as well as any urgency) and depression/anxiety using multivariable Cox proportional hazards regression (see supplementary figure 2 for a directed acyclic graph). For each UI phenotype, we conducted two separate analyses: first with UI as the exposure and depression and anxiety (separately) as the outcomes, then with depression/anxiety as the exposures and UI as the outcome. Analyses examining any urgency were restricted to participants in the EHR sample, as urgency diagnoses are not available in hospital records. For neuroticism, we examined associations only in the direction of neuroticism to UI because neuroticism was measured at baseline only. We considered the combined EHR/HES sample as our primary analysis sample. However, we additionally report results for each of the EHR and HES samples separately in the supplementary materials.

Follow-up time was calculated from baseline assessment until the earliest of first occurrence of the outcome (UI or depression/anxiety depending on the direction of analysis), death, or end-of-study censoring. For the combined EHR/HES sample, the end-of-study censoring was determined as the earlier of the primary care data extraction date or date of general practice deregistration (see supplementary methods for details). For participants in the HES sample, end-of-study censoring occurred on 31 October 2022.

We fitted a series of models with progressive confounder adjustment: Model 1 was unadjusted, Model 2 adjusted for sociodemographic factors, and Model 3 additionally adjusted for lifestyle and health/reproductive factors. We considered Model 3 (fully adjusted) as our primary analytical model. For all models, we specified assessment centre as a cluster variable to obtain robust standard errors accounting for potential within-centre correlation.

We assessed the proportional hazards assumption for each exposure using Schoenfeld residuals, evaluated through visual inspection of scaled residual plots and formal statistical tests. All effect estimates are reported as hazard ratios (HR) with 95% confidence intervals (95% CI).

#### Sensitivity analyses

We conducted several sensitivity analyses to evaluate the robustness of our primary analyses. First, we observed violations of the proportional hazards assumption for several confounders in Model 3. We therefore fitted Cox models stratified by age group (<52, 53-60, ≥60 years), BMI category (underweight, normal weight, overweight, obese), menopausal status, and parity, as these were most commonly violated. Stratification allows baseline hazards to vary across strata while estimating a common exposure effect. Second, we repeated Model 3 excluding any participants who experienced the outcome within the first two years of follow-up to address potential reverse causation. Third, we repeated Model 3 applying further exclusions for any participants who are also cases for other psychiatric disorders (see supplementary methods for details). Fourth, we repeated Model 3 for the SUI and UUI phenotypes mutually excluding for the other, i.e for the SUI phenotype, any cases of UUI were excluded from the cases and non-cases, and vice versa (see supplementary methods for details). Finally, we fitted an additional model (Model 4), that adjusted as per Model 3 but mutually adjusted for depression and anxiety.

#### Missing data

For Cox proportional hazards regression, a complete case analysis produces unbiased estimates when missingness is independent of the outcome conditional on the covariates in the analysis model [33]. To assess potential bias in a complete case analysis owing to missingness in the linkage to primary care data (55% are missing), we tested whether the missing data mechanism was associated with the UI subtypes, depression, and anxiety, as ascertained from the full HES-sample, as these are outcomes in our bidirectional analyses. We found little evidence for any association (see supplementary methods for further details) and since the confounders were measured at baseline, the proportion of missing data was relatively low (3.73% in the HES sample and 3.02% in the EHR sample, see table 1). We therefore present results for a complete case analysis only.

**Table 1.** Characteristics of UK Biobank Women.

| Characteristic [N (%); mean (SD)] | HES | HES (complete cases) | EHR | EHR (complete cases) |
| --- | --- | --- | --- | --- |
|  | N = 273,158 | N = 262,961 | N = 122,216 | N = 118,526 |
| **Age at recruitment (years)** | 56.35 (8.00) | 56.35 (7.99) | 56.37 (7.96) | 56.37 (7.95) |
| **Ethnicity (ref. White)** |  |  |  |  |
| Non-white | 14,652 (5.4%) | 13,218 (5.0%) | 5,521 (4.5%) | 4,951 (4.2%) |
| Missing | 1,265 | NA | 484 | NA |
| **Townsend deprivation index** | -1.33 (3.04) | -1.37 (3.02) | -1.37 (2.98) | -1.41 (2.95) |
| Missing | 328 | NA | 167 | NA |
| **Cohabiting (ref. Partner)** |  |  |  |  |
| Other | 84,710 (31.0%) | 80,391 (30.6%) | 37,661 (30.8%) | 36,070 (30.4%) |
| **Education (ref. Other)** |  |  |  |  |
| Degree | 84,439 (31.5%) | 83,088 (31.6%) | 37,486 (31.1%) | 36,909 (31.1%) |
| Missing | 5,337 | NA | 1,511 | NA |
| **Alcohol consumption (ref. Never)** |  |  |  |  |
| Infrequent | 76,597 (28.1%) | 73,579 (28.0%) | 34,309 (28.1%) | 33,216 (28.0%) |
| Frequent | 169,801 (62.3%) | 165,131 (62.8%) | 75,857 (62.2%) | 74,283 (62.7%) |
| Missing | 739 | NA | 276 | NA |
| **Smoking status (ref. Never)** |  |  |  |  |
| Previous | 85,374 (31.4%) | 83,057 (31.6%) | 37,987 (31.2%) | 37,215 (31.4%) |
| Current | 24,347 (9.0%) | 23,320 (8.9%) | 10,923 (9.0%) | 10,562 (8.9%) |
| Missing | 1,513 | NA | 629 | NA |
| **Multimorbidity (ref. No)** |  |  |  |  |
| ≥ 2 | 38,971 (14.3%) | 37,166 (14.1%) | 17,317 (14.2%) | 16,601 (14.0%) |
| **BMI (kg/m<sup>2</sup>)** | 27.09 (5.20) | 27.07 (5.18) | 27.20 (5.22) | 27.18 (5.21) |
| Missing | 1,459 | NA | 639 | NA |
| **Menopause status (ref. No)** |  |  |  |  |
| Yes | 165,301 (60.7%) | 159,778 (60.8%) | 74,018 (60.8%) | 71,994 (60.7%) |
|  | **N = 273,158** | **N = 262,961** | **N = 122,216** | **N = 118,526** |
| Not sure - had a hysterectomy | 31,155 (11.4%) | 29,984 (11.4%) | 14,163 (11.6%) | 13,753 (11.6%) |
| Not sure - other reason | 11,720 (4.3%) | 11,212 (4.3%) | 5,259 (4.3%) | 5,055 (4.3%) |
| Missing | 1,010 | NA | 408 | NA |
| **HRT (ref. No)** |  |  |  |  |
| Yes | 103,865 (38.2%) | 100,417 (38.2%) | 46,651 (38.4%) | 45,451 (38.3%) |
| Missing | 1,574 | NA | 672 | NA |
| **Hysterectomy (ref. No)** |  |  |  |  |
| Yes | 51,058 (18.8%) | 49,030 (18.6%) | 23,113 (19.0%) | 22,383 (18.9%) |
| Missing | 1,006 | NA | 416 | NA |
| **Parity (ref. None)** |  |  |  |  |
| One | 36,438 (13.4%) | 35,136 (13.4%) | 16,718 (13.7%) | 16,231 (13.7%) |
| ≥ 2 | 184,856 (67.9%) | 178,459 (67.9%) | 82,694 (67.8%) | 80,403 (67.8%) |
| Missing | 828 | NA | 315 | NA |
| **Neuroticism score** | 4.56 (3.25) | 4.56 (3.25) | 4.63 (3.27) | 4.62 (3.27) |
| Missing | 57,743 | 53,712 | 25,460 | 23,820 |

*EHR Electronic Health Records (primary care records); HES Hospital Episode Statistics (hospital inpatient records); N number; ref. reference; BMI body mass index; HRT hormone replacement therapy; NA not applicable. Complete cases denote the samples (HES and EHR) that have complete cases for each of the confounders: age, ethnicity, TDI, education, and cohabiting status, alcohol, smoking, BMI, menopause, HRT, hysterectomy, and parity.*

### Two-sample bidirectional MR

We conducted two-sample bidirectional MR to examine evidence of bidirectional causal effects between UI and depression, anxiety, and neuroticism (supplementary figure 2). In MR genetic variants are used as instrumental variables for an exposure and can improve causal inference under the following assumptions: the genetic instrument(s) are (i) robustly associated with the exposure, known as the relevance assumption; (ii) are not associated with confounders of the exposure-outcome association, known as the independence assumption; and (iii) are only associated with the outcome through the exposure, known as the exclusion-restriction assumption [34,35].

#### Genetic instrument selection of depression, anxiety, and neuroticism

We selected genetic instruments for each mental health exposure using summary statistics from recently published GWAS meta-analyses of depression (n=1 639 572 after excluding UKB and 23andMe cohorts) [36], anxiety (n=83 566) [37], and neuroticism (n=623 482) [38]. All three GWAS included male and female participants of European ancestry. Detailed information on each GWAS is provided in supplementary table 7. We selected SNPs reaching genome-wide significance (P<5×10^−8^) and clumped these to ensure independence at linkage disequilibrium (LD) r2 < 0.001 within a 10 000 kb window resulting in 147 SNPs for depression, 5 SNPs for anxiety, and 132 SNPs for neuroticism (see supplementary table 8).

#### GWAS and instrumental variable selection of UI subtypes

We performed GWASs of AUI, SUI, UUI, and MUI in the combined EHR/HES sample using BOLT-LMM [39] with an additive genetic model adjusting for age, genotyping array, and genetic correlation matrix, as described previously [40,41]. As BOLT-LMM uses a linear mixed model approximation for binary traits, we converted the effect estimates and their SEs to the log odds scale (see supplementary methods). After applying genome-wide significance thresholds (P<5×10⁻□) and clumping parameters as described above, we identified fewer than 5 independent SNPs for most UI GWASs. Therefore, we relaxed the significance threshold for instrument selection to a P value of (P<5×10^−6^), resulting in 37 SNPs for AUI, 39 SNPs for SUI, 10 SNPs for UUI, and 19 SNPs for MUI after clumping (see supplementary table 8).

#### Statistical analysis – two-sample bidirectional MR

We assessed evidence for weak instruments using the two-sample version of the mean F-statistic 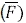 where a F-statistic <10 suggests evidence of weak instruments [42]. Exposure and outcome SNP effects were harmonised so that the effect allele was the same in both. If a SNP from the instrument was unavailable in the outcome, an attempt to find proxies was made with a minimum LD *r*^2^ = 0.8 and palindromic SNPs were aligned with Minor Allele Frequency (MAF) < 0.3 (supplementary table 8).

Two-sample MR was conducted using the *TwoSampleMR* R package (version 0.6.22 [43]). We used an inverse variance weighted (IVW) method [44] to meta-analyse the SNP specific Wald estimates (SNP-outcome estimate divided by SNP-exposure estimate) using fixed effects, to obtain an estimate for the causal effect of the exposure on the outcome. This was our primary analysis method. For exposure GWAS with fewer than five SNPs, we estimated the causal effect by calculating Wald ratios for each SNP. We also conducted analyses with SNPs reaching a less-stringent SNP-exposure association P value threshold of 5×10^−6^ [45] using a debiased IVW (db-IVW) method [46]. The debiased IVW method is robust to weak instruments because it applies a bias correction factor and relaxes the NO Measurement Error (NOME) assumption by estimating variances in the model.

Causal estimates were converted to odds ratios for binary outcomes which represents the odds of the outcome per unit increase in log odds ratio of a binary exposure. To aid in interpretation, log odds and SEs were rescaled (by multiplying by 0.693, i.e. log(2)) before exponentiating to represent the OR per doubling in odds of liability to binary exposures [47].

#### Sensitivity analyses

The IVW method will return an unbiased estimate in the absence of horizontal pleiotropy or where horizontal pleiotropy is balanced [48]. To account for potential directional pleiotropy, we compared results with three other MR methods: MR Egger [49], weighted median [50], and weighted mode [51]. Each of these sensitivity methods makes different assumptions about pleiotropy, therefore a consistent direction and similar magnitude of effect provides more robust evidence for causal inference [52].

We assessed stability of the IVW estimates using SNP Leave-One-Out (LOO) analyses. We evaluated heterogeneity across the casual estimates of each individual genetic variant using the Cochran’s Q statistic [53]. We assessed heterogeneity for the MR-Egger analysis using the Rucker’s Q statistic [48] and examined the MR-Egger intercept term whereby a significant intercept term (non-zero) indicates overall directional pleiotropy [49]. We assessed degree of violation of the NOME (NO Measurement Error) assumption by calculating the 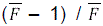 formula for IVW analyses and 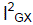 for MR-Egger analyses [53]. Increasing NOME violation (i.e. decreasing 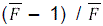 and 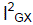) reduces power to detect directional pleiotropy and power to detect a causal effect for MR-Egger [53].

As there was some sample overlap between the GWASs for UI and anxiety and neuroticism (see supplementary table 7) we additionally implemented MRLap [54]. MRLap is a two-sample MR approach that simultaneously accounts for weak instrument bias and winner’s curse, while accounting for potential (known and unknown) sample overlap and its effects as a modifier of these biases. The framework provides a corrected causal effect estimate, that can be used as a sensitivity analysis to the main standard MR IVW. The degree of sample overlap is approximated using cross-trait LD-score regression (LDSC). For our analyses we assume that the degree of sample overlap does not differ between cases and non-cases.

### Software

Statistical analyses were performed using R software, version 4.3.2 (R Foundation for Statistical Computing, https://www.r-project.org/). The main packages used were *survival* (version 3.8.3), *survminer* (version 0.5.1), *TwoSampleMR* (version 0.6.22), *mr.divw* (version 0.1.0), and *MRlap* (version 0.0.3.3).

### Patient and public involvement

No patients were involved in setting the research question, selecting outcome measures, nor contributing towards developing plans for design or implementation of the study. No patients were asked to advise on interpretation or manuscript writing.

The current research was not informed by patient and public involvement because it used secondary data and had a lack of infrastructure, resources and funding to support patient involvement. However, future research following on from our findings should be guided by patient and public opinions. The results of the research will be disseminated to study participants on request, and to stakeholders and the broader public as relevant.

## RESULTS

The HES and EHR samples comprised 273 158 and 122 216 women respectively (see figure 1 and table 1) with a mean age of 56 years (SD 8.0). Demographic and health characteristics were similar across the two samples (table 1). Women were predominantly white (95%), cohabiting with a partner (69%) and around a third were educated to university degree (31%). The majority frequently consumed alcohol (62%) and 9% of women were current smokers. Sixty-eight percent of women had two or more children and 61% had reached menopause at baseline. After restricting to women who were complete cases for the study confounders, there were 262 961 (3.7% missing) and 118 526 (3.0% missing) women in the HES and EHR sample respectively. Characteristics were concordant between the complete cases and their respective HES/EHR samples.

The overall prevalence of UI, UI subtypes, depression, and anxiety is given in supplementary table 9. Briefly, in the combined EHR/HES sample, 10.6% of women had experienced AUI, 7.3% SUI, 2.6% UUI, 0.8% MUI, and 3.4% any urgency. Prevalence of depression was higher at 23.5% and anxiety at 16.5%. As expected prevalences in the EHR sample were higher compared to the HES sample, where cases may only present with a more severe phenotype. For example, AUI was diagnosed in 9.1% of women in the EHR sample compared to 3.6% of women in the HES sample. There was little difference in prevalences in the complete cases samples compared to the HES and EHR samples (supplementary table 9).

Mean follow-up years for the combined EHR/HES sample ranged from 7.1 to 7.4 years across all outcomes. After excluding prevalent cases at baseline for the outcomes in each model, the number at risk ranged from 98 075 to 117 840 (see figure 1). Kaplan Meier curves and a table of the numbers at risk across time are shown in supplementary figures 3 and 4. Evaluation of the proportional hazards assumption are shown in supplementary table 10 and supplementary figures 5 and 6.

### Bidirectional prospective analysis: mental health to urinary incontinence

We found strong evidence that women who experienced depression had an increased risk of new onset UI including any UI, and all UI subtypes (SUI, UUI, and MUI) (table 2). Associations were observed across all models (Models 1-3) and hereafter we describe results for Model 3 (fully adjusted for confounders). The strongest associations were observed for MUI (HR 1.91, 95%CI 1.59 to 2.31), followed by UUI (HR 1.80, 95%CI 1.63 to 1.98). We also found evidence that depression was prospectively associated with an increased risk of new onset of any urgency (irrespective of UI) (HR 1.53, 95%CI 1.41 to 1.65). There was also strong evidence that anxiety was associated with an increased risk of new onset UI (any UI and all subtypes) and any urgency. The strongest associations were observed for MUI (HR 1.64, 95%CI 1.24 to 2.17) and SUI (HR 1.46, 95%CI 1.25 to 1.69). We also found strong evidence that higher neuroticism scores were prospectively associated with an increased risk of new onset UI (any UI and all UI subtypes) and any urgency. The strongest association was observed for MUI where there was an 8% increase in risk of MUI per unit increase in neuroticism score (HR 1.08, 95%CI 1.04 to 1.12) (table 2).

**Table 2.**
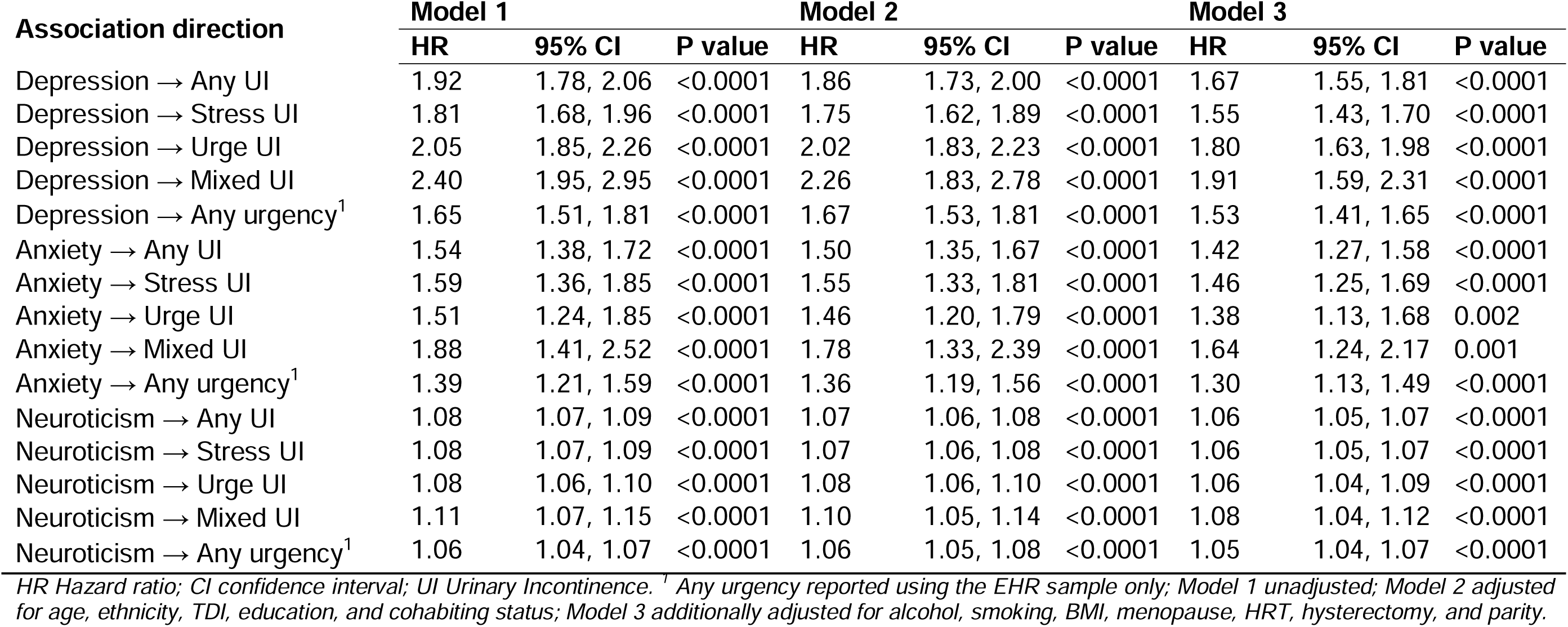
Cox regression analyses between mental health and urinary incontinence in UK Biobank women (N at risk between 110,371 and 117,840)

When exploring the associations within the EHR and HES samples separately (excluding any urgency as this was not available in the HES data), we observed the same pattern of associations, but the effect estimates were stronger in the HES sample (e.g. for depression to AUI: HR 1.87, 95%CI 1.55 to 2.24) compared to the EHR sample (e.g. for depression to AUI: HR 1.70, 95%CI 1.55 to 1.87) (see supplementary table 11).

### Bidirectional prospective analysis: urinary incontinence to mental health

We found strong evidence that women who experienced any UI, UI subtypes, and any urgency had an increased risk of new onset depression (table 3). Associations were observed across all models (Models 1-3) and hereafter we describe results for Model 3 (fully adjusted for confounders). The strongest associations were observed for MUI (HR 1.90, 95%CI 1.58 to 2.29), followed by UUI (HR 1.55, 95%CI 1.31 to 1.83). There was also strong evidence that any UI and UI subtypes were associated with new onset anxiety. The strongest associations were observed for MUI (HR 1.58, 95%CI 1.18 to 2.12) and UUI (HR 1.34, 95%CI 1.02 to 1.75). There was weak evidence of an association between any urgency and new onset anxiety (HR 1.30, 95%CI 0.99 to 1.71).

**Table 3.**
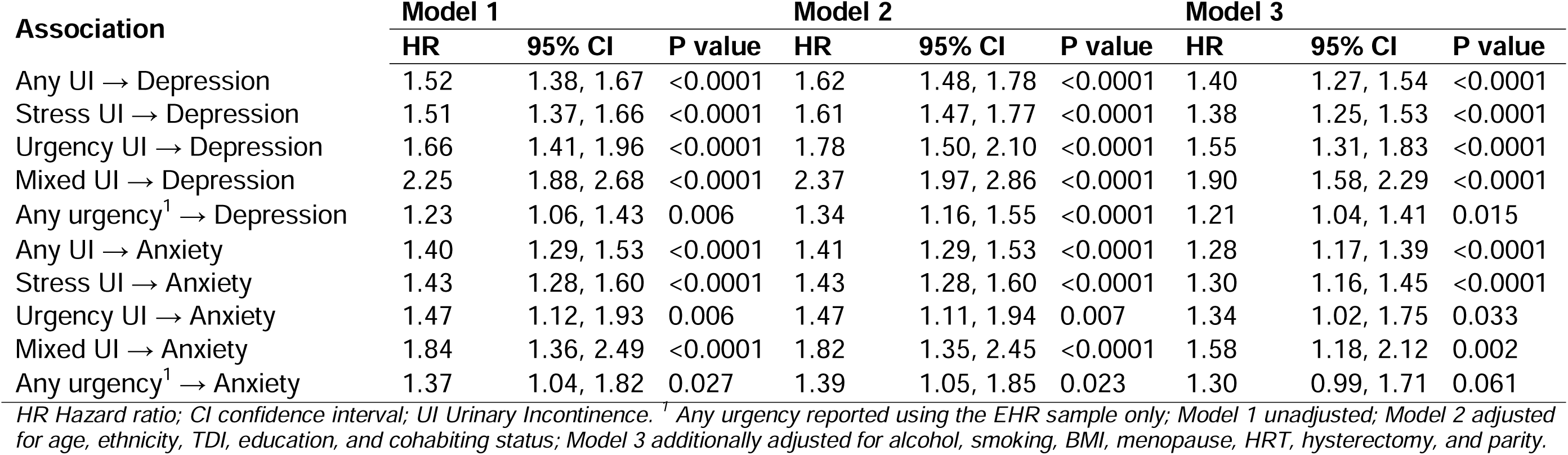
Cox regression analyses between urinary incontinence and mental health problems in UK Biobank women (N at risk between 98,075 and 107,496)

| Association | Model 1 |  |  | Model 2 |  |  | Model 3 |  |  |
| --- | --- | --- | --- | --- | --- | --- | --- | --- | --- |
|  | HR | 95% CI | P value | HR | 95% CI | P value | HR | 95% CI | P value |
| Any UI → Depression | 1.52 | 1.38, 1.67 | <0.0001 | 1.62 | 1.48, 1.78 | <0.0001 | 1.40 | 1.27, 1.54 | <0.0001 |
| Stress UI → Depression | 1.51 | 1.37, 1.66 | <0.0001 | 1.61 | 1.47, 1.77 | <0.0001 | 1.38 | 1.25, 1.53 | <0.0001 |
| Urgency UI → Depression | 1.66 | 1.41, 1.96 | <0.0001 | 1.78 | 1.50, 2.10 | <0.0001 | 1.55 | 1.31, 1.83 | <0.0001 |
| Mixed UI → Depression | 2.25 | 1.88, 2.68 | <0.0001 | 2.37 | 1.97, 2.86 | <0.0001 | 1.90 | 1.58, 2.29 | <0.0001 |
| Any urgency<sup>1</sup> → Depression | 1.23 | 1.06, 1.43 | 0.006 | 1.34 | 1.16, 1.55 | <0.0001 | 1.21 | 1.04, 1.41 | 0.015 |
| Any UI → Anxiety | 1.40 | 1.29, 1.53 | <0.0001 | 1.41 | 1.29, 1.53 | <0.0001 | 1.28 | 1.17, 1.39 | <0.0001 |
| Stress UI → Anxiety | 1.43 | 1.28, 1.60 | <0.0001 | 1.43 | 1.28, 1.60 | <0.0001 | 1.30 | 1.16, 1.45 | <0.0001 |
| Urgency UI → Anxiety | 1.47 | 1.12, 1.93 | 0.006 | 1.47 | 1.11, 1.94 | 0.007 | 1.34 | 1.02, 1.75 | 0.033 |
| Mixed UI → Anxiety | 1.84 | 1.36, 2.49 | <0.0001 | 1.82 | 1.35, 2.45 | <0.0001 | 1.58 | 1.18, 2.12 | 0.002 |
| Any urgency<sup>1</sup> → Anxiety | 1.37 | 1.04, 1.82 | 0.027 | 1.39 | 1.05, 1.85 | 0.023 | 1.30 | 0.99, 1.71 | 0.061 |

HR Hazard ratio; CI confidence interval; UI Urinary Incontinence. <sup>1</sup> Any urgency reported using the EHR sample only; Model 1 unadjusted; Model 2 adjusted for age, ethnicity, TDI, education, and cohabiting status; Model 3 additionally adjusted for alcohol, smoking, BMI, menopause, HRT, hysterectomy, and parity.

Effect estimates for each of the analyses slightly increased for Model 2 (adjusted for socio-demographic factors) compared to Model 1 (unadjusted) but then attenuated for Model 3 (fully adjusted) (see table 3). When exploring the associations within the EHR and HES samples separately (excluding any urgency as this was not available in the HES data), we observed the same pattern of associations, but the effect estimates were greater in the HES sample (e.g. for AUI to depression: HR 1.54, 95%CI 1.39 to 1.71) compared to the EHR sample (e.g. for AUI to depression: HR 1.45, 95%CI 1.29 to 1.64) (see supplementary table 12).

### Bidirectional prospective analysis: sensitivity analyses

In sensitivity analyses (see supplementary table 13), results were consistent when stratifying on confounders that violated the proportional hazards assumption (all differences in HRs <0.04); when excluding events within the first two years of follow-up (all differences in HRs <0.44); and when excluding cases of other psychiatric disorders (all differences in HRs <0.09). For analyses where SUI and UUI cases were mutually excluded, HRs were consistent with the primary analysis (all differences in HR <0.07). Finally, for the mutual adjustment in analyses of depression and anxiety (Model 4) (e.g. the AUI to depression model was additionally adjusted for anxiety in addition to confounders) we observed little change in HRs for depression mutually adjusted for anxiety (all differences <0.10) and for anxiety mutually adjusted for depression (all differences <0.26) when comparing to Model 3 estimates (see supplementary table 14).

### Bidirectional two-sample Mendelian Randomisation

The estimated IVW ORs for AUI and its subtypes per doubling in the genetic liability to depression and anxiety and per unit increase in neuroticism score are presented in figure 2 and supplementary table 15. We found evidence for causal effects of depression and neuroticism on AUI, SUI, UUI, and MUI (e.g. for depression and AUI: IVW-OR 1.25, 95%CI 1.16 to 1.35; see figure 2). The magnitude of causal effects were larger for UUI and MUI compared to AUI and SUI for the exposure’s depression and neuroticism, although the 95% CIs are wider. Conversely, we found little evidence for causal effects of anxiety on any of the UI subtypes as all 95% CI crossed the null (e.g. for AUI IVW-OR 1.03, 95%CI 0.99 to 1.06).

**Figure 2.**
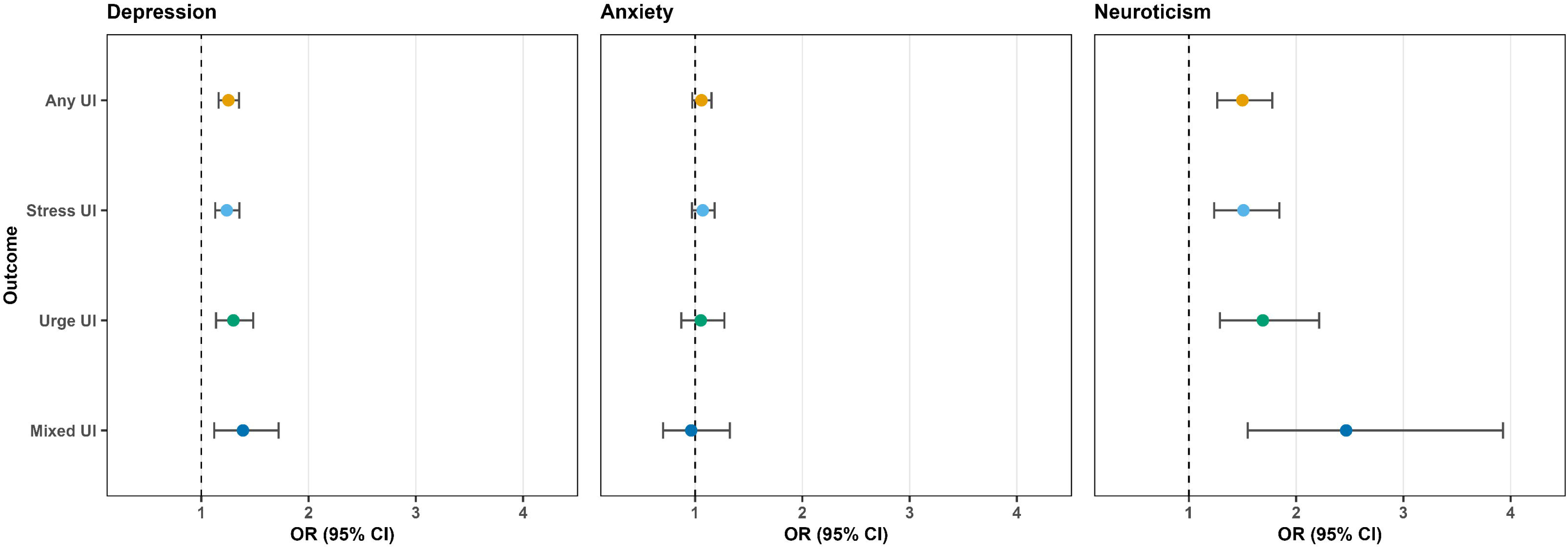
Mendelian randomisation of depression, anxiety, and neuroticism with any urinary incontinence and its subtypes. UI urinary incontinence; OR odds ratio; CI confidence interval. Causal effects represent the odds of any UI, stress UI, urge UI and mixed UI with the doubling of odds in the genetic liability to the exposures of depression and anxiety and per unit increase in neuroticism score.

The estimated db-IVW ORs for depression/anxiety and Betas for neuroticism per doubling in the genetic liability to AUI and its subtypes are presented in figure 3 and supplementary table 16. We found weak evidence for a causal effect of genetic liability to AUI on depression (IVW-OR 1.02, 95%CI 1.00 to 1.03; see figure 3) and for SUI on depression (IVW-OR 1.01, 95%CI 1.00 to 1.02) and neuroticism (IVW-Beta 0.01, 95%CI 1.4E-4 to 0.02), although the effect estimates for neuroticism are small. There was no evidence for causal effects of UUI and MUI on depression/anxiety or neuroticism. Inference remained the same when observing the individual Wald ratios (supplementary table 17)

**Figure 3.**
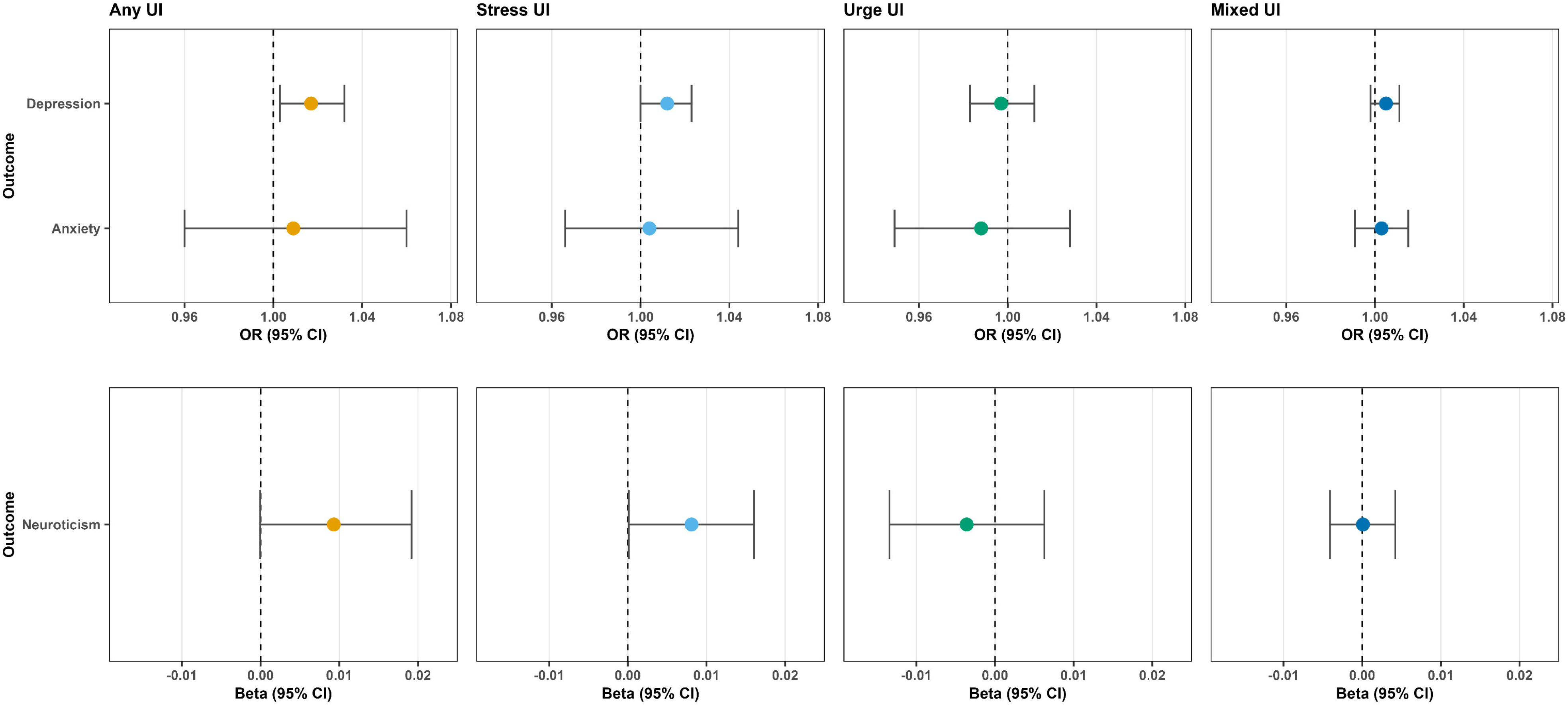
Mendelian randomisation of the genetic liability to urinary incontinence with depression, anxiety, and neuroticism outcomes. UI urinary incontinence; OR odds ratio; CI confidence interval. Causal effects represent the odds of depression and anxiety and the beta of neuroticism score with the doubling of odds in the genetic liability to the exposures of any UI, stress UI, urge UI and mixed UI.

### Appraisal of the robustness of MR estimates

#### For causal effects from mental health and neuroticism to urinary incontinence

All genetic instruments for exposures had a mean F-statistic >35 suggesting that weak instrument bias was unlikely to affect interpretation (see supplementary table 18). Leave-one-out analysis suggested that no one SNP was influencing the interpretation of results (see supplementary figure 7). Sensitivity analyses showed consistent estimates between the IVW, MR-Egger, weighted-median and weighted-mode approaches, although 95%CIs were wider given the lower statistical power of these sensitivity analyses (supplementary table 15). Strong evidence remained for a causal effect between depression and AUI for the MR-Egger and weighted-median approach. There was also a consistent direction of effect for the analyses between anxiety and AUI, SUI and UUI, however the 95%CI widened, and the direction of effect flipped for MUI MR-Egger and weighted mode analyses. A similar pattern was observed for analyses exploring the causal effects of neuroticism score with UI subtypes.

Cochran’s Q and Rucker’s Q values suggest heterogeneity in the SNP-exposure/SNP-outcome associations between mental health exposures and UI outcomes, which may indicate pleiotropy (supplementary table 18). However, all 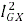 were low (≤14) indicating potential attenuation of effect estimates due to violation of the NOME assumption. However, all MR-Egger intercept P values were >0.10, suggesting little evidence of overall directional pleiotropy (see supplementary table 18). All 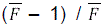 were ≥0.97 indicating that the degree of NOME assumption violation was minimal for the IVW results.

Results for the MRLap analysis were concordant with the main IVW results suggesting that sample overlap between the GWAS for UI and anxiety/neuroticism is unlikely to impact on causal inference (see supplementary table 19).

#### For causal effects from urinary incontinence to mental health and neuroticism

All genetic instruments for exposures had a mean F-statistic >22 suggesting that weak instrument bias was unlikely to affect interpretation (see supplementary table 18). Leave-one-out analysis suggested that no one SNP was influencing the interpretation of results (see supplementary figure 7). For the effects of UI subtypes on depression and anxiety, sensitivity analyses showed similar estimates between the db-IVW, MR-Egger, weighted-median and weighted-mode approaches, although 95%CIs were wider given the lower statistical power of these sensitivity analyses (supplementary table 16). Effect estimates for the liability to UI on neuroticism were inconsistent between methods, although estimates are very small.

Cochran’s Q and Rucker’s Q values suggest heterogeneity in the SNP-exposure/SNP-outcome associations between AUI/SUI and depression/neuroticism, which may indicate pleiotropy (supplementary table 18). There was little evidence of heterogeneity between the UI subtypes and anxiety as well as UUI/MUI and depression/anxiety. All 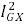 were low (≤0.86) indicating potential attenuation of effect estimates due to violation of the NOME assumption. However, all MR-Egger intercept P values were >0.10, suggesting little evidence of overall directional pleiotropy (see supplementary table 18). All 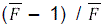 were ≥0.95 indicating that the degree of NOME assumption violation was minimal for the IVW results.

## DISCUSSION

In this study we triangulated evidence from multivariable prospective (time-to-event) analyses with two-sample MR to make inferences about the bidirectional causal effects of depression/anxiety and neuroticism and urinary incontinence subtypes in women from the UK Biobank.

In the time-to-event analysis we found strong evidence that depression/anxiety and neuroticism are associated with an increased risk of new onset UI (any UI and UI subtypes) and any urgency (irrespective of urinary leakage). In two-sample MR, we found strong evidence for a causal effect of genetic liability to depression and neuroticism, but not anxiety, on UI and its subtypes.

When we examined relationships in the opposite direction, we found that any UI and all UI subtypes were associated with an increased risk of new onset depression and anxiety. Urgency (irrespective of leakage) was associated with new onset depression only. In MR analysis we found evidence of a causal effect of any UI on depression and a causal effect of SUI on depression and neuroticism, although the effect estimates for neuroticism were small. We found no evidence of causal effects of UUI and MUI on depression, anxiety, or neuroticism.

### Strengths and limitations

This is the largest population-based cohort study of bidirectional relationships between mental health problems and UI. It is also the first study, to our knowledge, to examine associations between mental health problems and UI using linked health records to identify UI subtypes. Few previous studies have distinguished between UI subtypes but instead have asked women to self-report any occurrence of urinary leakage.

Another strength of this study is the integration of multiple data resources (UK Biobank and large mental health consortia) to triangulate evidence from two different approaches with different sources of bias [52]. To improve causal inference, we adjusted for a wide range of confounders, and we conducted a bidirectional two-sample MR to test for causal effects. Our primary analysis sample combined data from EHR and HES, but we also conducted separate analyses in EHR and HES, showing that associations were stronger in the hospital sample. We also performed the largest GWAS of UI and UI subtypes in women to date by defining clinically diagnosed UI in a large Biobank setting (e.g. AUI n=118 783; 16 649 cases). Previous GWAS have used self-report data [55,56] which may lead to misclassification of the UI subtypes, as well as making adjustments for other factors such as BMI and parity [56]. Adjustment for covariates in GWAS may bias MR analyses depending on the underlying causal structure and should generally be avoided [57].

Limitations should also be considered when interpreting these findings. We restricted our analyses to UK Biobank women with linked EHR primary care data (45% of the total cohort). This may have introduced selection bias; however, we anticipate this to be minimal given that the availability of EHR data was dependent on the data supplier for the participants primary care practice rather than any participant-level factors. We also explored the associations between the missing data mechanism and outcomes conditional on confounders and found little evidence that these participant-level factors predicted missing primary care data.

We attempted to mitigate sources of bias such as confounding and reverse causation in the prospective analysis by adjusting for a wide range of confounders and conducting sensitivity analyses by excluding those who had new onset outcomes within the first two years of follow-up time. Nonetheless, residual or unmeasured confounding and selection bias may have affected our effect estimates, for instance we did not consider any medications as potential confounders as it is difficult to link prescriptions as a cause of symptoms [58]. It is also possible that there was over-adjustment for certain health and lifestyle factors (e.g. BMI and alcohol intake) as these may lie on the causal pathway between exposure and outcome (i.e. depression to UI). We therefore report results both minimally adjusted for sociodemographic factors (Model 2) and additionally adjusted for health and lifestyle factors (Model 3) where we report results that were concordant across models.

There is potential for misclassification in the UI subtypes due to incorrect Read code recording or complex symptomology. Women may also have symptoms of UI but nevertheless do not seek help with their GPs [59]. This may be particularly relevant for women with depression due to its impact on care seeking and treatment adherence more broadly [60–62]. There may also be bias due to misclassification of urge UI. While there is a specific ICD-10 code for stress UI, urge UI is contained within the definition of code N39.4 (other specified urinary incontinence) along with reflex and overflow incontinence. It is possible that a small proportion of urge UI cases in the HES samples comprise women with reflex or overflow UI. However, these two subtypes are rare in women and any misclassification will be minimal [63].

There are also limitations regarding the MR. The GWASs for depression, anxiety, and neuroticism were conducted on samples of males and females, whereas the GWAS for UI and its subtypes was conducted on women only. This may cause bias as the association of the genetic variants with the exposure may not be replicated in the set of individuals where the outcome is estimated [64]. There is also some sample overlap between the GWASs for UI subtypes and anxiety and neuroticism. This may bias the MR causal estimates towards the confounded observational estimates. Bias is likely to be minimal when strong instruments are used (i.e. F-statistic >10; [65]. In sensitivity analysis we report similar effect estimates when using MRlap to correct for sample overlap.

Finally, our study is also limited by the populations in which GWAS have been conducted which were predominantly in those of European ancestry. While this reduces the risk of confounding by population stratification, it means our study, and other MR studies, cannot be generalised to other populations.

### Comparison with previous findings

Our findings are consistent with previous studies that have found evidence that depression [10–12,17] and neuroticism [13] are associated with an increased risk of UI (any self-reported UI irrespective of subtype). Fewer studies have examined the prospective association between anxiety and UI. One study reported an association between anxiety and an increased risk of any UI, but this association was attenuated following adjustment for depression (Joinson et al. 2025). When we examined UI subtypes, there was strong evidence that depression, anxiety and neuroticism are associated with new onset SUI, UUI and MUI. A previous study found that genetic liability to depression, anxiety, and neuroticism is associated with any UI, MUI and urinary urgency, but not with SUI [18].

We found evidence that the relationships between depression, anxiety and UI are bidirectional, with UI (any UI and all subtypes) being associated with an increased risk of new onset depression and anxiety. A previous study reported bidirectional relationships between UUI and SUI and mental health problems (depression and anxiety) [14,15] whilst other studies have found inconsistent results when examining bidirectional associations between UI subtypes, depression, and anxiety [16,17]. Inconsistent findings may be due to the use of self-report UI and questionnaires assessing depressive/anxiety symptoms, whereas our study used diagnoses from linked health care records.

We also found evidence that depression, anxiety, and neuroticism were prospectively associated with any urgency (irrespective of urinary leakage) and that any urgency was associated with new onset depression (but not anxiety). Cross-sectional studies have found evidence that overactive bladder (characterized by urinary urgency, with or without urinary incontinence) is associated with higher levels of depression and anxiety [9]. A prospective cohort study found evidence that depression, but not anxiety, was associated with new onset urgency, but urgency was not associated with new onset depression [17].

Using MR analysis, we found some evidence of bidirectional causal effects between depression, neuroticism, and UI, but not between anxiety and UI. This conflicts with the results from the time-to-event analysis and could be due to a lack of power because the anxiety GWAS had a smaller sample size compared with the GWAS for depression and neuroticism. Causal effects were strongest in the direction of depression/neuroticism to UI whilst there was less consistent evidence of causal effects in the opposite direction. It is possible that the occurrence of UI may not affect new onset mental health many years later [66] and the GWAS associations represent lifetime exposure to UI/mental health. Notably, one study found little evidence of prospective associations between UI subtypes and depression or anxiety after a follow–up period of approximately eight years (only SUI was associated with incident depression) [17].

A cross-sectional study in over 29 000 males and females reported that mental health problems were associated with a higher risk of UI and overactive bladder, and performed a two-sample MR reporting causal effects of anxiety on overactive bladder, and neuroticism on urinary frequency/incontinence [67]. However, the GWASs for mental health and lower urinary tract symptoms (LUTS) were performed on UK Biobank participants, creating substantial sample overlap, and included both sexes. Furthermore, the self-reported urinary frequency/incontinence variable is non-specific for UI nor its subtypes with a prevalence considerably lower than those using linked health records (n=1624; 0.4%) [67]. In contrast to our findings, a recent two-sample MR found little evidence of causal bidirectional relationships between UI and anxiety, depression, and neuroticism [68]. However, the GWASs available for any UI, SUI, UUI and MUI were conducted on a smaller sample of women using self-reported questionnaire data which may have led to misclassification and low statistical power. This highlights the need for further larger GWAS of UI subtypes.

### Potential mechanisms explaining the findings

The well-documented adverse effects of UI on health-related quality of life and the perceived stigma of incontinence have negative consequences for mental health [69]. Incontinence can also cause maladaptive cognitions and mental health problems, due to its stigmatised and chronic nature and the sense of frustration, hopelessness, and loss of control experienced by affected people [70]. There is some evidence from randomised controlled trials of improvements in mental health following successful treatment of UI [71–73].

Biopsychosocial mechanisms that underlie the effects of mental health problems on UI are less well documented. Mechanisms are thought to involve dysregulated stress responses and chronic inflammation [66,74–76]. Depression and anxiety are associated with altered functionality of the hypothalamic–pituitary–adrenal (HPA)-axis, with consequent effects on serotonin and cortisol levels that have been found to impact brain regions controlling bladder [75,77]. Studies have examined if drugs aimed at increasing serotonin levels have effects on urinary symptoms but have yielded inconsistent findings. One study reported an improvement in overactive bladder symptoms [78], one found an increased risk of UI [15], and another found that SSRIs are unlikely to cause or worsen stress UI [79]. There is a need for more research aimed at advancing understanding of how serotonin affects urinary symptoms and for RCTs examining the effectiveness of antidepressants in treating UI.

### Clinical implications and future directions

UI is often chronic and treatment resistant, resulting in reduced quality of life, increased healthcare utilisation, and high NHS costs. Mental health problems are not routinely assessed in urology clinics, as they do not feature in National Institute for Health and Care Excellence (NICE) guideline recommendations, and failure to recognise their contribution to the onset and persistence of UI could be a cause of low success rates of existing treatments for UI. Research is needed to advance understanding of the biopsychosocial mechanisms that underlie the causal effects of mental health problems on UI. The findings could lead to the identification of novel translational targets and therapeutic advances in the treatment of UI. Evidence of causal effects of mental health problems on UI could provide an impetus for research aimed at examining if treatments for depression/anxiety and chronic inflammation could be effective in alleviating UI. More research is needed to establish if there are distinct or shared causal pathways associated with the onset and progression of UI subtypes because this knowledge could potentially lead to more targeted, predictive and preventative care.

## Data Availability

Data for UK Biobank may be obtained from a third party and are not publicly available. Researchers can apply to use the UK Biobank resource for health-related research that is in the public interest (https://www.ukbiobank.ac.uk/use-our-data/apply-for-access/).

## Author contributions

CJ conceptualised the study. KB, MD, KT, CJ contributed towards the analytical plan. MD provided advise on medical coding. KB carried out analysed and interpreted study data. KT, TP, RK provided statistical and data advise. KB, CJ wrote the initial manuscript. KB, CJ, KT, MD, TP, RK contributed to the revision of the manuscript. KB and CJ are the study’s guarantors, having full access to all data included in the study and taking responsibility for its integrity and the accuracy of the analysis. All authors involved have carefully reviewed and given their approval for the final draft of the manuscript. The corresponding author attests that all listed authors meet authorship criteria and that no others meeting the criteria have been omitted.

## Copyright/license for publication

The Corresponding Author has the right to grant on behalf of all authors and does grant on behalf of all authors, *a worldwide licence* to the Publishers and its licensees in perpetuity, in all forms, formats and media (whether known now or created in the future), to i) publish, reproduce, distribute, display and store the Contribution, ii) translate the Contribution into other languages, create adaptations, reprints, include within collections and create summaries, extracts and/or, abstracts of the Contribution, iii) create any other derivative work(s) based on the Contribution, iv) to exploit all subsidiary rights in the Contribution, v) the inclusion of electronic links from the Contribution to third party material where-ever it may be located; and, vi) licence any third party to do any or all of the above.

## Code sharing

The code to conduct these analyses can be found at https://github.com/burrowsk/Mental-health-and-urinary-incontinence-in-UK-Biobank-women upon publication.

## Acknowledgement

We thank Professor Nikki Cotterill for her guidance in primary and secondary care coding of urinary incontinence and its subtypes. This research has been conducted using the UK Biobank resource under approved application number 73737 (Prof. Carol Joinson). This work uses data provided by patients and collected by the NHS as part of their care and support.

## Ethics approval

The UK Biobank study was approved by the NHS National Research Ethics Service (Ref. 11/NW/0382). All UKBB participants have provided written informed consent, and detailed research ethics approval can be found at https://www.ukbiobank.ac.uk/learn-more-about-uk-biobank/about-us/ethics.

