## supplementary methods for "Depression and anxiety as causes and consequences of urinary incontinence in women: a population-based study"

<u>Electronic health records for primary care data</u>
To date, primary care data (EHR) have been obtained for 45% of the UK Biobank cohort (~230,000 participants). Although all participants have given consent for linkage to electronic health records, the availability of EHR data is dependent on various data suppliers and computer system suppliers. Table A gives an overview of the GP data providers by country and the relevant coding classification associated with that provider.

Table A: code classifications by GP data provider

| Country | GP data provider | Clinical coding classification | N (approx.) of UK participants |
|---|---|---|---|
| England | TTP | Clinical Terms Version 3 (CTV3) | 165,000 |
| England | Vision | Read v2 | 18,000 |
| Scotland | EMIS/Vision | Read v2 | 27,000 |
| Wales | EMIS/Vision | Read v2 | 21,000 |

Read codes represent standardised clinical terms used in primary care since 1985. There are two versions: Read v2 and CTV3 (or Read v3). In addition to clinical codes, information on GP registrations is also provided. This data indicates registration and deregistration to multiple GPs, where applicable. However, this information varies by data provider where England's Vision provided a single registration record per person, while others provided multiple records. There was also a small number of participants with data in the TPP extract that do not have a registration record.

Another limitation of the EHR data is that the start of coverage is not known for all participants, and different approaches to extracts meant that end of follow-up dates varied by supplier. More information about the linkage of primary care records can be found at:

https://biobank.ndph.ox.ac.uk/showcase/showcase/docs/primary_care_data.pdf.

<u>Darke algorithm and data cleaning of health linkage records</u>
Due to the above-described limitations of EHR data, we implemented a modified version of the Darke algorithm (Darke et al. 2021). Briefly, this included the following steps:

1. Create censor dates as either:
    a. Data extract start (date of data collection)
    b. Date of death (whichever is earlier).
    c. If the deregistration date is missing, assume that the participant was still registered at the time of data extract. Replace this with the censor date.
2. Drop registration dates that do not meet the following criteria:
    a. Missing registration dates
    b. Starts after censor date
    c. Pre-birth registration
    d. Future registrations
    e. Pre-birth deregistration dates
3. Reconcile conflicting or overlapping registration periods
    a. There may also be participants who have clinical events occurring outside the registration period
    b. Apply the Darke algorithm to extend registration periods to events, thus extending the follow-up period.

*These tasks aimed to censor participants at the earlier of the inferred end of data collection, the data extract date, or the date of death.*

4. Additional exclusions of EHR data includes participants who had an invalid follow-time whereby their censor date (study end for the cox proportional hazards models) occurred prior to baseline giving negative follow-up times (N = 1,190 women). Participants who did not have registrations data were also excluded (N = 2,294 women).

Time-to-event analysis filtering and exclusions

Prior to running each individual Cox Proportional Hazard model, we excluded any participants who had a diagnosis of the outcome prevalent at baseline. This resulted in variable numbers of participants at risk (see Figure 2 in the main manuscript).

As sensitivity analyses, we implemented a series of restrictions and exclusions to test the robustness of our associations. First, we stratified on several confounders to allow the proportional hazards assumption to be met. This is described in the main manuscript. Second, we restricted our models by excluding incident cases within the first two-years of follow-up to reduce potential reverse causation. Third, we excluded participants with self-reported psychiatric conditions. Cases were derived from multiple data sources including self-report non-cancer illnesses (field code 20002), the Mental Health Questionnaire (MHQ) sub study (field code 20544), and Hospital Episode Statistics (HES) data (field code 21022). This resulted in excluding 5231 (2%) participants with history of one or more of schizophrenia, mania/bipolar, alcohol abuse, opioid abuse, and other substance abuse. For the fourth sensitivity analyses we mutually excluded prevalent SUI and UUI cases from their respective analyses as described in the main manuscript. This resulted 1307 participants with UUI being excluded from analyses of SUI, and 6107 participants with SUI being excluded from analyses of UUI.

Missing data

We explored whether any of the outcomes (both urinary incontinence and mental health) derived from the HES sample (diagnosed in hospital, available for the entire cohort) were associated with missing data in the EHR sample (where only 45% of the cohort is available). All measures showed no evidence for an association with the missing data (all P values >0.08) except for anxiety (P value 0.001). In addition, we reported no evidence of an association between self-report "frequency/incontinence" [field code 20002; N cases 1,155; N non-cases 272,003] and missing data in the EHR sample (see Table B below).

Table B: ORs and 95% CI for the association between outcomes and missing data for EHRs (primary care data)

| Outcome measured in HES sample | OR (95% CI) | P value |
|---|---|---|
| Self-report "frequency/incontinence" | 1.07 (0.96, 1.20) | 0.231 |
| Any UI | 0.99 (0.95, 1.03) | 0.715 |
| Stress UI | 1.00 (0.96, 1.05) | 0.924 |
| Urge UI | 0.93 (0.86, 1.01) | 0.075 |
| Mixed UI | 0.92 (0.81, 1.04) | 0.168 |
| Depression | 1.00 (0.97, 1.03) | 0.979 |
| Anxiety | 0.95 (0.92, 0.98) | 0.001 |

UI urinary incontinence; OR odds ratio; HES hospital episode statistics

GWAS of urinary incontinence traits

GWAS of the UK Biobank urinary incontinence traits was performed using an established analysis pipeline described in: Quality control filtering of the UK Biobank genetic data was conducted by Mitchell *et al.* as described in the published protocol (doi:10.5523/bris.1ovaau5sxunp2cv8rcy88688v) (Mitchell, Hemani, et al. 2019). The MRC IEU UK Biobank GWAS pipeline was developed by Mitchell *et al.* (doi:10.5523/bris.pnoat8cxo0u52p6ynfaekeigi.) (Mitchell, Elsworth, et al. 2019) .

Briefly, GWAS was conducted using a linear mixed model (LMM) association method as implemented in BOLT-LMM (v2.3) (Loh et al. 2015). To model population structure in the sample we used 143,006 directly genotyped SNPs, obtained after filtering on MAF > 0.01; genotyping rate > 0.015; Hardy Weinberg equilibrium P value < 0.0001 and LD pruning to an $r^2$ threshold of 0.1 using PLINKv2.00. Models were adjusted for genotyping array and age.

BOLT-LMM association statistics are on the linear scale. The effect estimates from this analysis can therefore be interpreted as the change in disease risk per copy of the effect allele. These results can be converted to log odds ratios using a Taylor transformation expansion series (Loh et al. 2018).

*Conversion of test statistics to the log odds ratio scale*

Betas and SEs can be converted to approximate log odds ratios using the following R code as an example:

**# Convert BOLT LMM effects to log odds**

**# formula: log OR = beta / (u(1-u)); where u=ncases/(ncases + ncontrol) REPEAT with SE**

**# ukbb_all is a data-frame of GWAS summary statistics**

**ukbb_all$ncase <- 16649**

**ukbb_all$ncontrol <- 102134**

**ukbb_all$u <- ukbb_all$ncase/(ukbb_all$ncase + ukbb_all$ncontrol)**

**ukbb_all$beta <- ukbb_all$beta/ (ukbb_all$u * (1 - ukbb_all$u))**

**ukbb_all$se <- ukbb_all$se / (ukbb_all$u * (1 - ukbb_all$u))**

We also excluded SNPs with unreliable minor allele frequencies in cases as per Howrigan et al (Neale Lab 2017) . It is known that the use of linear models to test genetic associations with binary phenotypes can lead to inflated false positive findings, especially for rare variants in analyses with a small number of cases compared to controls. To mitigate the impact of model misspecification on our results, we therefore removed SNPs with a minor allele frequency less than the following threshold:

$MAF\ threshold = 25 / (2 * case\ sample\ size)$
