## supplementary table for "Depression and anxiety as causes and consequences of urinary incontinence in women: a population-based study"

Table 1 Diagnosis codes used to define urinary incontinence and its subtypes

| **UI subtype** | **HES sample codes** | | | **EHR sample codes** | | | **Comments** |
|---|---|---|---|---|---|---|---|
| **Stress UI (SUI)** | ICD-9 | 625.6 | Stress urinary incontinence | Read v2 | K198. | Stress incontinence | NA |
| | ICD-10 | N39.3 | Stress incontinence (female) | Read v2 | K586. | Stress incontinence - female | |
| | | | | CTV3 | 1A24. | Stress incontinence | |
| | | | | CTV3 | K198. | Genuine stress incontinence | |
| | | | | CTV3 | K586. | Stress incontinence - female | |
| | | | | CTV3 | 1A24. | Stress incontinence (& symptom) | |
| | | | | CTV3 | X30OL | Urinary sphincter weakness incontinence | |
| | | | | CTV3 | XE0rR | Genuine stress incontinence | |
| | | | | CTV3 | XaZQg | Sneezing incontinence of urine | |
| | | | | CTV3 | XE0gn | Stress incontinence (& [female]) | |
| | | | | CTV3 | .J697 | Stress incontinence (& [female]) | |
| | | | | CTV3 | .1A24 | Genuine stress incontinence | |
| **Urge UI (UUI)** | ICD-9 | 788.3 | Other specified urinary incontinence | Read v2 | R0832 | [D] Urge incontinence | NA |
| | ICD-10 | N39.4 | Incontinence of urine | Read v2 | 1A26. | Urge incontinence of urine | |
| | | | | CTV3 | R0832 | [D] Urge incontinence | |
| | | | | CTV3 | 1A26. | Urge incontinence of urine | |
| | | | | CTV3 | X30OG | Urge incontinence of urine | |
| **Mixed UI (MUI)** | Case for SUI **and** UUI | | | Case for SUI **and** UUI | | | Diagnosis of SUI and UUI can occur at different times |
| **Any UI (AUI)** | Case for SUI **and/or** UUI | | | Case for SUI **and/or** UUI **and/or:** | | | For the EHR sample, there are additional Read codes that are specific for general UI (i.e. not a specific subtype). These are additionally included in the case definition. |
| | | | | Read v2 | 1A23. | Incontinence of urine | |
| | | | | Read v2 | R083. | [D]Incontinence of urine | |
| | | | | CTV3 | 1A23. | Incontinence of urine | |
| | | | | CTV3 | R083. | [D]Incontinence of urine | |
| | | | | CTV3 | R083z | [D]Incontinence of urine NOS | |
| | | | | CTV3 | .1A23 | Urinary incontinence | |
| | | | | CTV3 | .R83. | [D]Incontinence of urine | |
| **Any urgency (with or without UI)** | NA | | | Read v2 | 1A25. | Urgency | There are no specific ICD codes available in UK Biobank HES data |
| | | | | Read v2 | R0862 | [D]Urgency of micturition | |
| | | | | Read v2 | 1A35. | Precipitancy of micturition | |
| | | | | CTV3 | 1A25. | Urgent desire to urinate | |

| UI subtype | HES sample codes | EHR sample codes | | | Comments |
|---|---|---|---|---|---|
| | | CTV3 | R0862 | [D]Urgency of micturition | to enable derivation of this subtype. For the EHR sample, cases are those with urgency specific Read codes irrespective of whether a participant is a case for UI or not. |
| | | CTV3 | .1A25 | Urgent desire to urinate | |
| | | CTV3 | 1A35. | Urgent desire to urinate | |
| | | CTV3 | .1A35 | Urgent desire to urinate | |

*UI urinary incontinence; HES hospital episode statistics, the sample who have linked hospital secondary care data; EHR electronic health records, the sample who have linked primary care records data; ICD-9 International Classification of Diseases, 9th Revision; ICD-10 International Classification of Diseases, 10th Revision*

Table 2 Matrix of case definitions for urinary incontinence subtypes

<table>
<tr><td></td><td></td><td colspan="2">UUI</td></tr>
<tr><td></td><td></td><td>Yes</td><td>No</td></tr>
<tr><td rowspan="2">SUI</td><td>Yes</td><td>MUI<br>[SUI and UUI]<br>[AUI]</td><td>SUI only<br>[AUI but not MUI]</td></tr>
<tr><td>No</td><td>UUI only<br>[AUI but not MUI]</td><td>No AUI<br>[no SUI and no UUI]</td></tr>
</table>

*AUI any urinary incontinence (UI); SUI stress UI; UUI urge UI; MUI mixed UI*.

Table 3 Primary care Read codes for depression

| Read v2 | Term v2 | CTV3 | Term-CTV3 |
|---|---|---|---|
| E204. | neurotic depression reactive type | XE1YC | reactive depression |
| Eu323 | severe depressive episode with psychotic symptoms | XE1ZZ | severe depressive episode with psychotic symptoms |
| E1120 | single major depressive episode unspecified | E1120 | single major depressive episode unspecified |
| E1123 | single major depressive episode severe without mention of psychosis | E1123 | single major depressive episode severe without mention of psychosis |
| Eu329 | single major depressive episode severe with psychosis in remission | XaX53 | single major depressive episode severe with psychosis in remission |
| E1124 | single major depressive episode severe with psychosis | E1124 | single major depressive episode severe with psychosis |
| E112z | single major depressive episode nos | E112z | single major depressive episode nos |
| E1122 | single major depressive episode moderate | E1122 | single major depressive episode moderate |
| E1121 | single major depressive episode mild | E1121 | single major depressive episode mild |
| E1125 | single major depressive episode in partial or unspecified remission | E1125 | single major depressive episode in partial or unspecified remission |
| E1126 | single major depressive episode in full remission | E1126 | single major depressive episode in full remission |
| Eu32. | depressive episode | XE1Y0 | single major depressive episode |
| E112. | single major depressive episode | XE1Y0 | single major depressive episode |
| Eu327 | major depression severe without psychotic symptoms | XSGom | severe major depression without psychotic features |
| Eu328 | major depression severe with psychotic symptoms | XSGon | severe major depression with psychotic features |
| Eu322 | severe depressive episode without psychotic symptoms | XE1ZY | severe depressive episode without psychotic symptoms |
| E118. | seasonal affective disorder | X761L | seasonal affective disorder |
| E1130 | recurrent major depressive episodes unspecified | E1130 | recurrent major depressive episodes unspecified |
| E1133 | recurrent major depressive episodes severe without mention of psychosis | E1133 | recurrent major depressive episodes severe without mention of psychosis |
| Eu32A | recurrent major depressive episodes severe with psychosis in remission | XaX54 | recurrent major depressive episodes severe with psychosis in remission |
| E1134 | recurrent major depressive episodes severe with psychosis | E1134 | recurrent major depressive episodes severe with psychosis |
| E1132 | recurrent major depressive episodes moderate | E1132 | recurrent major depressive episodes moderate |
| E1131 | recurrent major depressive episodes mild | E1131 | recurrent major depressive episodes mild |
| E1135 | recurrent major depressive episodes in partial or unspecified remission | E1135 | recurrent major depressive episodes in partial or unspecified remission |
| E1136 | recurrent major depressive episodes in full remission | E1136 | recurrent major depressive episodes in full remission |
| E113. | recurrent major depressive episode | XE1Y1 | recurrent major depressive episodes |
| E113z | recurrent major depressive episode nos | E113z | recurrent major depressive episode nos |
| Eu33z | recurrent depressive disorder unspecified | XE1Zf | recurrent depressive disorder unspecified |
| Eu334 | recurrent depressive disorder currently in remission | Eu334 | recurrent depressive disorder currently in remission |
| Eu332 | recurrent depressive disorder current episode severe without psychotic symptoms | XE1Zd | recurrent depressive disorder current episode severe without psychotic symptoms |

| Read v2 | Term v2 | CTV3 | Term-CTV3 |
|---|---|---|---|
| Eu333 | recurrent depressive disorder current episode severe with psychotic symptoms | XE1Ze | recurrent depressive disorder current episode severe with psychotic symptoms |
| Eu331 | recurrent depressive disorder current episode moderate | Eu331 | recurrent depressive disorder current episode moderate |
| Eu330 | recurrent depressive disorder current episode mild | Eu330 | recurrent depressive disorder current episode mild |
| Eu33. | recurrent depressive disorder | XE1Zc | recurrent depressive disorder |
| E1137 | recurrent depression | E1137 | recurrent depression |
| E291. | prolonged depressive reaction | E291. | prolonged depressive adjustment reaction |
| Eu33y | other recurrent depressive disorders | Eu33y | other recurrent depressive disorders |
| Eu32y | other depressive episodes | XE1Za | other depressive episodes |
| Eu326 | major depression moderately severe | XSGol | moderate major depression |
| Eu321 | moderate depressive episode | Eu321 | moderate depressive episode |
| Eu412 | mixed anxiety and depressive disorder | X00Sb | mixed anxiety and depressive disorder |
| Eu325 | major depression mild | XSGok | mild major depression |
| Eu320 | mild depressive episode | Eu320 | mild depressive episode |
| Eu324 | mild depression | XaCIs | mild depression |
| E11z2 | masked depression | X00SU | masked depression |
| Eu341 | dysthymia | E2112 | dysthymia |
| Eu32z | depressive episode unspecified | XE1Zb | depressive episode unspecified |
| E2B.. | depressive disorder nec | E2B.. | depressive disorder nec |
| E2B1. | chronic depression | E2B1. | chronic depression |
| E290z | brief depressive reaction nos | E290z | brief depressive reaction nos |
| E11y2 | atypical depressive disorder | E11y2 | atypical depressive disorder |
| Eu32B | antenatal depression | XaY2C | antenatal depression |
| E135. | agitated depression | X00SQ | agitated depression |
| - | - | E112. | depression single major episode or agitated or endogenous including first episode |
| - | - | E1120 | single major depressive episode unspecified |
| - | - | E1121 | single major depressive episode mild |
| - | - | E1122 | single major depressive episode moderate |
| - | - | E1123 | single major depressive episode severe without mention of psychosis |
| - | - | E1124 | single major depressive episode severe with psychosis |
| - | - | E1125 | single major depressive episode in partial or unspecified remission |
| - | - | E1126 | single major depressive episode in full remission |
| - | - | E112z | single major depressive episode nos |
| - | - | E113. | recurrent depression major episode or endogenous |
| - | - | E1130 | recurrent major depressive episodes unspecified |
| - | - | E1131 | recurrent major depressive episodes mild |
| - | - | E1132 | recurrent major depressive episodes moderate |

| Read v2 | Term v2 | CTV3 | Term-CTV3 |
|---|---|---|---|
| - | - | E1133 | recurrent major depressive episodes severe without mention of psychosis |
| - | - | E1134 | recurrent major depressive episodes severe with psychosis |
| - | - | E1135 | recurrent major depressive episodes in partial or unspecified remission |
| - | - | E1136 | recurrent major depressive episodes in full remission |
| - | - | E1137 | recurrent depression |
| - | - | E113z | recurrent major depressive episode nos |
| - | - | E11z2 | masked depression |
| - | - | E135. | agitated depression |
| - | - | E2003 | mixed anxiety and depressive disorder |
| - | - | E204. | neurotic depression reactive type or postnatal depression |
| - | - | E2112 | dysthymia |
| - | - | E290z | brief depressive reaction nos |
| - | - | E291. | prolonged depressive adjustment reaction |
| - | - | E2B.. | depressive disorder nec |
| - | - | E2B1. | chronic depression |
| - | - | Eu320 | mild depressive episode |
| - | - | Eu321 | moderate depressive episode |
| - | - | Eu322 | severe depressive episode without psychotic symptoms single episode agitated depression or single episode major depression or single episode vital depression |
| - | - | Eu323 | severe depressive episode with psychotic symptoms single episode of major depression or psychogenic depressive psychosis or psychotic depression or reactive depressive psychosis |
| - | - | Eu324 | mild depression |
| - | - | Eu325 | mild major depression |
| - | - | Eu326 | moderate major depression |
| - | - | Eu32y | depression other episodes or atypical or single episode masked nos |
| - | - | Eu32z | depression episode unspecified or nos reactive or depressive disorder nos |
| - | - | Eu33. | recurrent depressive disorder episodes of depressive reaction or episodes of psychogenic depression or episodes of reactive depression or seasonal depressive disorder |
| - | - | Eu330 | recurrent depressive disorder current episode mild |
| - | - | Eu331 | recurrent depressive disorder current episode moderate |

| Read v2 | Term v2 | CTV3 | Term-CTV3 |
|---|---|---|---|
| - | - | Eu332 | depression without psychotic symptoms recurrent major or manic depressive psychosis depressed type or vital or current severe episode or endogenous |
| - | - | Eu333 | depression with psychotic symptoms recurrent current episode severe or manic depressive depressed type or severe major episodes or severe episodes or endogenous |
| - | - | Eu334 | recurrent depressive disorder currently in remission |
| - | - | Eu33y | other recurrent depressive disorders |
| - | - | Eu33z | depression recurrent unspecified or monopolar nos |
| - | - | Eu34. | persistent mood affective disorders |
| - | - | Eu3y1 | recurrent mood affective disorders brief episodes or other |
| - | - | Eu412 | mixed anxiety and depressive disorder mild anxiety depression |
| - | - | X00Sb | mixed anxiety and depressive disorder |
| - | - | X00SO | depressive disorder |
| - | - | X00SQ | agitated depression |
| - | - | X00SR | endogenous depression |
| - | - | X00SS | endogenous depression first episode |
| - | - | X00SU | masked depression |
| - | - | X761L | seasonal affective disorder |
| - | - | Xa0wV | recurrent brief depressive disorder |
| - | - | XaB9J | depression nos |
| - | - | XaCHr | single episode agitated depression without psychotic symptoms |
| - | - | XaCHs | single episode major depression without psychotic symptoms |
| - | - | XaCIs | mild depression |
| - | - | XaCIu | severe depression |
| - | - | XaX53 | single major depressive episode severe with psychosis in remission |
| - | - | XaX54 | recurrent major depressive episodes severe with psychosis in remission |
| - | - | XaY2C | antenatal depression |
| - | - | XE1aY | depression reactive neurotic or postnatal |
| - | - | XE1Y0 | single major depressive episode |
| - | - | XE1Y1 | recurrent major depressive episodes |
| - | - | XE1Za | other depressive episodes |
| - | - | XE1Zb | depressive episode unspecified |
| - | - | XE1Zc | recurrent depressive disorder |
| - | - | XE1Zd | recurrent depressive disorder current episode severe without psychotic symptoms |

| Read v2 | Term v2 | CTV3 | Term-CTV3 |
|---|---|---|---|
| - | - | XE1Ze | recurrent depressive disorder current episode severe with psychotic symptoms |
| - | - | XE1Zf | recurrent depressive disorder unspecified |
| - | - | XE1ZY | severe depressive episode without psychotic symptoms |
| - | - | XM1GC | endogenous depression recurrent |
| - | - | XSEGJ | major depressive disorder |
| - | - | XSGok | mild major depression |
| - | - | XSGol | moderate major depression |
| - | - | XSGom | severe major depression without psychotic features |
| - | - | XSGon | severe major depression with psychotic features |
| - | - | XE1YC | reactive depression |
| - | - | XE1ZZ | severe depressive episode with psychotic symptoms |
| - | - | XE1Xy | mood disorder |

Table 4 International Classification of Diseases codes for depression and anxiety

| Phenotype | ICD10 code | Description |
|---|---|---|
| **Depression** | F32 | Depressive episodes |
| | | F32.0 Mild depressive episode |
| | | F32.1 Moderate depressive episode |
| | | F32.2 Severe depressive episode without psychotic symptoms |
| | | F32.3 Severe depressive episode with psychotic symptoms |
| | | F32.8 Other depressive episodes |
| | | F32.9 Depressive episode, unspecified |
| | F33 | Recurrent depressive episode |
| | | F33.0 Recurrent depressive disorder, current episode mild |
| | | F33.1 Recurrent depressive disorder, current episode moderate |
| | | F33.2 Recurrent depressive disorder, current episode severe without psychotic symptoms |
| | | F33.3 Recurrent depressive disorder, current episode severe with psychotic symptoms |
| | | F33.4 Recurrent depressive disorder, currently in remission |
| | | F33.8 Other recurrent depressive disorders |
| | | F33.9 Recurrent depressive disorder, unspecified |
| | F34 | Persistent mood (affective disorders) |
| | | F34.0 Cyclothymia |
| | | F34.1 Dysthymia |
| | | F34.8 Other persistent mood [affective] disorders |
| | | F34.9 Persistent mood [affective] disorder, unspecified |
| | F38 | Other mood (affective) disorders |
| | | F38.0 Other single mood [affective] disorders |
| | | F38.1 Other recurrent mood [affective] disorders |
| | | F38.8 Other specified mood [affective] disorders |
| | F39 | Unspecified mood (affective) disorders |
| **Anxiety** | F40 | Phobic anxiety disorders |
| | | F40.0 Agoraphobia |
| | | F40.1 Social phobias |
| | | F40.2 Specific (isolated) phobias |
| | | F40.8 Other phobic anxiety disorders |
| | | F40.9 Phobic anxiety disorder, unspecified |
| | F41 | Other anxiety disorders |
| | | F41.0 Panic disorder [episodic paroxysmal anxiety] |
| | | F41.1 Generalised anxiety disorder |
| | | F41.2 Mixed anxiety and depressive disorder |
| | | F41.3 Other mixed anxiety disorders |
| | | F41.8 Other specified anxiety disorders |
| | | F41.9 Anxiety disorder, unspecified |

*ICD-10 International Classification of Diseases, 10$^{th}$ Revision*

Table 5 Primary care Read codes for anxiety

| Read v2 | Term v2 | CTV3 | Term-CTV3 |
|---|---|---|---|
| 146G. | H/O: agoraphobia | 146G. | H/O: agoraphobia |
| 1Bb.. | Specific fear | E2022 | Agoraphobia without mention of panic attacks |
| 1Bb0. | Fear of falling | Eu400 | [X] Agoraphobia (& [without history of panic disorder] or [with panic disorder]) |
| 1Bb1. | Fear of getting cancer | X00SV | Agoraphobia |
| E200. | Anxiety states | X761n | Fear of going out |
| E2000 | Anxiety state unspecified | X761R | Situation avoidance behaviour |
| E2001 | Panic disorder | XaK2c | H/O: agoraphobia |
| E2002 | Generalised anxiety disorder | E200. | Anxiety disorder |
| E2004 | Chronic anxiety | E2000 | Anxiety state unspecified |
| E2005 | Recurrent anxiety | E200z | Anxiety state NOS |
| E200z | Anxiety state NOS | XE1aW | (Anxiety state (& [states] or [panic attack])) or (pseudocyesis) |
| E202. | Phobic disorders | E2004 | Chronic anxiety |
| E2020 | Phobia unspecified | E2002 | Generalised anxiety disorder |
| E2021 | Agoraphobia with panic attacks | X761T | Anxiety about body function or health |
| E2022 | Agoraphobia without mention of panic attacks | X762I | Fear associated with illness and body function |
| E2023 | Social phobia, fear of eating in public | Eu41. | [X]Other anxiety disorders |
| E2024 | Social phobia, fear of public speaking | Eu411 | Anxiety neurosis |
| E2025 | Social phobia, fear of public washing | Eu413 | [X]Other mixed anxiety disorders |
| E2026 | Acrophobia | Eu41y | [X] Anxiety disorders: [other specified] or [anxiety hysteria] |
| E2027 | Animal phobia | Eu41z | [X]Anxiety disorder, unspecified |
| E2028 | Claustrophobia | XE1Zj | [X]Other specified anxiety disorders |
| E2029 | Fear of crowds | E2001 | (Panic disorder) or (panic attack) |
| E202A | Fear of flying | Eu410 | [X]Panic disorder [episodic paroxysmal anxiety] |
| E202B | Cancer phobia | XE1Y7 | Panic disorder |
| E202C | Dental phobia | E2021 | Agoraphobia with panic attacks |
| E202D | Fear of death | E202. | Phobic disorders (& [social] or [phobic anxiety]) |
| E202E | Fear of pregnancy | E2020 | Phobia unspecified |
| E202z | Phobic disorder NOS | E2026 | Acrophobia |
| Eu40. | [X]Phobic anxiety disorders | E2027 | Animal phobia |
| Eu400 | [X]Agoraphobia | E2028 | Claustrophobia |
| Eu401 | [X]Social phobias | E2029 | Fear of crowds |
| Eu402 | [X]Specific (isolated) phobias | E202A | Fear of flying |
| Eu403 | [X]Needle phobia | E202B | Cancer phobia |
| Eu40y | [X]Other phobic anxiety disorders | E202C | Dental phobia |
| Eu40z | [X]Phobic anxiety disorder, unspecified | E202D | Fear of death |
| Eu41. | [X]Other anxiety disorders | E202E | Fear of pregnancy |
| Eu410 | [X]Panic disorder [episodic paroxysmal anxiety] | E202z | (Weight fixation) or (phobic disorder NOS) |
| Eu411 | [X]Generalized anxiety disorder | E28z. | (Examination fear) or (flying phobia) or (stage fright) or (acute stress reaction NOS) |
| Eu413 | [X]Other mixed anxiety disorders | Eu40. | [X]Phobic anxiety disorders |
| Eu41y | [X]Other specified anxiety disorders | Eu402 | [X] Specific (isolated) phobias (& [acrophobia] or [animal] or [claustrophobia] or [simple phobia]) |
| Eu41z | [X]Anxiety disorder, unspecified | Eu403 | Needle phobia |
| - | - | Eu40z | [X]Phobic anxiety disorder, unspecified |
| - | - | Ua1qa | Fear of dentist |
| - | - | Ua1qc | Fear of not coping with treatment |
| - | - | Ua1qd | Fear of lifts |
| - | - | Ua1qe | Fear of thunderstorm |
| - | - | Ua1qS | Panic attack |
| - | - | Ua1qs | Fear of wetting self in public |
| - | - | Ua1qt | Fear of losing control of bowels in public |
| - | - | Ua1qU | Fear of walking |
| - | - | Ua1qV | Fear of mobilising |
| - | - | Ua1qW | Fear of disconnection from ventilator |
| - | - | Ua1qX | Fear of being left alone during period of dependence |
| - | - | Ua1qY | Fear of being left alone |
| - | - | X00Sr | Erythrophobia |
| - | - | X00SX | Specific phobia |
| - | - | X00SY | Needle phobia |

| Read v2 | Term v2 | CTV3 | Term-CTV3 |
|---|---|---|---|
| - | - | X00Sy | Weight fixation |
| - | - | X50G2 | Trichophobia |
| - | - | X50G3 | Parasitophobia |
| - | - | X50G5 | Venereophobia |
| - | - | X50G6 | Syphilophobia |
| - | - | X50GI | Bromisodrophobia |
| - | - | X761a | Fear of swallowing |
| - | - | X761b | Fear of collapsing |
| - | - | X761c | Fear of fainting |
| - | - | X761d | Fear of having a heart attack |
| - | - | X761e | Fear of shaking |
| - | - | X761f | Fear of sweating |
| - | - | X761G | Loss of hope for the future |
| - | - | X761g | Fear of dying |
| - | - | X761h | Fear of going crazy |
| - | - | X761i | Fear of losing emotional control |
| - | - | X761j | Fear of becoming fat |
| - | - | X761p | Fear of empty streets |
| - | - | X761q | Fear of open spaces |
| - | - | X761r | Fear of crossing streets |
| - | - | X761t | Fear of transport |
| - | - | X761U | Fear of losing control of bowels |
| - | - | X761V | Fear of wetting self |
| - | - | X761X | Fear of having a fit |
| - | - | X761y | Fear of using public toilets |
| - | - | X761Y | Fear of choking |
| - | - | X761Z | Anxiety about blushing |
| - | - | X7621 | Fear of being in a small group |
| - | - | X7623 | Fear of speaking on the phone |
| - | - | X7627 | Specific fear |
| - | - | X7628 | Fear of natural phenomena |
| - | - | X7629 | Fear of the dark |
| - | - | X762a | Fear of ghosts |
| - | - | X762A | Fear of animals |
| - | - | X762b | Fear of school |
| - | - | X762B | Fear of feathers |
| - | - | X762C | Fear of enclosed spaces |
| - | - | X762E | Fear of tunnels |
| - | - | X762F | Fear of phone boxes |
| - | - | X762G | Fear of flying |
| - | - | X762H | Flying phobia |
| - | - | X762J | Fear of anaesthetic |
| - | - | X762K | Fear of general anaesthetic |
| - | - | X762L | Fear of awareness under general anaesthetic |
| - | - | X762M | Fear of not waking from general anaesthetic |
| - | - | X762N | Fear of local anaesthetic |
| - | - | X762O | Fear of problem after anaesthetic |
| - | - | X762P | Fear of needles |
| - | - | X762R | Fear of surgical masks |
| - | - | X762S | Fear of hospitals |
| - | - | X762T | Fear of death |
| - | - | X762U | Fear of contracting disease |
| - | - | X762V | Fear of infection |
| - | - | X762W | Fear of contracting venereal disease |
| - | - | X762X | Fear of contracting HIV infection |
| - | - | X762Y | Fear of contracting radiation sickness |
| - | - | X762Z | Fear of the bogey man |
| - | - | Xa00r | Fear of heights |
| - | - | Xa00s | Fear of water |
| - | - | Xa1a8 | Examination phobia |
| - | - | Xa1Ev | Fear of needles |
| - | - | Xa3Vk | Fear of insects |
| - | - | Xa3Vl | Fear of birds |

| Read v2 | Term v2 | CTV3 | Term-CTV3 |
|---|---|---|---|
| - | - | Xa3WI | Fear of blood |
| - | - | Xa3WJ | Fear of getting cancer |
| - | - | Xa7lj | Cancer phobia |
| - | - | XaB96 | Other phobias |
| - | - | XaEKL | Phonophobia |
| - | - | XaIvf | Fear of falling |
| - | - | XE1YA | Phobic anxiety disorder |
| - | - | XE1YB | Phobic disorder NOS |
| - | - | E2005 | Recurrent anxiety |
| - | - | E2023 | Social phobia, fear of eating in public |
| - | - | E2024 | Social phobia, fear of public speaking |
| - | - | E2025 | Social phobia, fear of public washing |
| - | - | Eu401 | Social phobia |
| - | - | X00SW | Social phobia |
| - | - | X761k | Anxiety about behaviour or performance |
| - | - | X761l | Fear of appearing ridiculous |
| - | - | X761m | Fear of saying the wrong thing |
| - | - | X761u | Social fear |
| - | - | X761v | Fear of activities in public |
| - | - | X761w | Fear of eating in public |
| - | - | X761W | Fear of vomiting in public |
| - | - | X761x | Fear of public speaking |
| - | - | X761z | Fear of writing in public |
| - | - | X7620 | Fear of social group activities |
| - | - | X7622 | Fear of social gatherings |
| - | - | X7624 | Fear of speaking to people in authority |
| - | - | X7625 | Fear of being laughed at |
| - | - | X7626 | Fear of being watched |

Table 6 Description and coding of confounders

| Measure | Field code | Description and re-coding | Final variable coding |
|---|---|---|---|
| *Sociodemographic variables* | | | |
| Ethnicity | 21000 | Ethnicity at baseline:<br>▪ "Prefer not to answer", "Do not know" = NA (missing)<br>▪ "White", "British", "Irish", "Any other white background" = white (1)<br>▪ All other ethnicities = non-white (2) | White (1) vs. Non-white (2) |
| Socio-economic status (TDI Townsend deprivation index) | 189 | Townsend deprivation score at baseline:<br>Reflects information on social class, housing, employment, and car availability | Continuous score |
| Education | 6138 | Highest educational attainment at baseline:<br>▪ "Prefer not to answer" = NA (missing)<br>▪ "College or University degree", "Other professional qualifications eg: nursing, teaching" = degree (or professional) (2)<br>▪ "None of the above", "A levels/AS levels or equivalent", "O levels/GCSEs or equivalent", "CSEs or equivalent", "NVQ or HND or HNC or equivalent", = other (1) | Other (1) vs. Degree (2) |
| Cohabit with a partner or spouse | 6141 | "How are the people in household related to participant" at baseline:<br>▪ "Prefer not to answer", "Do not know" = NA (missing)<br>▪ "Husband, wife or partner" = cohabit partner (1)<br>▪ All other options and NAs = Other (2)<br>*Note: question only completed if answering >1 to Field 709 "Including yourself how many people live in your household"*<br>*"Other" includes no cohabit or cohabit with those other than partner or spouse* | Cohabit partner (1) vs. Other (2) |
| *Lifestyle and health variables* | | | |
| Body mass index (BMI) | 21001 | BMI measured at baseline visit:<br>Height and weight measured in clinic. BMI calculated as weight (kg) / height (m)$^2$ | Continuous variable |
| Smoking status | 20116 | Smoking status self-reported at baseline:<br>"Prefer not to answer" = NA (missing)<br>▪ "Never" (1) | Never (1) vs. Previous (2) vs. Current (3) |

| Measure | Field code | Description and re-coding | Final variable coding |
|---|---|---|---|
| | | ▪ "Previous" (2) | |
| | | ▪ "Current" (3) | |
| Alcohol status | 1558 | Frequency of alcohol intake reported at baseline: | Never (1) vs. Infrequent (2) vs. Frequent (3) |
| | | ▪ "Prefer not to answer" = NA (missing) | |
| | | ▪ "never" = never (1) | |
| | | ▪ "One to three times a month", "Special occasions only" = infrequent (2) | |
| | | ▪ "Daily or almost daily", "Three or four times a week", "Once or twice a week" = frequent (3) | |
| Multimorbidity | 20002 | Non-cancer illnesses self-reported at baseline (ignoring depression, anxiety) | No (1) vs. Multimorbidity (2) |
| | | See the 36 most common illnesses from https://bmcmedicine.biomedcentral.com/articles/10.1186/s12916-019-1339-0#Sec2 Illnesses excluding anxiety, depression, prostate problems, and schizophrenia | |
| | | ▪ <2 = none_ or_one, | |
| | | ▪ 2 or more = multi morbidity | |
| *Reproductive variables* | | | |
| Menopause status | 2774 | Menopause status self-reported at baseline: | No (1) vs. Do not know (2) vs. Yes (3) |
| | | ▪ "Prefer not to answer" = NA (missing) | |
| | | ▪ "No" = no (1) | |
| | | ▪ *"Not sure - had a hysterectomy", "Not sure – other reason" = Do not know (2)* | |
| | | ▪ "Yes" = yes (3) | |
| Hysterectomy (derived variable) | 3591 & 2774 | Hysterectomy reported at baseline: | No (1) vs. Yes (2) |
| | | ▪ "Prefer not to answer", "Not sure" = NA (missing) | |
| | | ▪ "No" = No (1) | |
| | | ▪ "Yes" & ["Not sure - had a hysterectomy" in Field 2774] = yes (2) | |
| Hormone replacement therapy (HRT) | 2814 | Self-reported HRT use at baseline: | No (1) vs. Yes (2) |
| | | ▪ "Prefer not to answer", "Do not know" = NA (missing) | |

| Measure | Field code | Description and re-coding | Final variable coding |
|---|---|---|---|
| | | ▪ "No" = no (1) | |
| | | ▪ "Yes" = yes (2) | |
| Parity | 2734 | Self-reported parity at baseline: | None (1) vs. 1 (2) vs. 2+ (3) |
| | | Continuous measure (0-22) | |
| | | Recode: | |
| | | ▪ "0" = none (1) | |
| | | ▪ "1" = one (2) | |
| | | ▪ "2-22" = 2+ (3) | |

Table 7 Information about the publicly available GWAS summary statistics for depression, anxiety, and neuroticism

| GWAS | Population | Measurement | Link to summary data | Comment |
|---|---|---|---|---|
| **Depression:**<br>MDD working group of PGC 2025<br>PMID: 39814019 | 3.9 Million males and females of European ancestry.<br>*Note: Trans-ancestry (29 countries); individual summary results for Europeans available.* | Case/Control lifetime diagnosis of MDD measured via clinical, electronic health records, and questionnaires. | https://figshare.com/articles/dataset/GWAS_summary_statistics_for_major_depression_PGC_MDD2025_/27061255 | **Summary data are available sans UK Biobank** (and 23andMe) participants, allowing for *known* sample overlap to be reduced to zero.<br>N-cases/N-controls for the European population (sans UKBB and 23andMe):<br>357,636/1,281,936<br>**Ntotal 1,639,572** |
| **Anxiety:**<br>Purves *et al.* 2019<br>PMCID: PMC7237282 | 83, 566 males and females of European ancestry from UK Biobank. | Lifetime Anxiety Disorder: either self-report lifetime professional diagnosis of one or more core anxiety disorders or meeting criteria for a likely lifetime diagnosis of DSM-IV generalised anxiety disorder based on questions from the Composite International Diagnostic Interview (CIDI) Short-form questionnaire. | https://drive.google.com/drive/folders/1fguHvz7l2G45sbMI9h_veQun4aXNTy1v | **83,566** (25,453 cases and 58,113 controls)<br>**Note that this sample comprises entirely of UK Biobank participants.** However, the sample includes a subset of UKBB (~37%) who participated in the MHQ. This includes both males and females (57%) and participants were not restricted to those with EHR data.<br>57% were female giving approximately 48,772 women.<br>**This means the maximum overlap proportion with the AUI GWAS is 41% however**, given both the MHQ and EHR samples are independent subsets of participants the proportion of sample overlap is likely much lower. |
| **Neuroticism:**<br>Gupta *et al.* 2024<br>PMID: 39134740 | 449,484 males and females of European ancestry<br>*Note: Trans-ancestry (European and African ancestry); UK Biobank and the Million Veteran Program cohort. Individual* | Continuous score of neuroticism captured via questionnaire data (BFI-10). | https://medicine.yale.edu/lab/leveylab/data/ | **Summary results are not available sans UK Biobank,** therefore there is some sample overlap: |

| GWAS | Population | Measurement | Link to summary data | Comment |
|---|---|---|---|---|
| | *summary results for Europeans available.* | | | There are 449,484 UKBB participants in the **total sample of 682,688.**<br>The UKBB AUI GWAS has a total sample N of 118,783, therefore the **possible maximum overlap could be up to 17.4% of the total sample.** |

Table 8 Number of SNPs for genetic instruments for the exposures in MR

| Exposure GWAS | Outcome GWAS | Number of SNPs P<1E-8 (exposure) | Number of SNPs after clumping | Number of exposure SNPs in outcome GWAS | Number of exposure SNPs missing | Number of identified proxy SNPs | Total number of available SNPs as exposure | Total SNPs remaining following harmonisation | N SNPs using relaxed threshold (P<5E-6 in exposure) |
|---|---|---|---|---|---|---|---|---|---|
| Depression | AUI | 9389 | 147 | 146 | 1 | 0 | 146 | 146 | NA |
| | SUI | | | | | | | | |
| | UUI | | | | | | | | |
| | MUI | | | | | | | | |
| Anxiety | AUI | 432 | 5 | 5 | 0 | NA | 5 | 5 | 31/35 |
| | SUI | | | | | | | | |
| | UUI | | | | | | | | |
| | MUI | | | | | | | | |
| Neuroticism | AUI | 13039 | 132 | 132 | 0 | NA | 132 | 130 | NA |
| | SUI | | | | | | | | |
| | UUI | | | | | | | | |
| | MUI | | | | | | | | |
| AUI | Depression | 73 | 3 | 3 | 0 | NA | 3 | 3 | 28/37 |
| | Anxiety | | | | | | | | 32/37 |
| | Neuroticism | | | | | | | | 34/37 |
| SUI | Depression | 224 | 6 | 5 | 1 | 0 | 5 | 5 | 33/39 |
| | Anxiety | | | | | | | | 36/39 |
| | Neuroticism | | | | | | | | 35/39 |
| UUI | Depression | 0 | NA | NA | NA | NA | NA | NA | 9/10 |
| | Anxiety | | | | | | | | 10/10 |
| | Neuroticism | | | | | | | | 10/10 |
| MUI | Depression | 3 | 1 | 1 | 0 | NA | 1 | 1 | 16/19 |
| | Anxiety | | | | | | | | 19/19 |
| | Neuroticism | | | | | | | | 16/19 |

*GWAS genome-wide association study; AUI any urinary incontinence (UI); SUI stress UI; UUI urge UI; MUI mixed UI; SNP single nucleotide polymorphism; N number; NA not applicable*

Table 9 Prevalence of UI and MH phenotypes across data samples

| **All data: N HES =273,158 ; N EHR & Combined = 122,216** | | | | | | | | | |
|---|---|---|---|---|---|---|---|---|---|
| Prevalence: N (%) | Cases in EHR | Non-cases in EHR | Prevalence EHR (%) | Cases in HES | Non-cases in HES | Prevalence HES (%) | Cases in combined sample | Non-cases in combined sample | Prevalence combined(%) |
| AUI | 11141 | 111075 | 9.12 | 9825 | 263333 | 3.60 | 12978 | 109238 | 10.62 |
| SUI | 6946 | 115270 | 5.68 | 8362 | 264796 | 3.06 | 8885 | 113331 | 7.27 |
| UUI | 2266 | 119950 | 1.85 | 2524 | 270634 | 0.92 | 3143 | 119073 | 2.57 |
| MUI | 610 | 121606 | 0.50 | 1061 | 272097 | 0.39 | 1017 | 121199 | 0.83 |
| Any urgency | 4188 | 118028 | 3.43 | NA | NA | NA | 4188 | 118028 | 3.43 |
| Depression | 25384 | 96832 | 20.77 | 21345 | 251813 | 7.81 | 28700 | 93516 | 23.48 |
| Anxiety | 15065 | 107151 | 12.33 | 18237 | 254921 | 6.68 | 20147 | 102069 | 16.48 |
| **Complete case data for confounders: N HES =262,961 ; N EHR & Combined = 118,526** | | | | | | | | | |
| Prevalence: N (%) | Cases in EHR | Non-cases in EHR | Prevalence EHR (%) | Cases in HES | Non-cases in HES | Prevalence HES (%) | Cases in combined sample | Non-cases in combined sample | Prevalence combined(%) |
| AUI | 10725 | 107801 | 9.05 | 9343 | 253618 | 3.55 | 12473 | 106053 | 10.52 |
| SUI | 6700 | 111826 | 5.65 | 7965 | 254996 | 3.03 | 8554 | 109972 | 7.22 |
| UUI | 2179 | 116347 | 1.84 | 2383 | 260578 | 0.91 | 3013 | 115513 | 2.54 |
| MUI | 579 | 117947 | 0.49 | 1005 | 261956 | 0.38 | 970 | 117556 | 0.82 |
| Any urgency | 4028 | 114498 | 3.40 | NA | NA | NA | 4028 | 114498 | 3.40 |
| Depression | 24530 | 93996 | 20.70 | 20218 | 242743 | 7.69 | 27701 | 90825 | 23.37 |
| Anxiety | 14574 | 103952 | 12.30 | 17322 | 245639 | 6.59 | 19458 | 99068 | 16.42 |

*AUI any urinary incontinence (UI); SUI stress UI; UUI urge UI; MUI mixed UI; EHR Electronic Health Records (primary care records); HES Hospital Episode Statistics (hospital inpatient records); N number; NA not applicable*

Table 10 Evaluation of the proportional hazards' assumption using scaled Schoenfeld residuals

| Outcome | Exposure/Confounder variable | df | Exposure: Depression | | | Exposure: Anxiety | | | Exposure: Neuroticism | | |
|---|---|---|---|---|---|---|---|---|---|---|---|
| | | | $\chi^2$ | P value | PH Violated? | $\chi^2$ | P value | PH Violated? | $\chi^2$ | P value | PH Violated? |
| Any UI | Prevalent exposure | 1 | 7.62E-04 | 0.98 | No | 0.15 | 0.70 | No | 0.02 | 0.88 | No |
| | Age at recruitment | 1 | 13.81 | <0.001 | Yes | 13.64 | <0.001 | Yes | 7.25 | 0.01 | Yes |
| | Ethnicity | 1 | 4.57 | 0.03 | Yes | 4.48 | 0.03 | Yes | 1.19 | 0.28 | No |
| | TDI | 1 | 0.99 | 0.32 | No | 0.94 | 0.33 | No | 0.56 | 0.45 | No |
| | Cohabit with partner | 1 | 4.82 | 0.03 | Yes | 4.80 | 0.03 | Yes | 2.38 | 0.12 | No |
| | Education | 1 | 0.98 | 0.32 | No | 0.95 | 0.33 | No | 0.03 | 0.86 | No |
| | Alcohol | 2 | 0.25 | 0.88 | No | 0.25 | 0.88 | No | 0.18 | 0.91 | No |
| | Smoking | 2 | 2.42 | 0.30 | No | 2.43 | 0.30 | No | 1.06 | 0.59 | No |
| | Multi-morbidity | 1 | 0.78 | 0.38 | No | 0.80 | 0.37 | No | 0.94 | 0.33 | No |
| | BMI | 1 | 0.25 | 0.62 | No | 0.26 | 0.61 | No | 0.52 | 0.47 | No |
| | Menopause | 3 | 11.84 | 0.01 | Yes | 11.75 | 0.01 | Yes | 16.49 | <0.001 | Yes |
| | HRT | 1 | 0.21 | 0.65 | No | 0.24 | 0.63 | No | 0.18 | 0.67 | No |
| | Hysterectomy | 1 | 1.23 | 0.27 | No | 1.24 | 0.27 | No | 3.23 | 0.07 | No |
| | Parity | 2 | 2.65 | 0.27 | No | 2.66 | 0.27 | No | 2.37 | 0.31 | No |
| | **GLOBAL** | **19** | **45.55** | **<0.001** | **Yes** | **45.25** | **<0.001** | **Yes** | **31.91** | **0.03** | **Yes** |
| Stress UI | Prevalent exposure | 1 | 0.25 | 0.62 | No | 0.08 | 0.78 | No | 0.56 | 0.46 | No |
| | Age at recruitment | 1 | 0.42 | 0.52 | No | 0.40 | 0.53 | No | 0.24 | 0.63 | No |
| | Ethnicity | 1 | 1.71 | 0.19 | No | 1.70 | 0.19 | No | 0.19 | 0.67 | No |
| | TDI | 1 | 0.12 | 0.73 | No | 0.11 | 0.74 | No | 4.17E-03 | 0.95 | No |
| | Cohabit with partner | 1 | 2.37 | 0.12 | No | 2.37 | 0.12 | No | 0.83 | 0.36 | No |
| | Education | 1 | 0.95 | 0.33 | No | 0.94 | 0.33 | No | 0.46 | 0.50 | No |
| | Alcohol | 2 | 0.01 | 1.00 | No | 4.99E-03 | 1.00 | No | 0.20 | 0.90 | No |
| | Smoking | 2 | 1.71 | 0.43 | No | 1.70 | 0.43 | No | 0.49 | 0.78 | No |
| | Multi-morbidity | 1 | 1.09 | 0.30 | No | 1.09 | 0.30 | No | 1.70 | 0.19 | No |
| | BMI | 1 | 0.44 | 0.51 | No | 0.43 | 0.51 | No | 0.33 | 0.56 | No |
| | Menopause | 3 | 3.84 | 0.28 | No | 3.80 | 0.28 | No | 10.10 | 0.02 | Yes |
| | HRT | 1 | 1.87 | 0.17 | No | 1.84 | 0.18 | No | 2.96 | 0.09 | No |
| | Hysterectomy | 1 | 1.44 | 0.23 | No | 1.44 | 0.23 | No | 1.55 | 0.21 | No |
| | Parity | 2 | 2.21 | 0.33 | No | 2.22 | 0.33 | No | 3.99 | 0.14 | No |
| | **GLOBAL** | **19** | **19.87** | **0.40** | **No** | **19.48** | **0.43** | **No** | **25.40** | **0.15** | **No** |
| Urge UI | Prevalent exposure | 1 | 0.06 | 0.81 | No | 4.41 | 0.04 | Yes | 1.08 | 0.30 | No |

| | | | | | | | | | | | |
|---|---|---|---|---|---|---|---|---|---|---|---|
| | Age at recruitment | 1 | 1.51 | 0.22 | No | 1.47 | 0.23 | No | 0.03 | 0.86 | No |
| | Ethnicity | 1 | 0.78 | 0.38 | No | 0.75 | 0.39 | No | 0.01 | 0.91 | No |
| | TDI | 1 | 5.38 | 0.02 | Yes | 5.29 | 0.02 | Yes | 2.94 | 0.09 | No |
| | Cohabit with partner | 1 | 0.24 | 0.63 | No | 0.23 | 0.63 | No | 1.08 | 0.30 | No |
| | Education | 1 | 0.75 | 0.39 | No | 0.76 | 0.38 | No | 2.91 | 0.09 | No |
| | Alcohol | 2 | 1.99 | 0.37 | No | 1.99 | 0.37 | No | 1.19 | 0.55 | No |
| | Smoking | 2 | 1.20 | 0.55 | No | 1.20 | 0.55 | No | 0.70 | 0.70 | No |
| | Multi-morbidity | 1 | 1.04 | 0.31 | No | 1.07 | 0.30 | No | 1.24 | 0.27 | No |
| | BMI | 1 | 0.59 | 0.44 | No | 0.63 | 0.43 | No | 0.67 | 0.41 | No |
| | Menopause | 3 | 3.76 | 0.29 | No | 3.73 | 0.29 | No | 3.59 | 0.31 | No |
| | HRT | 1 | 0.20 | 0.65 | No | 0.22 | 0.64 | No | 0.76 | 0.38 | No |
| | Hysterectomy | 1 | 0.54 | 0.46 | No | 0.56 | 0.46 | No | 0.09 | 0.76 | No |
| | Parity | 2 | 5.42 | 0.07 | No | 5.41 | 0.07 | No | 5.78 | 0.06 | No |
| | **GLOBAL** | **19** | **26.11** | **0.13** | **No** | **30.58** | **0.05** | **Yes** | **20.63** | **0.36** | **No** |
| Mixed UI | Prevalent exposure | 1 | 0.37 | 0.54 | No | 0.38 | 0.54 | No | 0.11 | 0.74 | No |
| | Age at recruitment | 1 | 0.18 | 0.67 | No | 0.18 | 0.67 | No | 0.11 | 0.74 | No |
| | Ethnicity | 1 | 0.77 | 0.38 | No | 0.77 | 0.38 | No | 1.02E-03 | 0.97 | No |
| | TDI | 1 | 5.55 | 0.02 | Yes | 5.52 | 0.02 | Yes | 4.72 | 0.03 | Yes |
| | Cohabit with partner | 1 | 3.70E-04 | 0.99 | No | 4.47E-04 | 0.98 | No | 0.75 | 0.39 | No |
| | Education | 1 | 0.16 | 0.69 | No | 0.16 | 0.69 | No | 0.26 | 0.61 | No |
| | Alcohol | 2 | 1.89 | 0.39 | No | 1.89 | 0.39 | No | 1.30 | 0.52 | No |
| | Smoking | 2 | 2.14 | 0.34 | No | 2.14 | 0.34 | No | 1.89 | 0.39 | No |
| | Multi-morbidity | 1 | 4.59 | 0.03 | Yes | 4.59 | 0.03 | Yes | 2.24 | 0.13 | No |
| | BMI | 1 | 0.58 | 0.45 | No | 0.60 | 0.44 | No | 1.08 | 0.30 | No |
| | Menopause | 3 | 0.62 | 0.89 | No | 0.62 | 0.89 | No | 0.97 | 0.81 | No |
| | HRT | 1 | 0.13 | 0.72 | No | 0.13 | 0.72 | No | 0.30 | 0.58 | No |
| | Hysterectomy | 1 | 0.29 | 0.59 | No | 0.29 | 0.59 | No | 0.03 | 0.86 | No |
| | Parity | 2 | 1.20 | 0.55 | No | 1.21 | 0.55 | No | 0.15 | 0.93 | No |
| | **GLOBAL** | **19** | **20.67** | **0.36** | **No** | **20.90** | **0.34** | **No** | **13.74** | **0.80** | **No** |
| Any urgency | Prevalent exposure | 1 | 1.77 | 0.18 | No | 8.79 | 3.00E-03 | Yes | 1.41 | 0.24 | No |
| | Age at recruitment | 1 | 0.39 | 0.53 | No | 0.40 | 0.53 | No | 1.59 | 0.21 | No |
| | Ethnicity | 1 | 0.66 | 0.42 | No | 0.62 | 0.43 | No | 0.55 | 0.46 | No |
| | TDI | 1 | 0.90 | 0.34 | No | 0.87 | 0.35 | No | 0.29 | 0.59 | No |
| | Cohabit with partner | 1 | 0.32 | 0.57 | No | 0.33 | 0.56 | No | 0.09 | 0.76 | No |
| | Education | 1 | 1.49 | 0.22 | No | 1.50 | 0.22 | No | 2.91 | 0.09 | No |
| | Alcohol | 2 | 0.69 | 0.71 | No | 0.69 | 0.71 | No | 0.60 | 0.74 | No |
| | Smoking | 2 | 0.77 | 0.68 | No | 0.76 | 0.68 | No | 0.62 | 0.73 | No |

| | | | | | | | | | | | |
|---|---|---|---|---|---|---|---|---|---|---|---|
| | Multi-morbidity | 1 | 1.42 | 0.23 | No | 1.47 | 0.23 | No | 0.69 | 0.41 | No |
| | BMI | 1 | 0.45 | 0.50 | No | 0.47 | 0.49 | No | 0.21 | 0.64 | No |
| | Menopause | 3 | 8.50 | 0.04 | Yes | 8.50 | 0.04 | Yes | 6.50 | 0.09 | No |
| | HRT | 1 | 0.66 | 0.42 | No | 0.69 | 0.41 | No | 0.95 | 0.33 | No |
| | Hysterectomy | 1 | 0.10 | 0.75 | No | 0.11 | 0.75 | No | 0.58 | 0.45 | No |
| | Parity | 2 | 7.46 | 0.02 | Yes | 7.44 | 0.02 | Yes | 6.84 | 0.03 | Yes |
| | **GLOBAL** | **19** | **27.68** | **0.09** | **No** | **34.71** | **0.02** | **Yes** | **20.77** | **0.35** | **No** |

*UI urinary incontinence; TDI Townsend deprivation index; BMI body mass index, HRT hormone replacement therapy; df degrees of freedom;* $\chi^2$ *Chi-squared; PH proportional hazards.*

Table 11 Cox regression analyses between mental health and urinary incontinence in UK Biobank women (EHR and HES samples)

| Data source | Association | Model 1 | | | Model 2 | | | Model 3 | | |
|---|---|---|---|---|---|---|---|---|---|---|
| | | HR | 95% CI | p-value | HR | 95% CI | p-value | HR | 95% CI | p-value |
| EHR | Depression → Any UI | 1.94 | 1.77, 2.11 | <0.0001 | 1.89 | 1.73, 2.06 | <0.0001 | 1.70 | 1.55, 1.87 | <0.0001 |
| | Depression → Stress UI | 1.85 | 1.67, 2.06 | <0.0001 | 1.80 | 1.62, 1.99 | <0.0001 | 1.62 | 1.45, 1.81 | <0.0001 |
| | Depression → Urge UI | 2.04 | 1.82, 2.30 | <0.0001 | 2.04 | 1.82, 2.29 | <0.0001 | 1.84 | 1.64, 2.07 | <0.0001 |
| | Depression → Mixed UI | 2.53 | 1.88, 3.40 | <0.0001 | 2.41 | 1.78, 3.27 | <0.0001 | 2.16 | 1.62, 2.88 | <0.0001 |
| | Depression → Any urgency[1] | 1.65 | 1.51, 1.81 | <0.0001 | 1.67 | 1.53, 1.82 | <0.0001 | 1.53 | 1.40, 1.66 | <0.0001 |
| | Anxiety → Any UI | 1.57 | 1.38, 1.78 | <0.0001 | 1.53 | 1.35, 1.73 | <0.0001 | 1.45 | 1.28, 1.65 | <0.0001 |
| | Anxiety → Stress UI | 1.68 | 1.40, 2.01 | <0.0001 | 1.65 | 1.38, 1.97 | <0.0001 | 1.56 | 1.31, 1.85 | <0.0001 |
| | Anxiety → Urge UI | 1.49 | 1.20, 1.84 | <0.0001 | 1.45 | 1.17, 1.79 | 0.001 | 1.38 | 1.12, 1.70 | 0.003 |
| | Anxiety → Mixed UI | 2.20 | 1.44, 3.35 | <0.0001 | 2.11 | 1.38, 3.22 | 0.001 | 2.00 | 1.32, 3.01 | 0.001 |
| | Anxiety → Any urgency[1] | 1.41 | 1.23, 1.63 | <0.0001 | 1.39 | 1.21, 1.60 | <0.0001 | 1.33 | 1.15, 1.53 | <0.0001 |
| | Neuroticism → Any UI | 1.08 | 1.06, 1.09 | <0.0001 | 1.07 | 1.06, 1.08 | <0.0001 | 1.06 | 1.05, 1.07 | <0.0001 |
| | Neuroticism → Stress UI | 1.08 | 1.07, 1.10 | <0.0001 | 1.07 | 1.06, 1.08 | <0.0001 | 1.06 | 1.06, 1.07 | <0.0001 |
| | Neuroticism → Urge UI | 1.07 | 1.04, 1.10 | <0.0001 | 1.07 | 1.05, 1.10 | <0.0001 | 1.06 | 1.04, 1.10 | <0.0001 |
| | Neuroticism → Mixed UI | 1.09 | 1.02, 1.16 | 0.011 | 1.08 | 1.01, 1.15 | 0.019 | 1.07 | 1.00, 1.14 | 0.056 |
| | Neuroticism → Any urgency[1] | 1.06 | 1.04, 1.07 | <0.0001 | 1.06 | 1.05, 1.08 | <0.0001 | 1.05 | 1.04, 1.07 | <0.0001 |
| HES | Depression → Any UI | 2.60 | 2.19, 3.09 | <0.0001 | 2.42 | 2.02, 2.90 | <0.0001 | 1.87 | 1.55, 2.24 | <0.0001 |
| | Depression → Stress UI | 2.32 | 1.85, 2.91 | <0.0001 | 2.14 | 1.69, 2.71 | <0.0001 | 1.65 | 1.29, 2.12 | <0.0001 |
| | Depression → Urge UI | 3.41 | 2.57, 4.53 | <0.0001 | 3.16 | 2.36, 4.23 | <0.0001 | 2.28 | 1.71, 3.03 | <0.0001 |
| | Depression → Mixed UI | 3.04 | 1.68, 5.50 | <0.0001 | 2.62 | 1.41, 4.88 | 0.002 | 1.77 | 0.92, 3.38 | 0.086 |
| | Depression → Any urgency[1] | NA | NA | NA | NA | NA | NA | NA | NA | NA |
| | Anxiety → Any UI | 2.70 | 2.07, 3.51 | <0.0001 | 2.50 | 1.92, 3.26 | <0.0001 | 1.98 | 1.50, 2.62 | <0.0001 |
| | Anxiety → Stress UI | 2.42 | 1.63, 3.60 | <0.0001 | 2.24 | 1.51, 3.34 | <0.0001 | 1.77 | 1.18, 2.66 | 0.006 |
| | Anxiety → Urge UI | 4.26 | 3.14, 5.77 | <0.0001 | 3.89 | 2.86, 5.28 | <0.0001 | 3.00 | 2.19, 4.12 | <0.0001 |
| | Anxiety → Mixed UI | 5.17 | 2.72, 9.85 | <0.0001 | 4.58 | 2.40, 8.75 | <0.0001 | 3.31 | 1.74, 6.31 | <0.0001 |
| | Anxiety → Any urgency[1] | NA | NA | NA | NA | NA | NA | NA | NA | NA |
| | Neuroticism → Any UI | 1.08 | 1.07, 1.09 | <0.0001 | 1.08 | 1.07, 1.09 | <0.0001 | 1.06 | 1.05, 1.08 | <0.0001 |
| | Neuroticism → Stress UI | 1.08 | 1.07, 1.09 | <0.0001 | 1.07 | 1.06, 1.08 | <0.0001 | 1.06 | 1.05, 1.07 | <0.0001 |
| | Neuroticism → Urge UI | 1.11 | 1.09, 1.14 | <0.0001 | 1.11 | 1.09, 1.14 | <0.0001 | 1.09 | 1.07, 1.12 | <0.0001 |
| | Neuroticism → Mixed UI | 1.15 | 1.11, 1.19 | <0.0001 | 1.14 | 1.10, 1.18 | <0.0001 | 1.12 | 1.08, 1.16 | <0.0001 |

| Data source | Association | Model 1 | | | Model 2 | | | Model 3 | | |
|---|---|---|---|---|---|---|---|---|---|---|
| | | HR | 95% CI | p-value | HR | 95% CI | p-value | HR | 95% CI | p-value |
| | Neuroticism → Any urgency[1] | NA | NA | NA | NA | NA | NA | NA | NA | NA |

*EHR Electronic Health Records (primary care records); HES Hospital Episode Statistics (hospital inpatient records); HR Hazard ratio; CI confidence interval; UI Urinary Incontinence; NA not applicable. [1] Any urgency reported using the EHR sample only; Model 1 unadjusted; Model 2 adjusted for age, ethnicity, TDI, education, and cohabiting status; Model 3 additionally adjusted for alcohol, smoking, BMI, menopause, HRT, hysterectomy, and parity.*

Table 12 Cox regression analyses between urinary incontinence and mental health in UK Biobank women (EHR and HES samples)

| Data source | Association | Model 1 | | | Model 2 | | | Model 3 | | |
|---|---|---|---|---|---|---|---|---|---|---|
| | | HR | 95% CI | p-value | HR | 95% CI | p-value | HR | 95% CI | p-value |
| EHR | Depression → Any UI | 1.52 | 1.36, 1.71 | <0.0001 | 1.65 | 1.46, 1.86 | <0.0001 | 1.45 | 1.29, 1.64 | <0.0001 |
| | Depression → Stress UI | 1.50 | 1.35, 1.68 | <0.0001 | 1.63 | 1.46, 1.82 | <0.0001 | 1.43 | 1.27, 1.61 | <0.0001 |
| | Depression → Urge UI | 1.56 | 1.25, 1.96 | <0.0001 | 1.75 | 1.40, 2.18 | <0.0001 | 1.56 | 1.25, 1.95 | <0.0001 |
| | Depression → Mixed UI | 1.96 | 1.37, 2.80 | <0.0001 | 2.15 | 1.50, 3.08 | <0.0001 | 1.78 | 1.26, 2.52 | 0.001 |
| | Depression → Any urgency[1] | 1.24 | 1.00, 1.53 | 0.047 | 1.37 | 1.11, 1.70 | 0.003 | 1.25 | 1.01, 1.55 | 0.037 |
| | Anxiety → Any UI | 1.29 | 1.16, 1.43 | <0.0001 | 1.32 | 1.19, 1.46 | <0.0001 | 1.24 | 1.12, 1.38 | <0.0001 |
| | Anxiety → Stress UI | 1.30 | 1.17, 1.45 | <0.0001 | 1.33 | 1.20, 1.48 | <0.0001 | 1.25 | 1.12, 1.39 | <0.0001 |
| | Anxiety → Urge UI | 1.23 | 0.94, 1.62 | 0.128 | 1.28 | 0.97, 1.69 | 0.078 | 1.23 | 0.92, 1.63 | 0.156 |
| | Anxiety → Mixed UI | 1.85 | 1.18, 2.88 | 0.007 | 1.88 | 1.21, 2.93 | 0.005 | 1.76 | 1.15, 2.70 | 0.009 |
| | Anxiety → Any urgency[1] | 1.23 | 0.99, 1.51 | 0.057 | 1.27 | 1.03, 1.58 | 0.028 | 1.22 | 0.99, 1.50 | 0.064 |
| HES | Depression → Any UI | 2.07 | 1.88, 2.28 | <0.0001 | 1.95 | 1.76, 2.16 | <0.0001 | 1.54 | 1.39, 1.71 | <0.0001 |
| | Depression → Stress UI | 2.06 | 1.86, 2.28 | <0.0001 | 1.96 | 1.76, 2.17 | <0.0001 | 1.54 | 1.39, 1.72 | <0.0001 |
| | Depression → Urge UI | 2.46 | 1.97, 3.08 | <0.0001 | 2.21 | 1.75, 2.79 | <0.0001 | 1.71 | 1.37, 2.14 | <0.0001 |
| | Depression → Mixed UI | 3.23 | 2.58, 4.04 | <0.0001 | 2.91 | 2.32, 3.65 | <0.0001 | 2.13 | 1.68, 2.70 | <0.0001 |
| | Depression → Any urgency[1] | NA | NA | NA | NA | NA | NA | NA | NA | NA |
| | Anxiety → Any UI | 1.73 | 1.53, 1.95 | <0.0001 | 1.58 | 1.40, 1.80 | <0.0001 | 1.34 | 1.17, 1.52 | <0.0001 |
| | Anxiety → Stress UI | 1.68 | 1.49, 1.90 | <0.0001 | 1.55 | 1.37, 1.76 | <0.0001 | 1.30 | 1.14, 1.49 | <0.0001 |
| | Anxiety → Urge UI | 2.08 | 1.67, 2.59 | <0.0001 | 1.81 | 1.43, 2.28 | <0.0001 | 1.49 | 1.19, 1.87 | <0.0001 |
| | Anxiety → Mixed UI | 2.44 | 1.84, 3.23 | <0.0001 | 2.13 | 1.60, 2.86 | <0.0001 | 1.71 | 1.27, 2.32 | <0.0001 |
| | Anxiety → Any urgency[1] | NA | NA | NA | NA | NA | NA | NA | NA | NA |

*EHR Electronic Health Records (primary care records); HES Hospital Episode Statistics (hospital inpatient records); HR Hazard ratio; CI confidence interval; UI Urinary Incontinence; NA not applicable. [1] Any urgency reported using the EHR sample only; Model 1 unadjusted; Model 2 adjusted for age, ethnicity, TDI, education, and cohabiting status; Model 3 additionally adjusted for alcohol, smoking, BMI, menopause, HRT, hysterectomy, and parity.*

Table 13 Sensitivity analyses for the cox proportional hazard models

| Exposure | Outcome | HR (95% CI) - Main | HR (95% CI) - Sensitivity 1 | HR (95% CI) - Sensitivity 2 | HR (95% CI) - Sensitivity 3 | HR (95% CI) - Sensitivity 4 |
|---|---|---|---|---|---|---|
| Depression | AUI | 1.67 (1.55, 1.81) | 1.68 (1.56, 1.81) | 1.67 (1.54, 1.82) | 1.66 (1.54, 1.80) | NA |
| | SUI | 1.55 (1.43, 1.70) | 1.57 (1.44, 1.71) | 1.53 (1.37, 1.70) | 1.55 (1.41, 1.71) | 1.54 (1.42, 1.67) |
| | UUI | 1.80 (1.63, 1.98) | 1.80 (1.64, 1.98) | 1.76 (1.61, 1.93) | 1.73 (1.56, 1.93) | 1.77 (1.56, 2.01) |
| | MUI | 1.91 (1.59, 2.31) | 1.91 (1.57, 2.34) | 1.75 (1.39, 2.19) | 1.82 (1.48, 2.25) | NA |
| | Any urgency | 1.53(1.41, 1.65) | 1.52 (1.40, 1.65) | 1.50 (1.39, 1.62) | 1.49 (1.38, 1.62) | NA |
| Anxiety | AUI | 1.42 (1.27, 1.58) | 1.41 (1.26, 1.57) | 1.46 (1.30, 1.62) | 1.41 (1.26, 1.57) | NA |
| | SUI | 1.46 (1.25, 1.69) | 1.46 (1.26, 1.70) | 1.46 (1.20, 1.77) | 1.45 (1.25, 1.69) | 1.47 (1.26, 1.72) |
| | UUI | 1.38 (1.13, 1.68) | 1.37 (1.12, 1.67) | 1.35 (1.11, 1.63) | 1.33 (1.10, 1.61) | 1.45 (1.16, 1.81) |
| | MUI | 1.64 (1.24, 2.17) | 1.64 (1.23, 2.19) | 1.47 (0.98, 2.20) | 1.59 (1.24, 2.03) | NA |
| | Any urgency | 1.30 (1.13, 1.49) | 1.29 (1.12, 1.49) | 1.24 (1.06, 1.45) | 1.28 (1.12, 1.46) | NA |
| Neuroticism | AUI | 1.06 (1.05, 1.07) | 1.06 (1.05, 1.07) | 1.06 (1.05, 1.07) | NA | NA |
| | SUI | 1.06 (1.05, 1.07) | 1.06 (1.06, 1.07) | 1.06 (1.05, 1.07) | NA | 1.06 (1.05, 1.07) |
| | UUI | 1.06 (1.04, 1.09) | 1.06 (1.04, 1.08) | 1.06 (1.04, 1.08) | NA | 1.06 (1.04, 1.09) |
| | MUI | 1.08 (1.04, 1.12) | 1.08 (1.03, 1.12) | 1.06 (1.00, 1.13) | NA | NA |
| | Any urgency | 1.05 (1.04, 1.07) | 1.05 (1.04, 1.07) | 1.05 (1.04, 1.06) | NA | NA |
| AUI | Depression | 1.40 (1.27, 1.54) | 1.38 (1.27, 1.51) | 1.29 (1.17, 1.41) | 1.38 (1.26, 1.53) | NA |
| SUI | | 1.38 (1.25, 1.53) | 1.37 (1.24, 1.50) | 1.28 (1.14, 1.43) | 1.36 (1.22, 1.52) | 1.37 (1.25, 1.51) |
| UUI | | 1.55 (1.31, 1.83) | 1.52 (1.29, 1.80) | 1.37 (1.14, 1.66) | 1.59 (1.33, 1.90) | 1.60 (1.25, 2.04) |
| MUI | | 1.90 (1.58, 2.29) | 1.86 (1.52, 2.27) | 1.49 (1.10, 2.03) | 1.94 (1.56, 2.41) | NA |
| Any urgency | | 1.21 (1.04, 1.41) | 1.21 (1.04, 1.42) | 1.14 (0.93, 1.39) | 1.21 (1.02, 1.42) | NA |
| AUI | Anxiety | 1.28 (1.17, 1.39) | 1.27 (1.17, 1.38) | 1.22 (1.11, 1.35) | 1.26 (1.17, 1.36) | NA |
| SUI | | 1.30 (1.16, 1.45) | 1.29 (1.15, 1.45) | 1.25 (1.09, 1.43) | 1.28 (1.15, 1.43) | 1.28 (1.15, 1.44) |
| UUI | | 1.34 (1.02, 1.75) | 1.34 (1.03, 1.75) | 1.20 (0.88, 1.62) | 1.33 (1.00, 1.77) | 1.30 (0.94, 1.81) |
| MUI | | 1.58 (1.18, 2.12) | 1.58 (1.19, 2.11) | 1.14 (0.80, 1.63) | 1.50 (1.12, 2.01) | NA |
| Any urgency | | 1.30 (0.99, 1.71) | 1.30 (0.99, 1.71) | 1.25 (0.91, 1.72) | 1.29 (0.98, 1.71) | NA |

*AUI any Urinary Incontinence (UI); SUI stress UI; UUI urgency UI; MUI mixed UI; NA not applicable; HR hazards ratio; CI confidence interval. Analysis performed using combined EHR/HES sample with adjustment for confounders (Model 3). Sensitivity 1 stratifies by select confounders; Sensitivity 2 restricts to outcomes that are >2 years follow-up post baseline; Sensitivity 3 excludes other psychiatric disorders; Sensitivity 4 mutually excludes SUI and UUI cases from each respective analysis.*

Table 14 Sensitivity analyses for the Cox proportional hazards models with mutual adjustment for depression and anxiety (Model 3 compared to Model 4)

| Association | Model 3 | | | Model 4 | | |
|---|---|---|---|---|---|---|
| | **HR** | **95% CI** | **P value** | **HR** | **95% CI** | **P value** |
| Depression → Any UI | 1.67 | 1.55, 1.81 | <0.0001 | 1.61 | 1.51, 1.73 | <0.0001 |
| Depression → Stress UI | 1.55 | 1.43, 1.70 | <0.0001 | 1.49 | 1.37, 1.61 | <0.0001 |
| Depression → Urge UI | 1.8 | 1.63, 1.98 | <0.0001 | 1.75 | 1.61, 1.90 | <0.0001 |
| Depression → Mixed UI | 1.91 | 1.59, 2.31 | <0.0001 | 1.81 | 1.51, 2.16 | <0.0001 |
| Depression → Any urgency[1] | 1.53 | 1.41, 1.65 | <0.0001 | 1.49 | 1.36, 1.63 | <0.0001 |
| Anxiety → Any UI | 1.42 | 1.27, 1.58 | <0.0001 | 1.24 | 1.12, 1.37 | <0.0001 |
| Anxiety → Stress UI | 1.46 | 1.25, 1.69 | <0.0001 | 1.30 | 1.12, 1.51 | <0.0001 |
| Anxiety → Urge UI | 1.38 | 1.13, 1.68 | 0.002 | 1.17 | 0.97, 1.41 | 0.100 |
| Anxiety → Mixed UI | 1.64 | 1.24, 2.17 | 0.001 | 1.38 | 1.05, 1.81 | 0.019 |
| Anxiety → Any urgency[1] | 1.30 | 1.13, 1.49 | <0.0001 | 1.16 | 1.00, 1.34 | 0.047 |
| Any UI → Depression | 1.4 | 1.27, 1.54 | <0.0001 | 1.35 | 1.23, 1.49 | <0.0001 |
| Stress UI → Depression | 1.38 | 1.25, 1.53 | <0.0001 | 1.34 | 1.22, 1.48 | <0.0001 |
| Urgency UI → Depression | 1.55 | 1.31, 1.83 | <0.0001 | 1.51 | 1.27, 1.80 | <0.0001 |
| Mixed UI → Depression | 1.90 | 1.58, 2.29 | <0.0001 | 1.88 | 1.55, 2.27 | <0.0001 |
| Any urgency[1] → Depression | 1.21 | 1.04, 1.41 | 0.015 | 1.17 | 1.01, 1.36 | 0.037 |
| Any UI → Anxiety | 1.28 | 1.17, 1.39 | <0.0001 | 1.16 | 1.07, 1.26 | <0.0001 |
| Stress UI → Anxiety | 1.30 | 1.16, 1.45 | <0.0001 | 1.19 | 1.06, 1.33 | 0.003 |
| Urgency UI → Anxiety | 1.34 | 1.02, 1.75 | 0.033 | 1.23 | 0.95, 1.60 | 0.116 |
| Mixed UI → Anxiety | 1.58 | 1.18, 2.12 | 0.002 | 1.42 | 1.07, 1.88 | 0.016 |
| Any urgency[1] → Anxiety | 1.30 | 0.99, 1.71 | 0.061 | 1.19 | 0.91, 1.55 | 0.199 |

*UI urinary incontinence; HR hazards ratio; CI confidence interval; Model 3 adjusts for age, ethnicity, TDI, education, cohabiting status, alcohol, smoking, BMI, menopause, HRT, hysterectomy, and parity. Model 4 additionally mutually adjusts for depression or anxiety. We present Model 3 results again in this table for easy comparison to Model 4 results.*

Table 15 Two-sample Mendelian Randomisation of mental health and urinary incontinence

| Exposure | Outcome | IVW | | MR Egger | | Weighted median | | Weighted mode | |
|---|---|---|---|---|---|---|---|---|---|
| | | **OR (95% CI)** | **P value** | **OR (95% CI)** | **P value** | **OR (95% CI)** | **P value** | **OR (95% CI)** | **P value** |
| Depression | AUI | 1.25 (1.16, 1.35) | 6.4E-09 | 1.46 (1.02, 2.07) | 0.04 | 1.18 (1.07, 1.30) | 6.7E-04 | 1.10 (0.80, 1.52) | 0.55 |
| | SUI | 1.24 (1.13, 1.36) | 4.5E-06 | 1.38 (0.90, 2.11) | 0.14 | 1.16 (1.04, 1.30) | 0.01 | 1.08 (0.75, 1.57) | 0.67 |
| | UUI | 1.30 (1.14, 1.48) | 1.2E-04 | 1.43 (0.77, 1.48) | 0.26 | 1.23 (1.03, 1.47) | 0.03 | 1.10 (0.59, 2.03) | 0.77 |
| | MUI | 1.39 (1.12, 1.72) | 2.7E-03 | 1.95 (0.72, 5.29) | 0.19 | 1.57 (1.16, 2.13) | 3.7E-03 | 2.24 (0.89, 5.68) | 0.09 |
| Anxiety[a] | AUI | 1.03 (0.99, 1.06) | 0.17 | 1.32 (0.93, 1.89) | 0.13 | 1.01 (0.96, 1.07) | 0.62 | 1.01 (0.93, 1.10) | 0.80 |
| | SUI | 1.03 (0.99, 1.08) | 0.12 | 1.36 (0.91, 2.05) | 0.15 | 1.04 (0.98, 1.10) | 0.22 | 1.01 (0.91, 1.12) | 0.85 |
| | UUI | 1.04 (0.97, 1.12) | 0.23 | 1.19 (0.61, 2.33) | 0.62 | 1.03 (0.93, 1.13) | 0.58 | 1.02 (0.88, 1.18) | 0.84 |
| | MUI | 1.07 (0.95, 1.20) | 0.26 | 0.70 (0.21, 2.32) | 0.56 | 1.06 (0.90, 1.25) | 0.52 | 0.97 (0.74, 1.27) | 0.84 |
| Neuroticism | AUI | 1.50 (1.26, 1.78) | 3.2E-06 | 1.26 (0.58, 2.71) | 0.56 | 1.39 (1.11, 1.73) | 4.3E-03 | 1.19 (0.63, 2.26) | 0.59 |
| | SUI | 1.51 (1.24, 1.84) | 5.6E-05 | 1.01 (0.41, 2.49) | 0.98 | 1.59 (1.24, 2.05) | 2.5E-04 | 1.93 (0.92, 4.03) | 0.08 |
| | UUI | 1.69 (1.29, 2.21) | 1.4E-04 | 3.88 (1.15, 13.12) | 0.03 | 1.75 (1.17, 2.63) | 0.01 | 1.93 (0.63, 5.92) | 0.25 |
| | MUI | 2.47 (1.55, 3.93) | 1.5E-04 | 0.44 (0.05, 3.54) | 0.44 | 2.86 (1.48, 5.55) | 1.8E-03 | 3.77 (0.83, 17.10) | 0.09 |

*UI urinary incontinence; AUI any UI; SUI stress UI; UUI urgency UI; MUI mixed UI; IVW inverse-variance weighted; MR Mendelian randomisation; OR odds ratio; CI confidence interval. [a] For anxiety the IVW is debiased deriving from a more lenient P value of 5E-10, all other methods are also based on the more lenient instrument.*

Table 16 Two-Sample Mendelian randomisation of urinary incontinence and mental health: debiased IVW

| Exposure | Outcome | Debiased IVW | | | MR Egger | | Weighted median | | Weighted mode | |
|---|---|---|---|---|---|---|---|---|---|---|
| | | **Number of SNPs** | **OR/Beta (95% CI)** | **P value** | **OR/Beta (95% CI)** | **P value** | **OR/Beta (95% CI)** | **P value** | **OR/Beta (95% CI)** | **P value** |
| AUI | Depression | 28 | 1.02 (1.00, 1.03) | 0.02 | 1.00 (0.92, 1.07) | 0.91 | 1.00 (0.98, 1.03) | 0.67 | 1.00 (0.97, 1.03) | 0.95 |
| SUI | | 33 | 1.01 (1.00, 1.02) | 0.05 | 0.99 (0.93, 1.05) | 0.63 | 1.00 (0.99, 1.02) | 0.84 | 1.00 (0.98, 1.02) | 0.86 |
| UUI | | 9 | 1.00 (0.98, 1.01) | 0.71 | 0.99 (0.95, 1.03) | 0.70 | 0.99 (0.97, 1.01) | 0.21 | 0.99 (0.96, 1.01) | 0.33 |
| MUI | | 16 | 1.01 (1.00, 1.01) | 0.16 | 1.01 (1.00, 1.01) | 0.18 | 1.00 (0.99, 1.01) | 0.61 | 1.00 (0.98, 1.01) | 0.60 |
| AUI | Anxiety | 32 | 1.01 (0.96, 1.06) | 0.72 | 1.05 (0.96, 1.16) | 0.32 | 0.99 (0.93, 1.07) | 0.84 | 0.99 (0.92, 1.06) | 0.74 |
| SUI | | 36 | 1.00 (0.97, 1.04) | 0.83 | 1.00 (0.92, 1.09) | 0.96 | 1.00 (0.94, 1.05) | 0.88 | 1.00 (0.94, 1.06) | 0.98 |
| UUI | | 10 | 0.99 (0.95, 1.03) | 0.54 | 1.00 (0.93, 1.08) | 0.96 | 1.00 (0.95, 1.06) | 0.89 | 1.00 (0.95, 1.05) | 0.94 |
| MUI | | 19 | 1.00 (0.99, 1.02) | 0.61 | 1.02 (0.99, 1.05) | 0.27 | 1.00 (0.98, 1.01) | 0.78 | 1.00 (0.98, 1.02) | 0.77 |
| AUI | Neuroticism[a] | 34 | 0.01 (-0.001, 0.02) | 0.07 | 0.02 (-0.03, 0.07) | 0.44 | -0.004 (-0.02, 0.01) | 0.62 | -0.002 (-0.02, 0.02) | 0.82 |
| SUI | | 35 | 0.01 (1.4E-04, 0.02) | 0.05 | 0.01 (-0.03, 0.04) | 0.62 | 0.01 (-0.002, 0.02) | 0.10 | -0.002 (-0.02, 0.02) | 0.83 |
| UUI | | 10 | -0.004 (-0.01, 0.01) | 0.48 | -0.003 (-0.03, 0.02) | 0.82 | 0.002 (-0.01, 0.02) | 0.75 | 0.004 (-0.02, 0.02) | 0.70 |
| MUI | | 16 | 5.6E-05 (-0.004, 0.004) | 0.98 | 0.001 (-0.01, 0.01) | 0.85 | 0.001 (-0.01, 0.01) | 0.80 | 0.001 (-0.01, 0.01) | 0.89 |

*IVW inverse variance weighted; OR odds ratio; CI confidence interval; AUI any urinary incontinence (UI); SUI stress UI; UUI urge UI; MUI mixed UI. The causal effect estimates represent the OR and 95% CI for an approximate doubling of the genetic liability to each UI exposure. [a] The causal estimates for the outcome neuroticism represents change in neuroticism score (beta) per approximate doubling in liability to each UI exposure.*

Table 17 Single SNP Wald ratios for the causal effects between exposure to UI and mental health

| Exposure | Outcome | SNP | Wald ratio | |
|---|---|---|---|---|
| | | | OR (95% CI) | P value |
| AUI | Depression | rs10774741 | 0.992 (0.954, 1.033) | 0.708 |
| | | rs11770163 | 1.127 (1.059, 1.199) | 1.7E-04 |
| | | rs35783198 | 0.994 (0.934, 1.057) | 0.842 |
| SUI | | rs10774741 | 0.993 (0.959, 1.029) | 0.708 |
| | | rs10891477 | 0.984 (0.927, 1.045) | 0.607 |
| | | rs3858460 | 1.003 (0.952, 1.057) | 0.916 |
| | | rs6816327 | 0.992 (0.942, 1.045) | 0.772 |
| | | rs73234742 | 0.991 (0.943, 1.042) | 0.722 |
| UUI | | NA | NA | NA |
| MUI | | rs147030051 | 1.021 (1.001, 1.042) | 0.037 |
| AUI | Anxiety | rs10774741 | 0.906 (0.691, 1.188) | 0.476 |
| | | rs11770163 | 0.967 (0.643, 1.454) | 0.873 |
| | | rs35783198 | 0.893 (0.664, 1.200) | 0.454 |
| SUI | | rs10774741 | 0.916 (0.720, 1.166) | 0.476 |
| | | rs10891477 | 0.736 (0.507, 1.068) | 0.107 |
| | | rs3858460 | 1.102 (0.805, 1.509) | 0.543 |
| | | rs6816327 | 1.018 (0.756, 1.371) | 0.906 |
| | | rs73234742 | 0.917 (0.723, 1.161) | 0.470 |
| UUI | | NA | NA | NA |
| MUI | | rs147030051 | 0.971 (0.912, 1.034) | 0.354 |
| AUI | Neuroticism[a] | rs10774741 | -0.006 (-0.047, 0.036) | 0.794 |
| | | rs11770163 | 0.049 (-0.019, 0.116) | 0.162 |
| | | rs35783198 | -0.010 (-0.076, 0.056) | 0.762 |
| SUI | | rs10774741 | -0.005 (-0.042, 0.032) | 0.794 |
| | | rs10891477 | -0.124 (-0.183, -0.064) | 4.44E-05 |
| | | rs3858460 | 0.037 (-0.014, 0.089) | 0.155 |
| | | rs6816327 | 0.038 (-0.015, 0.091) | 0.159 |
| | | rs73234742 | -0.011 (-0.064, 0.041) | 0.671 |
| UUI | | NA | NA | NA |
| MUI | | rs147030051 | -0.015 (-0.036, 0.005) | 0.134 |

*AUI any urinary incontinence (UI); SUI stress UI; UUI urge UI; MUI mixed UI; OR odds ratio; CI confidence interval; NA not applicable*

Table 18 Mendelian randomisation sensitivity analyses for the bidirectional effects between mental health and urinary incontinence

| **Exposure** | **Outcome** | **Cochran's Q (P value)** | **Rücker's Q (P value)** | $\overline{F}$ | $(\overline{F} - 1) / \overline{F}$ | $I^2_{GX}$ | **MR Egger intercept P value** |
|---|---|---|---|---|---|---|---|
| Depression | AUI | 200.72 (0.002) | 199.72 (0.001) | 41.93 | 0.98 | 0 | 0.40 |
| | SUI | 220.74 (0.0001) | 220.34 (0.00004) | 41.93 | 0.98 | 0 | 0.61 |
| | UUI | 168.25 (0.09) | 168.13 (0.08) | 41.93 | 0.98 | 0 | 0.76 |
| | MUI | 158.72 (0.21) | 158.22 (0.20) | 41.93 | 0.98 | 0 | 0.50 |
| Anxiety | AUI | 7.45 (0.11) | 7.36 (0.06) | 34.68 | 0.97 | 0 | 0.86 |
| | SUI | 7.67 (0.10) | 7.67 (0.10) | 34.68 | 0.97 | 0 | 0.97 |
| | UUI | 10.23 (0.04) | 10.16 (0.02) | 34.68 | 0.97 | 0 | 0.89 |
| | MUI | 10.43 (0.03) | 10.03 (0.02) | 34.68 | 0.97 | 0 | 0.75 |
| Neuroticism | AUI | 187.34 (0.001) | 187.03 (0.001) | 42.82 | 0.98 | 0.14 | 0.65 |
| | SUI | 198.35 (0.0001) | 197.12 (0.0001) | 42.82 | 0.98 | 0.14 | 0.37 |
| | UUI | 126.76 (0.54) | 124.89 (0.56) | 42.82 | 0.98 | 0.14 | 0.17 |
| | MUI | 138.89 (0.26) | 135.96 (0.30) | 42.82 | 0.98 | 0.14 | 0.10 |
| AUI | Depression | 62.33 (0.0001) | 61.57 (0.0001) | 25.81 | 0.96 | 0 | 0.58 |
| | Anxiety | 30.58 (0.49) | 29.66 (0.48) | 25.34 | 0.96 | 0.60 | 0.35 |
| | Neuroticism | 69.49 (0.0002) | 69.09 (0.0002) | 25.17 | 0.96 | 0.86 | 0.67 |
| SUI | Depression | 97.50 (1.5E-08) | 95.03 (2.0E-08) | 26.34 | 0.96 | 0 | 0.38 |
| | Anxiety | 41.41 (0.211) | 41.37 (0.18) | 26.12 | 0.96 | 0.66 | 0.87 |
| | Neuroticism | 71.98 (0.0002) | 71.98 (0.0001) | 26.08 | 0.96 | 0.77 | 0.96 |
| UUI | Depression | 9.62 (0.29) | 9.49 (0.22) | 22.07 | 0.95 | 0 | 0.77 |
| | Anxiety | 6.25 (0.71) | 6.08 (0.64) | 22.76 | 0.96 | 0.74 | 0.70 |
| | Neuroticism | 10.88 (0.28) | 10.88 (0.21) | 22.76 | 0.96 | 0.80 | 0.99 |
| MUI | Depression | 16.57 (0.34) | 15.95 (0.32) | 23.72 | 0.96 | 0 | 0.47 |
| | Anxiety | 16.94 (0.53) | 15.87 (0.53) | 23.46 | 0.96 | 0.84 | 0.32 |
| | Neuroticism | 16.74 (0.33) | 16.69 (0.27) | 23.78 | 0.96 | 0.85 | 0.84 |

*AUI any urinary incontinence (UI); SUI stress UI; UUI urge UI; MUI mixed UI; MR Mendelian randomisation.* $\overline{F}$ *mean F-statistic;* $(\overline{F} - 1) / \overline{F}$ *degree of violation of the NOME (NO measurement error) IVW and for Egger (*$I^2_{GX}$*)*

Table 19 Results for MRLap analysis: bias correction for sample overlap

| Exposure | Outcome | Observed IVW | | Corrected IVW | |
|---|---|---|---|---|---|
| | | **OR** | **95% CI** | **OR** | **95% CI** |
| Anxiety | AUI | 1.12 | 0.94, 1.33 | 1.19 | 0.90, 1.57 |
| | SUI | 1.12 | 0.94, 1.34 | 1.2 | 0.90, 1.60 |
| | UUI | 1.05 | 0.85, 1.28 | 1.08 | 0.77, 1.50 |
| | MUI | 0.97 | 0.79, 1.20 | 0.96 | 0.69, 1.34 |
| Neuroticism | AUI | 1.19 | 1.11, 1.27 | 1.23 | 1.13, 1.35 |
| | SUI | 1.17 | 1.09, 1.25 | 1.21 | 1.10, 1.32 |
| | UUI | 1.12 | 1.06, 1.19 | 1.15 | 1.07, 1.24 |
| | MUI | 1.13 | 1.06, 1.20 | 1.17 | 1.08, 1.26 |

*IVW inverse variance weighted; OR odds ratio; CI confidence interval; AUI any urinary incontinence (UI); SUI stress UI; UUI urge UI; MUI mixed UI. Note that the causal effect estimates have not been rescaled to a per doubling of genetic liability to the exposure variables and represent a per unit increase in log-odds of the exposure.*
