## supplementary figure for "Depression and anxiety as causes and consequences of urinary incontinence in women: a population-based study"

Figure 1 Timeline of data collection and data linkage in the UK Biobank

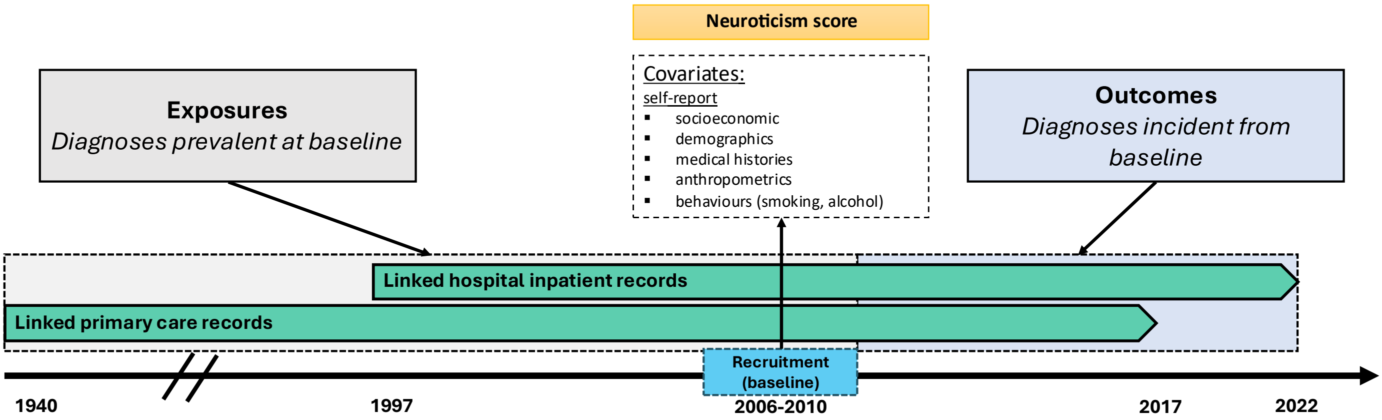

*Linked primary care records (Electronic Health Records; EHR) are available for approximately 45% of the UK Biobank cohort. Linked hospital inpatient records (Hospital Episode Statistics; HES) are available for all UK Biobank participants. Exposures prevalent at baseline and outcomes incident from baseline are derived using both EHR and HES records. Confounders of the proposed relationships are measured at baseline visit. Neuroticism was measured at baseline visit via questionnaire.*

Figure 2 Directed Acyclic Graph of the proposed bidirectional relationships between urinary incontinence and mental health

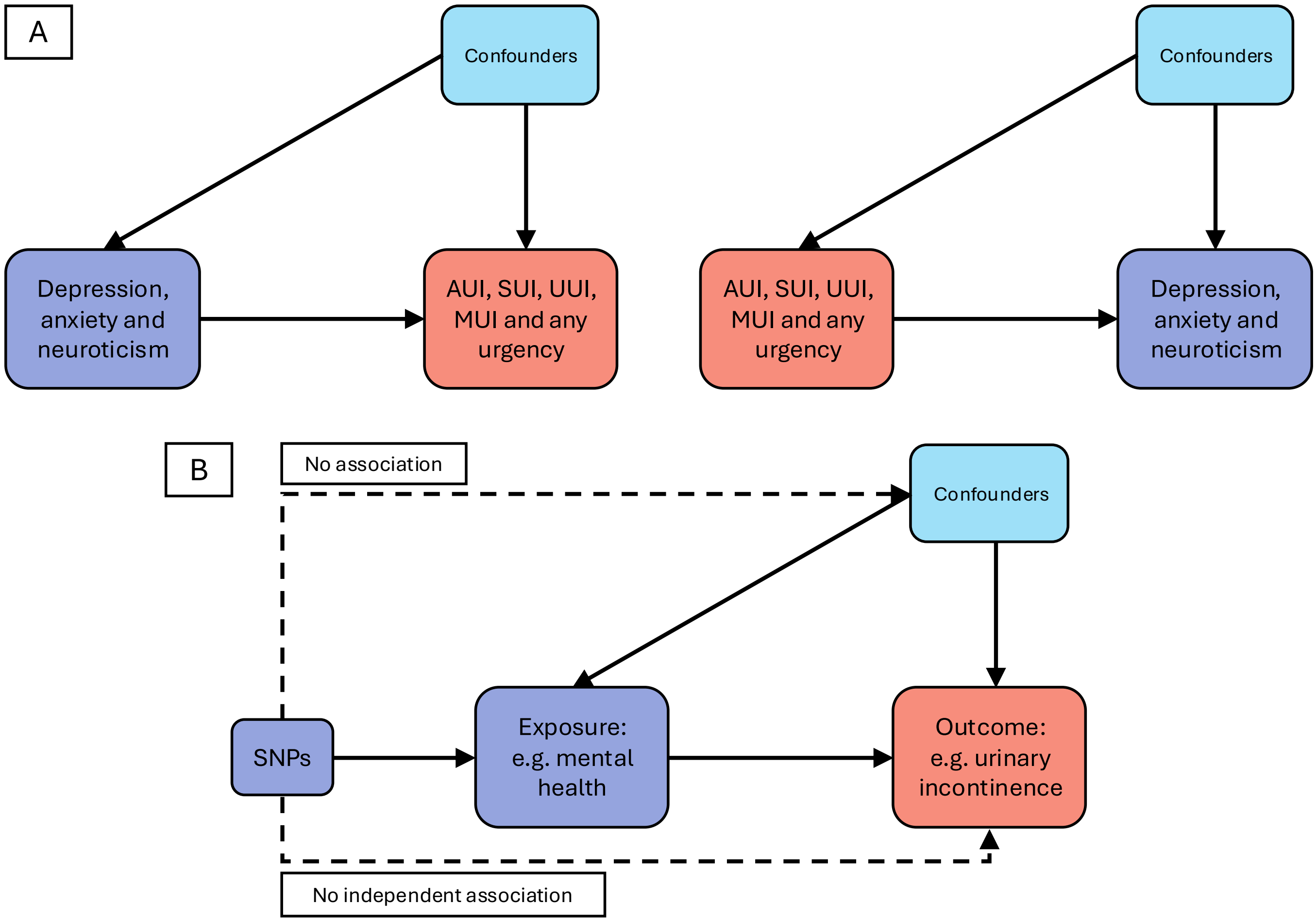

*Directed Acyclic Graph (DAG) illustrating **A:** the proposed bidirectional prospective associations between urinary incontinence and mental health. Confounders include age, ethnicity, Townsend deprivation index, education, cohabiting status, alcohol, smoking, BMI, menopause, HRT, hysterectomy, and parity; **B:** DAG illustrating the proposed causal relationships between an exposure e.g. mental health and an outcome e.g. urinary incontinence. SNPs associated with the exposure can be used as an instrument providing three core assumptions are met: (1) the genetic instrument is associated with the exposure (relevance), (2) there are no measured and unmeasured confounders of the genetic instrument and the outcome (independence), and (3) there is no independent pathway between the genetic instrument and the outcome other than through the exposure (exclusion restriction). UI urinary incontinence; AUI any UI; SUI stress UI; UUI urge UI; MUI mixed UI.*

Figure 3 Kaplan-Meier curves and number at risk: mental health exposures to urinary incontinence outcomes

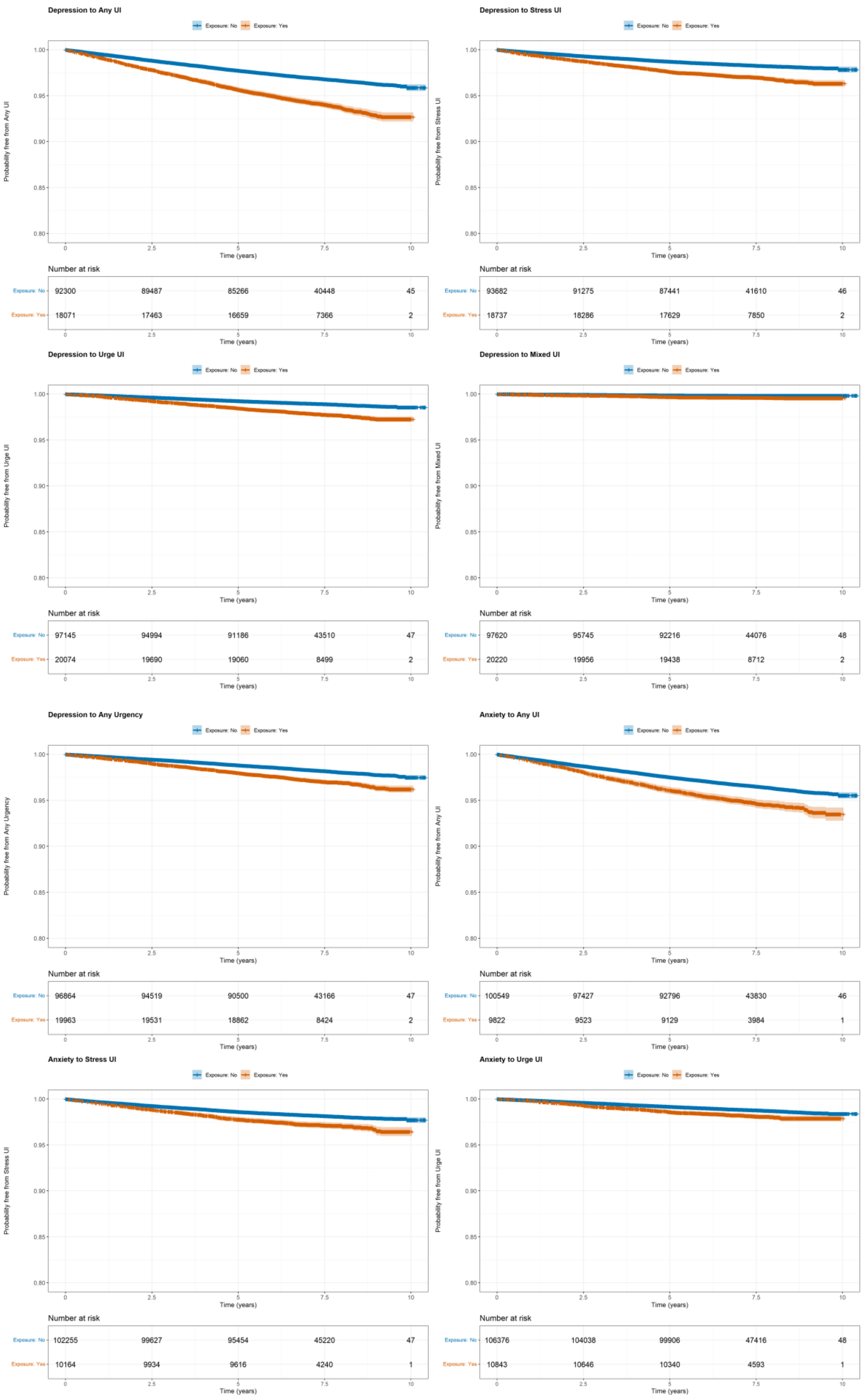

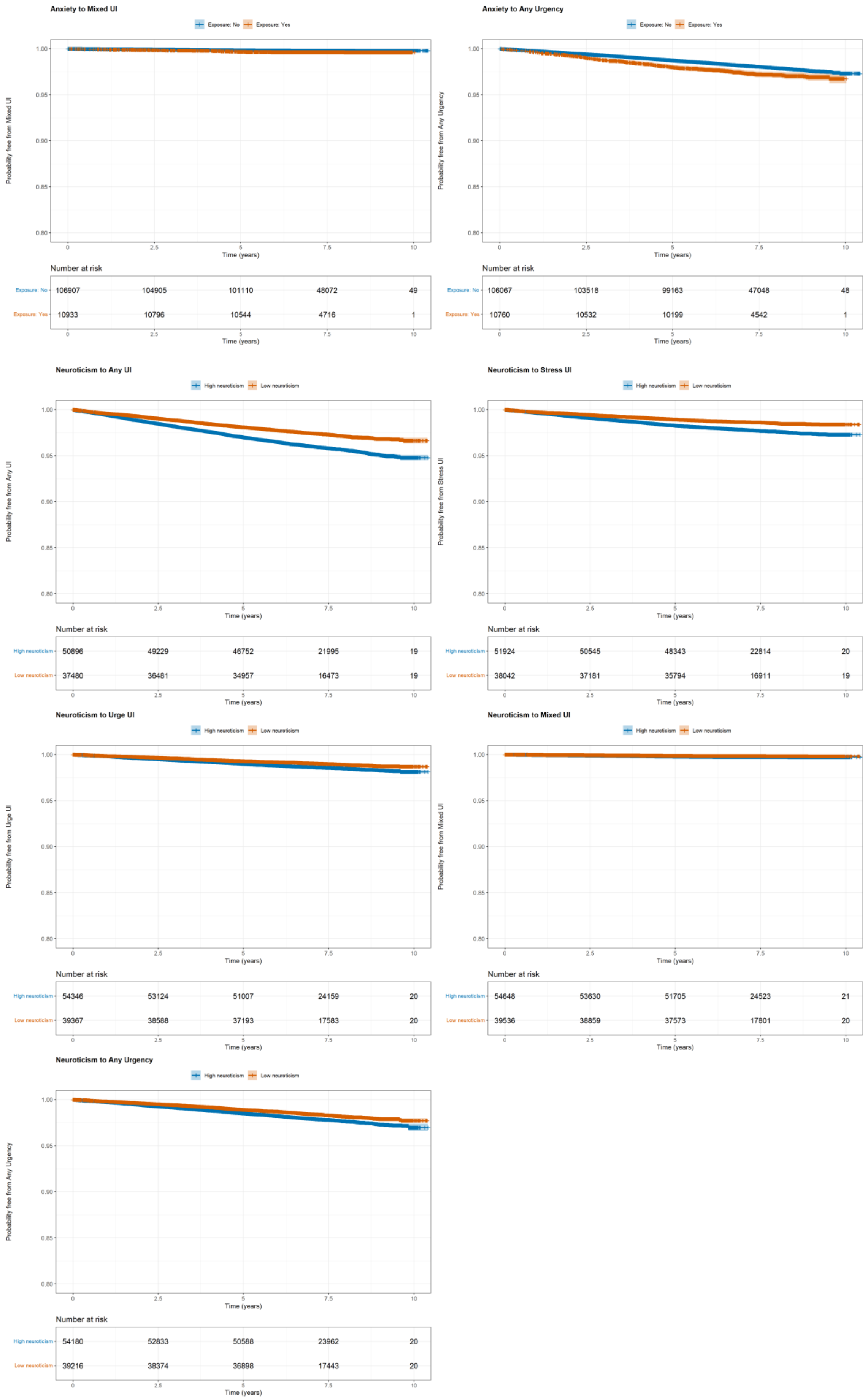

*UI urinary incontinence. Note that the neuroticism score was dichotomised into low and high neuroticism according to the median score (4.00) for the purposes of these plots.*

Figure 4 Kaplan-Meier curves and number at risk: urinary incontinence exposures to mental health outcomes

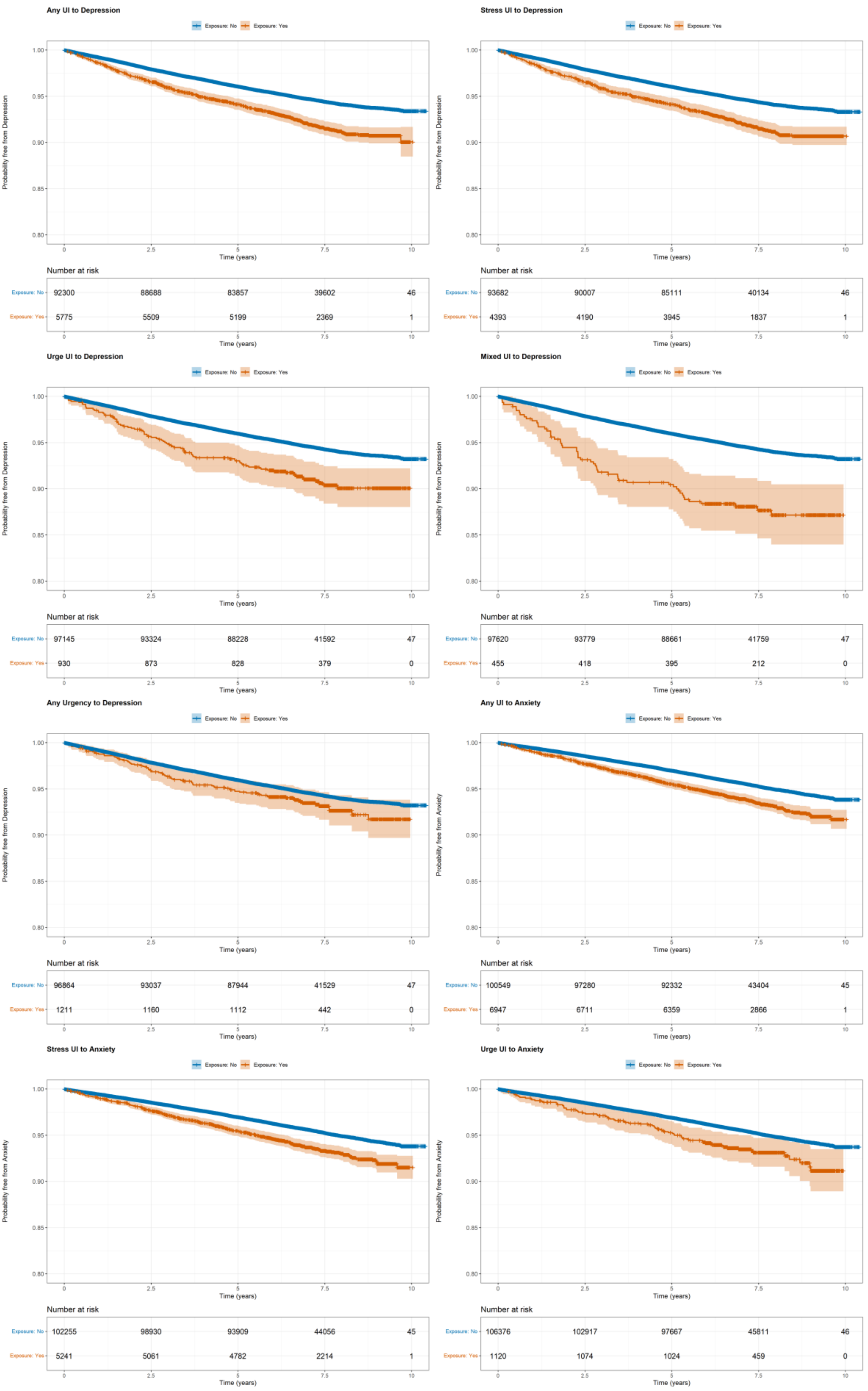

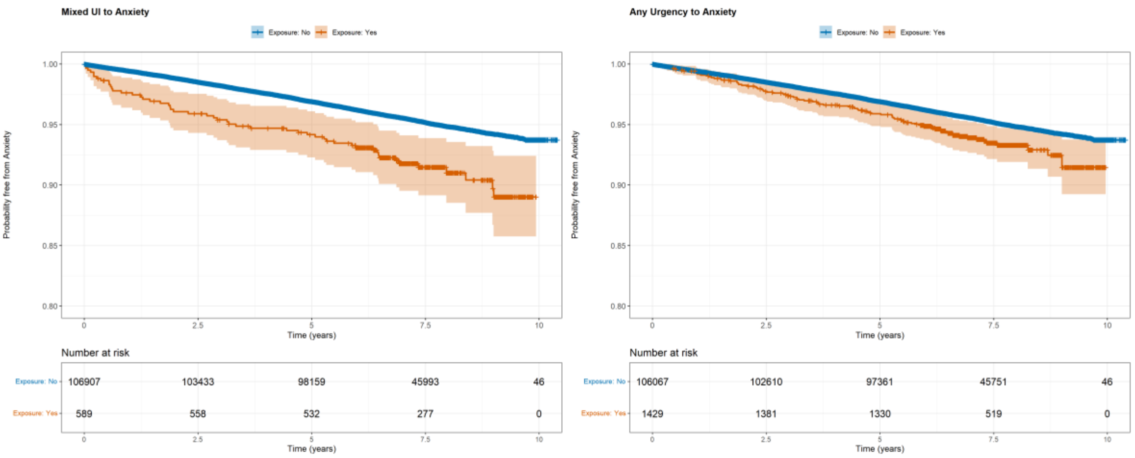

*UI urinary incontinence.*

Figure 5 Schoenfeld residual plots: mental health exposures to urinary incontinence outcomes

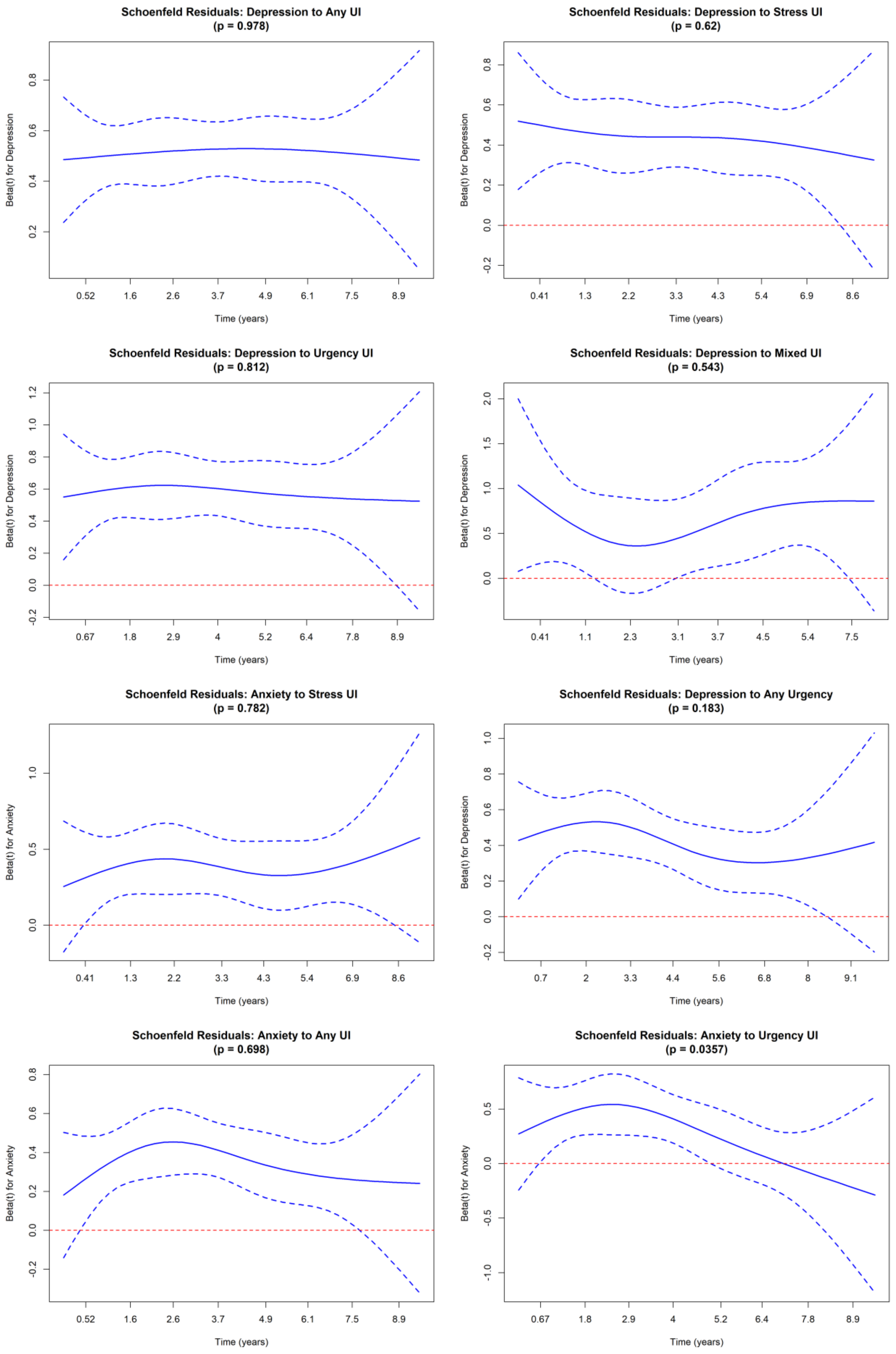

**Schoenfeld Residuals: Anxiety to Mixed UI**
**(p = 0.54)**

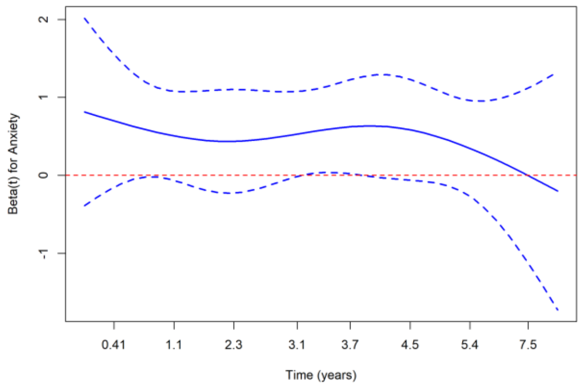

**Schoenfeld Residuals: Anxiety to Any Urgency**
**(p = 0.00303)**

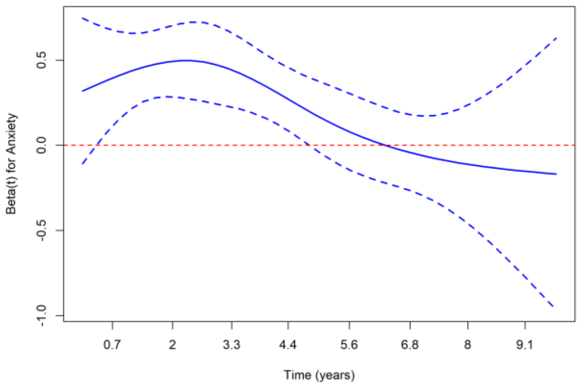

**Schoenfeld Residuals: Neuroticism to Any UI**
**(p = 0.88)**

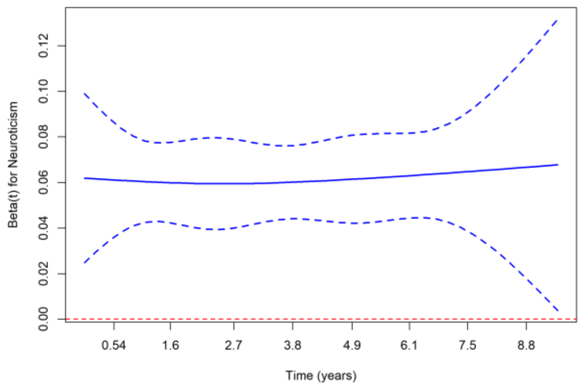

**Schoenfeld Residuals: Neuroticism to Stress UI**
**(p = 0.456)**

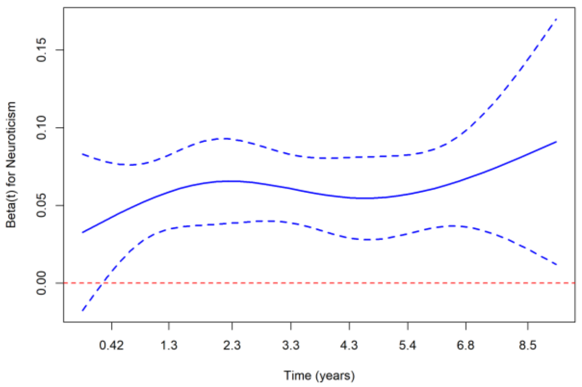

**Schoenfeld Residuals: Neuroticism to Urgency UI**
**(p = 0.299)**

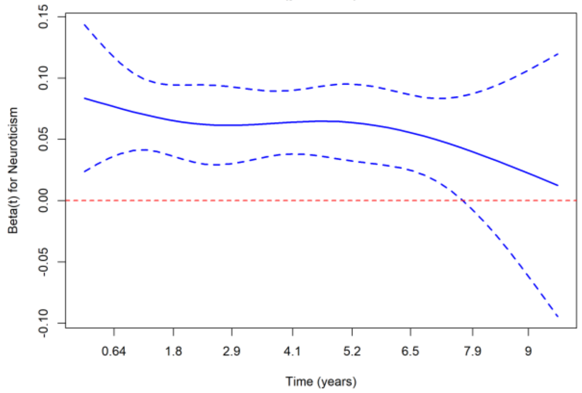

**Schoenfeld Residuals: Neuroticism to Mixed UI**
**(p = 0.744)**

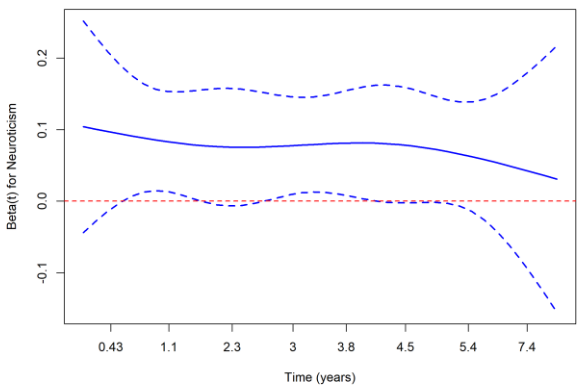

**Schoenfeld Residuals: Neuroticism to Any Urgency**
**(p = 0.235)**

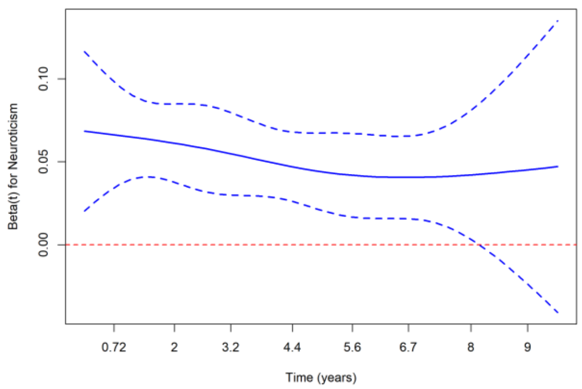

*UI urinary incontinence*

Figure 6 Schoenfeld residual plots: urinary incontinence exposures to mental health outcomes

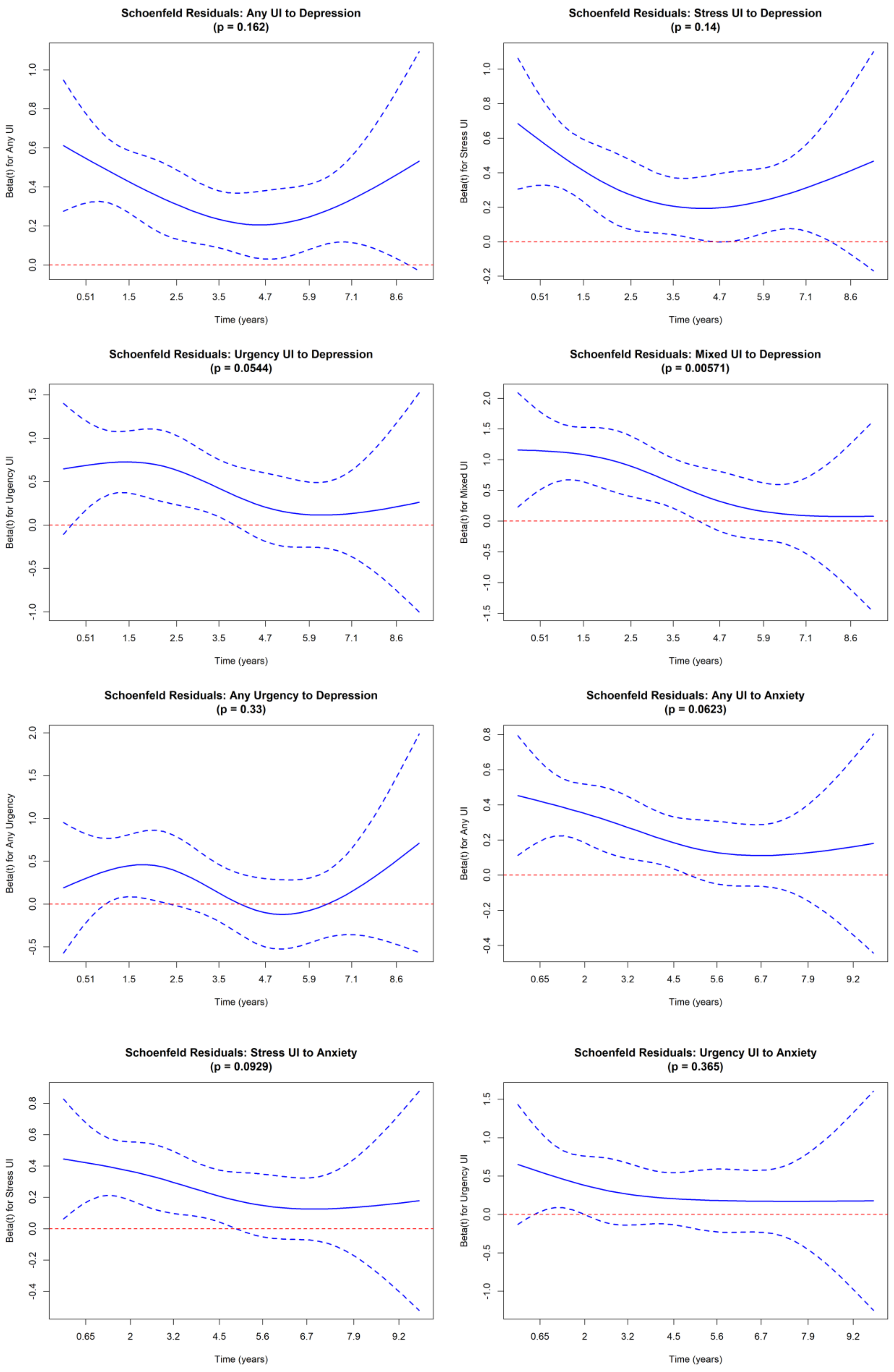

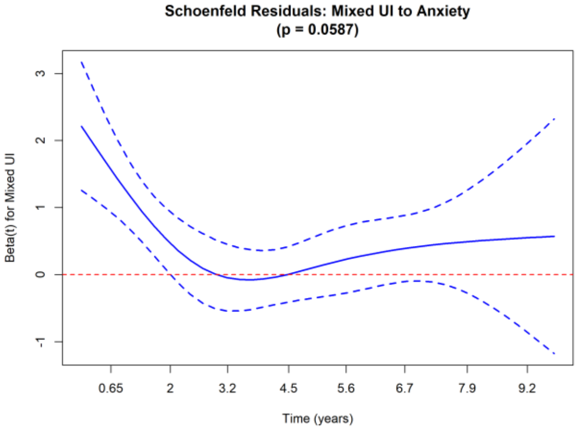

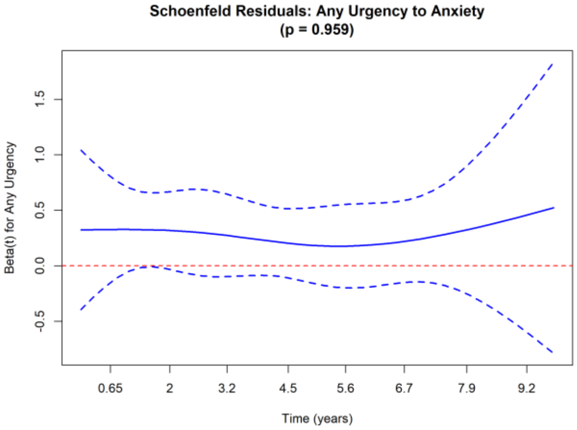

*UI urinary incontinence*

Figure 7 Leave-one-out plots for Mendelian randomisation of the genetic liability to depression, anxiety, and neuroticism with urinary incontinence outcomes

*The X axis shows the log-odds of each outcome per unit increase in the genetic liability to each exposure.*

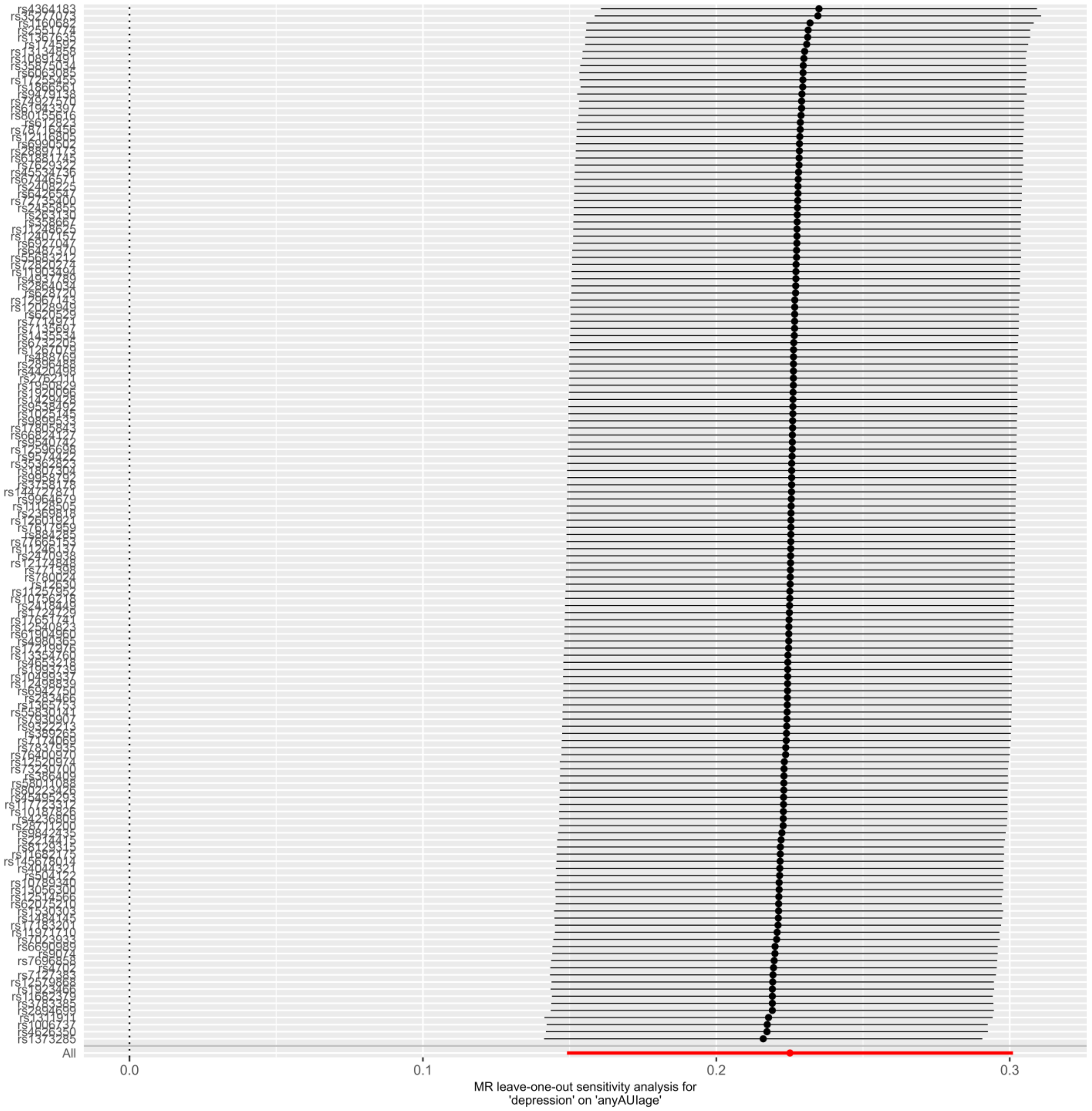

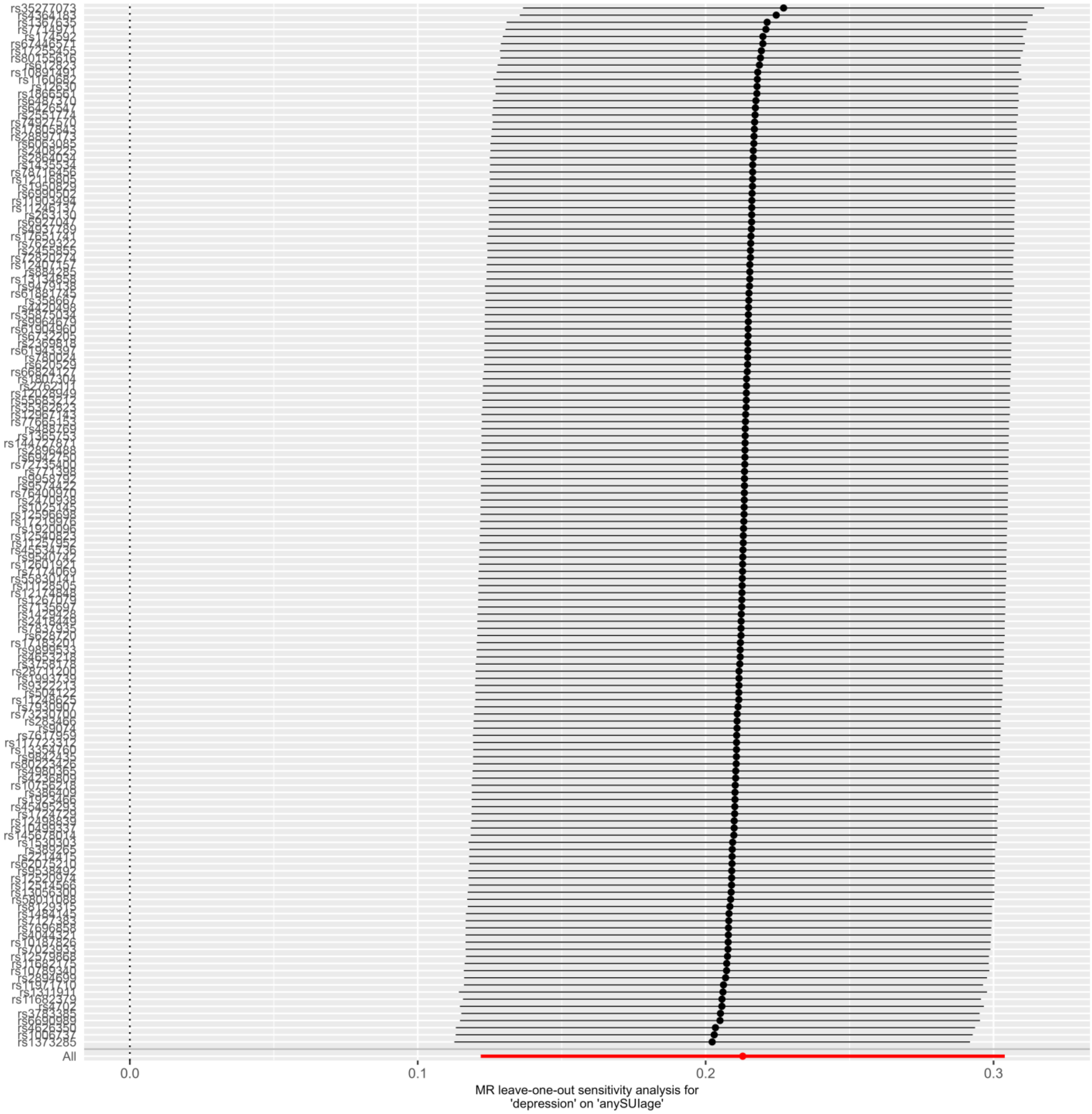
rs35277073
rs1373285
All
0.0
0.1
0.2
0.3
MR leave-one-out sensitivity analysis for
'depression' on 'anySUIage'

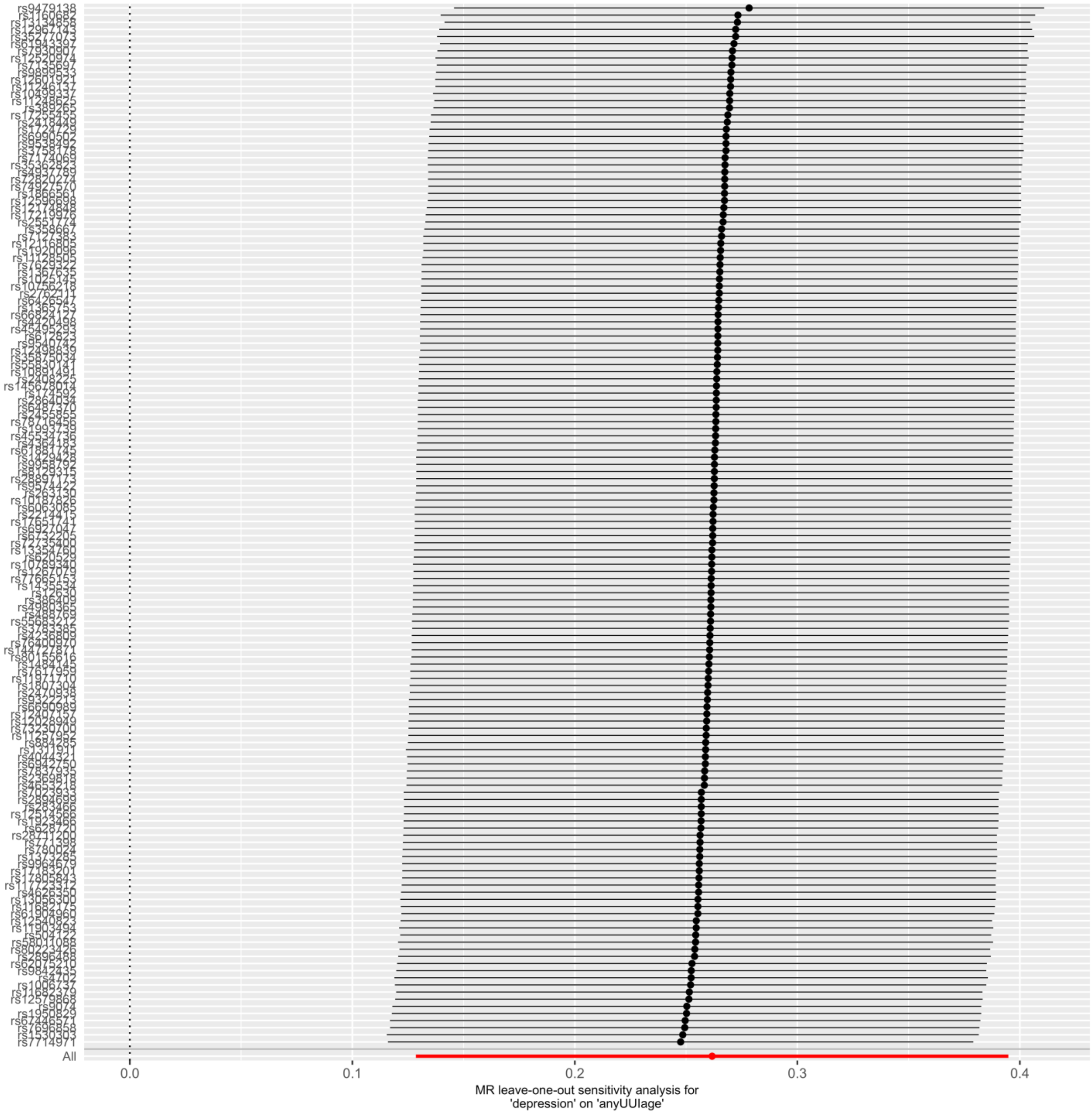
All
0.0
0.1
0.2
0.3
0.4
MR leave-one-out sensitivity analysis for
'depression' on 'anyUUIage'

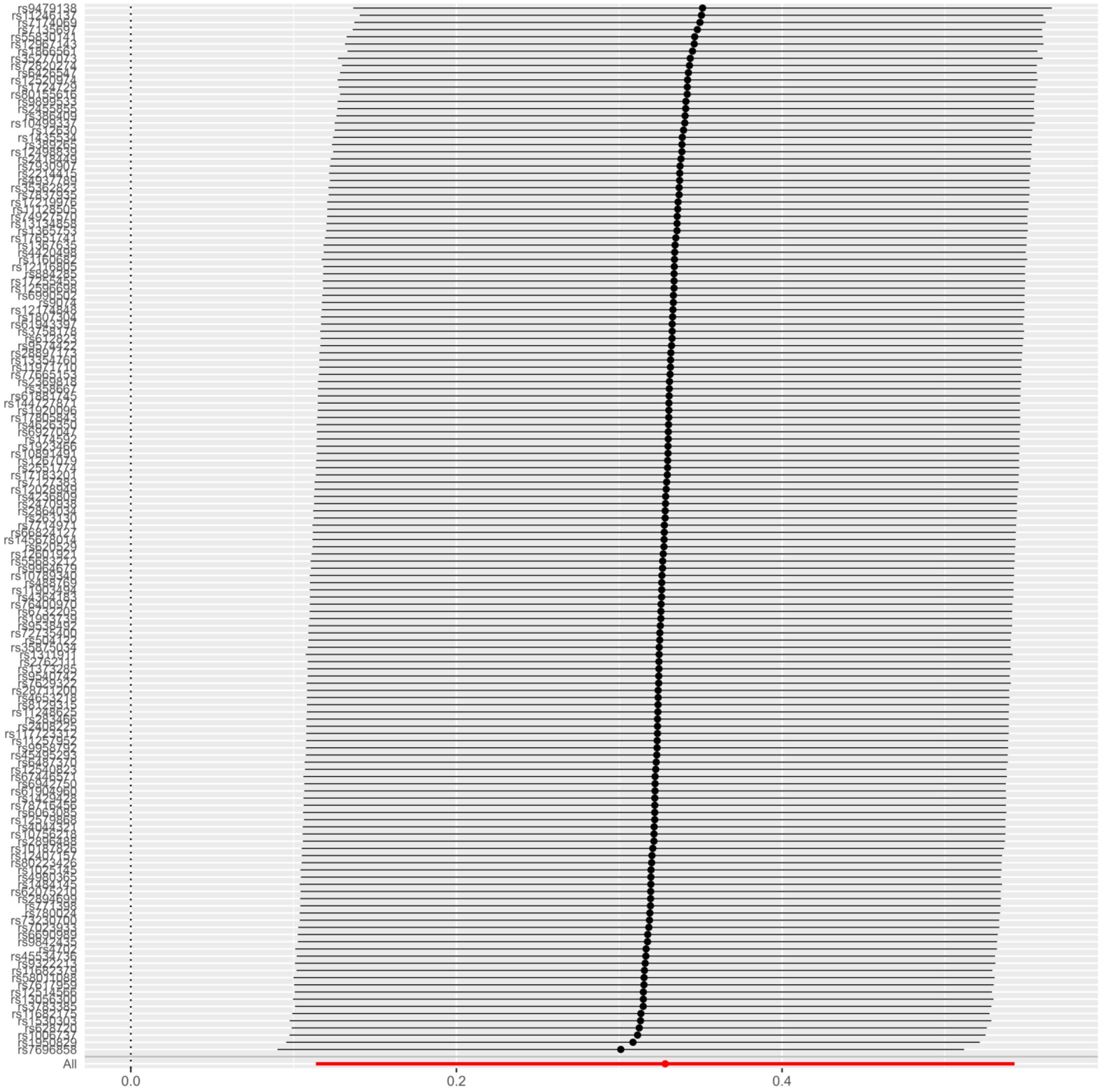

MR leave-one-out sensitivity analysis for
'depression' on 'anyMUIage'

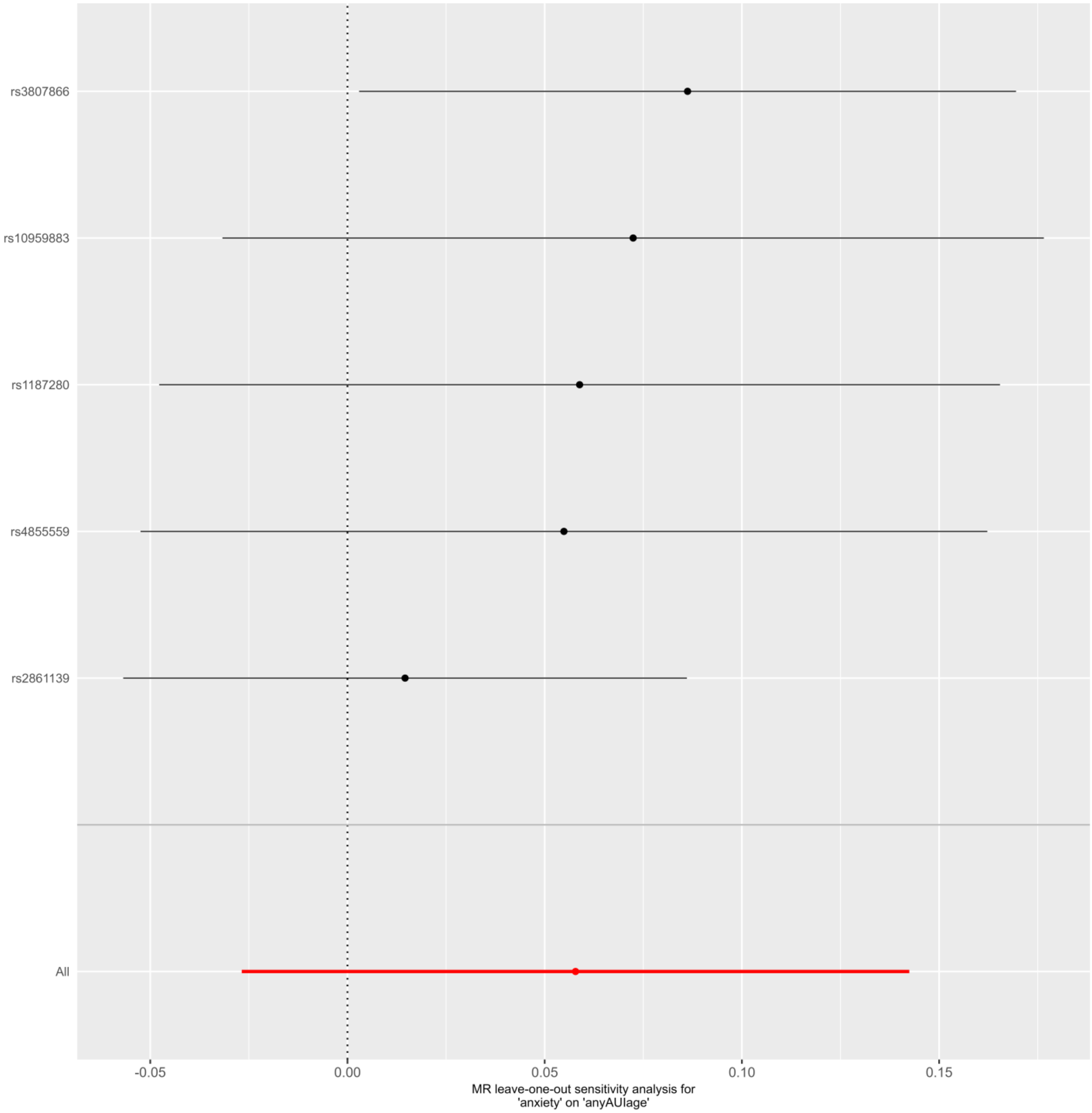
rs3807866
rs10959883
rs1187280
rs4855559
rs2861139
All
-0.05
0.00
0.05
0.10
0.15
MR leave-one-out sensitivity analysis for
'anxiety' on 'anyAUIage'

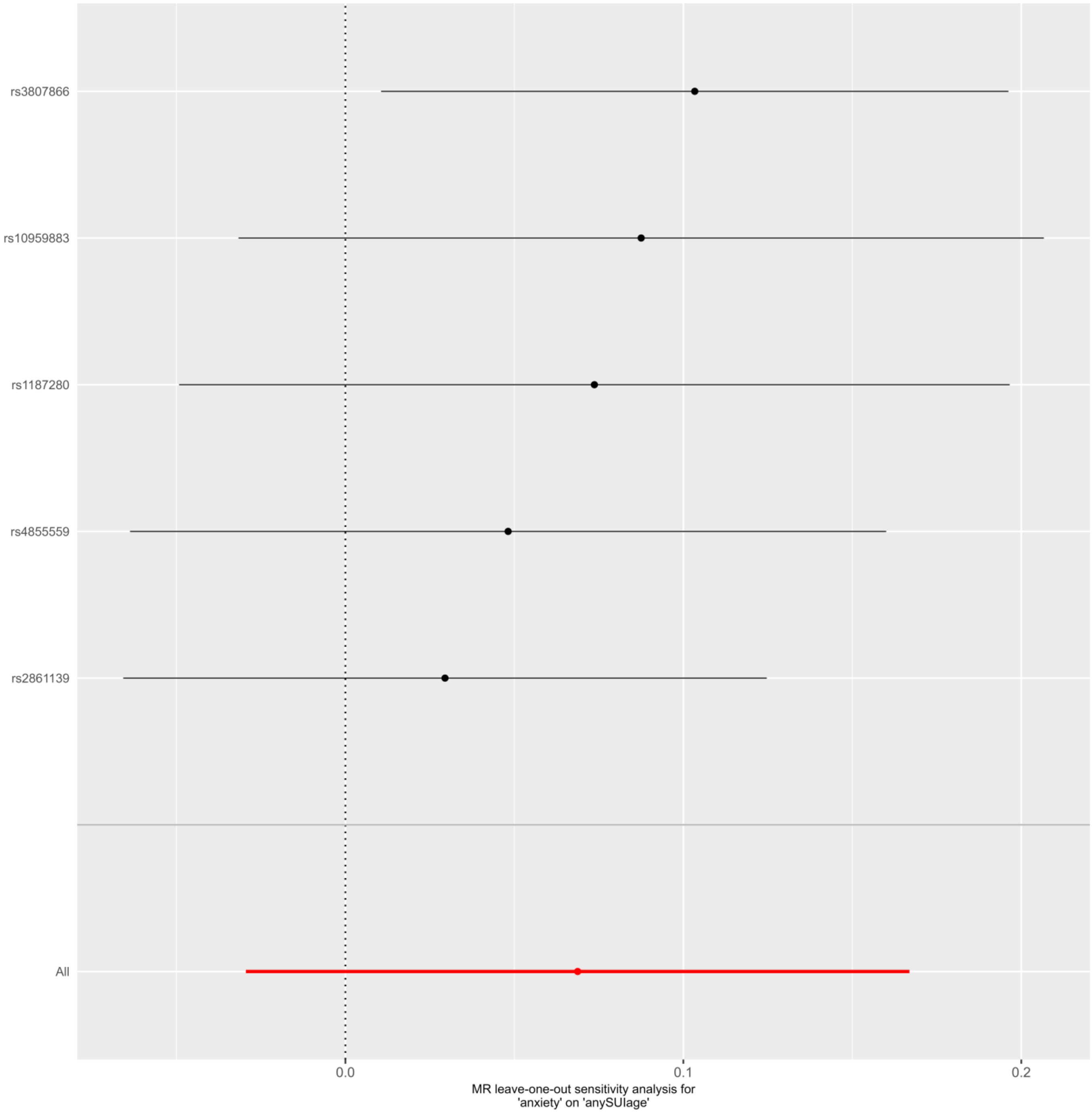
rs3807866
rs10959883
rs1187280
rs4855559
rs2861139
All
0.0
0.1
0.2
MR leave-one-out sensitivity analysis for
'anxiety' on 'anySUIage'

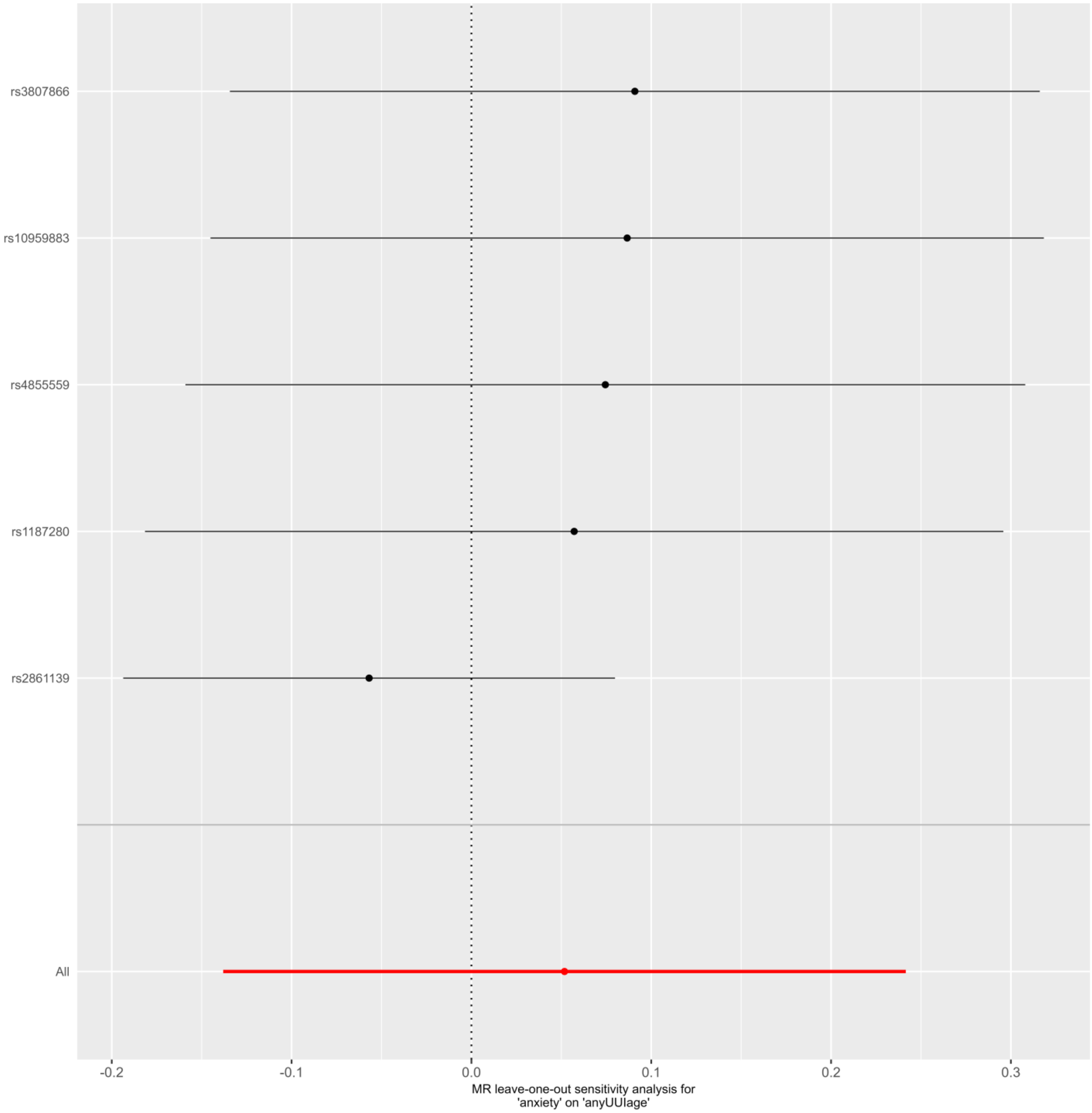
rs3807866
rs10959883
rs4855559
rs1187280
rs2861139
All
-0.2
-0.1
0.0
0.1
0.2
0.3
MR leave-one-out sensitivity analysis for
'anxiety' on 'anyUUIage'

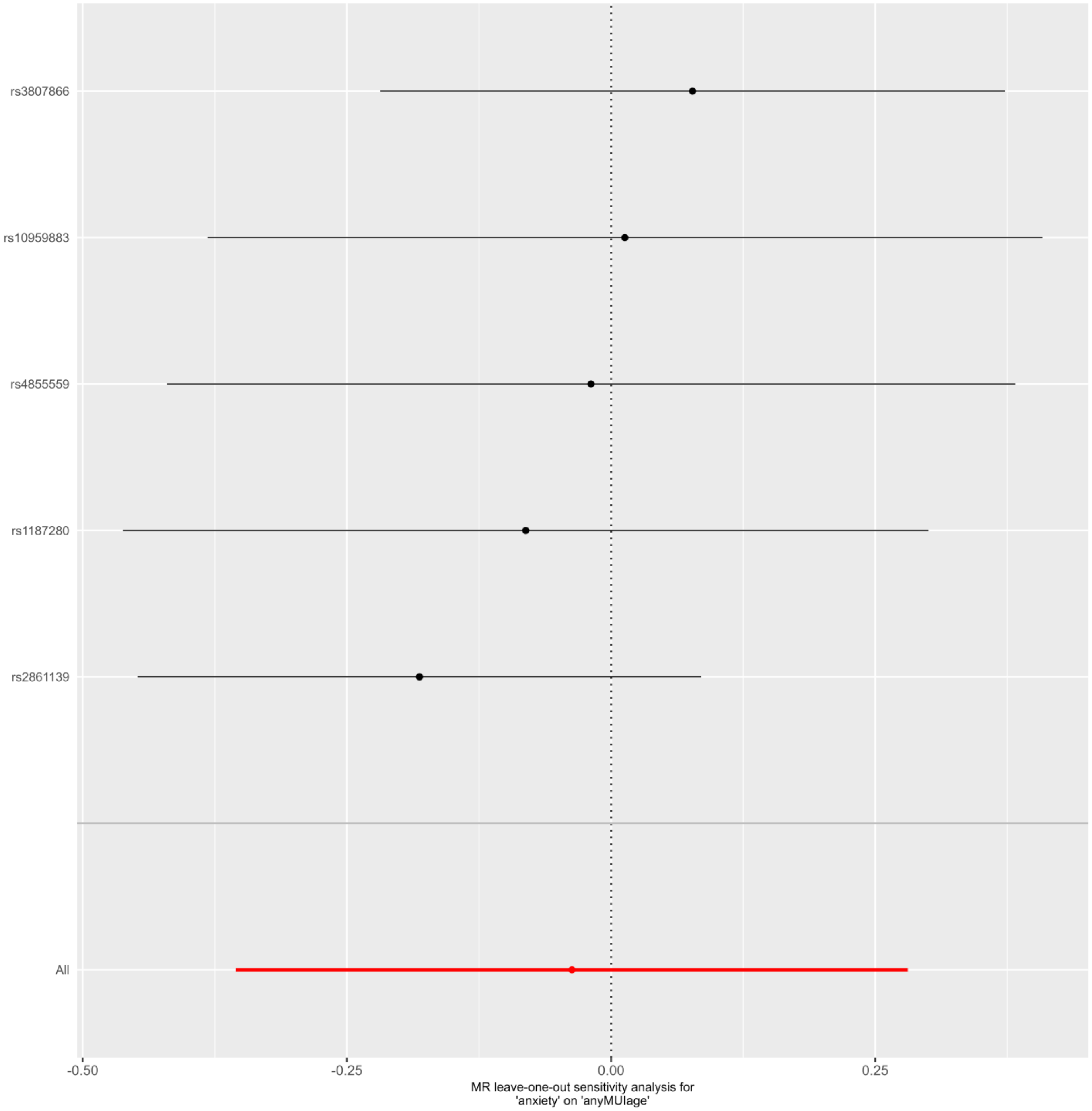
rs3807866
rs10959883
rs4855559
rs1187280
rs2861139
All
-0.50
-0.25
0.00
0.25
MR leave-one-out sensitivity analysis for
'anxiety' on 'anyMUIage'

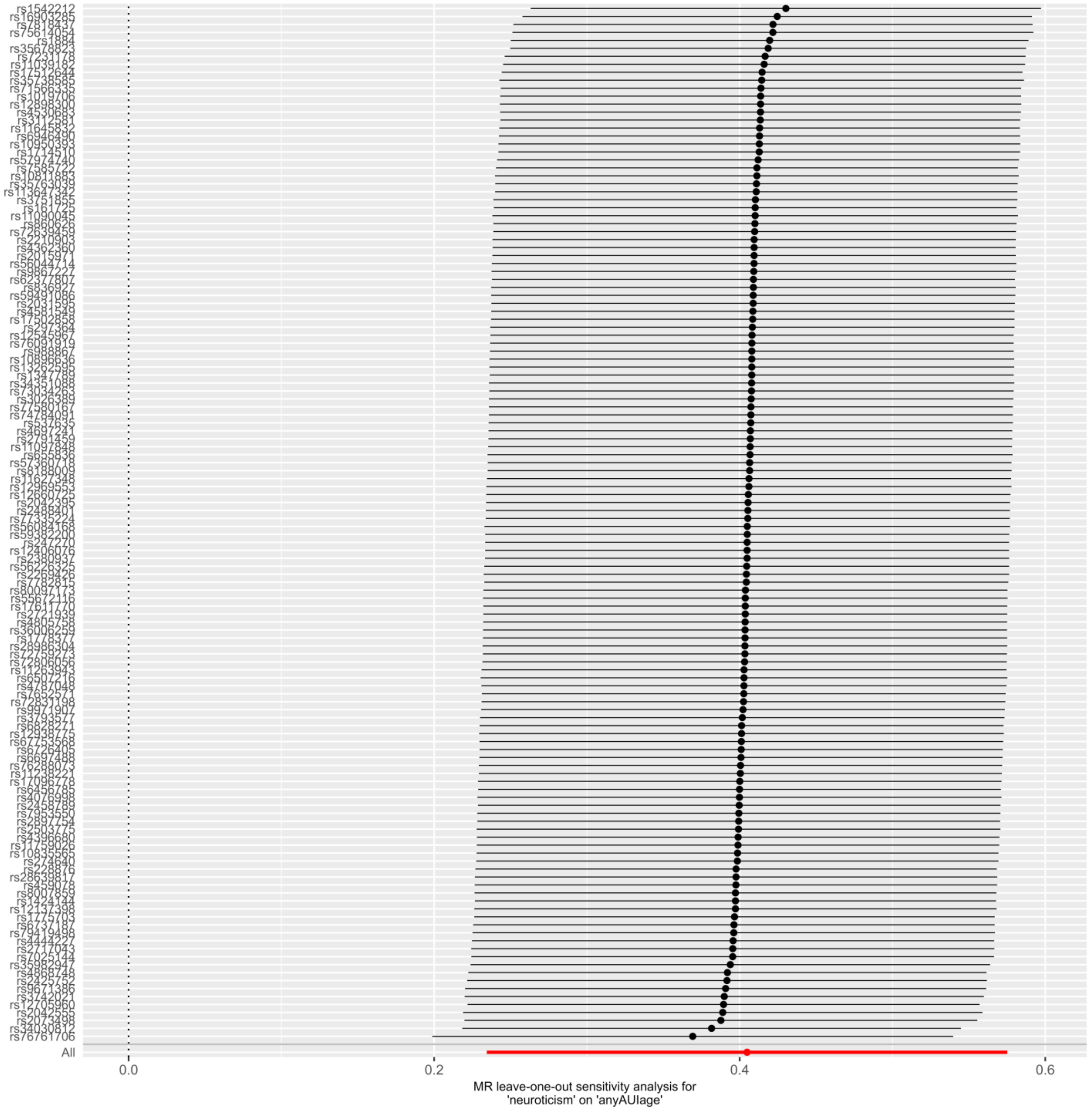
rs1542212
rs76761706
All
0.0
0.2
0.4
0.6
MR leave-one-out sensitivity analysis for
'neuroticism' on 'anyAUIage'

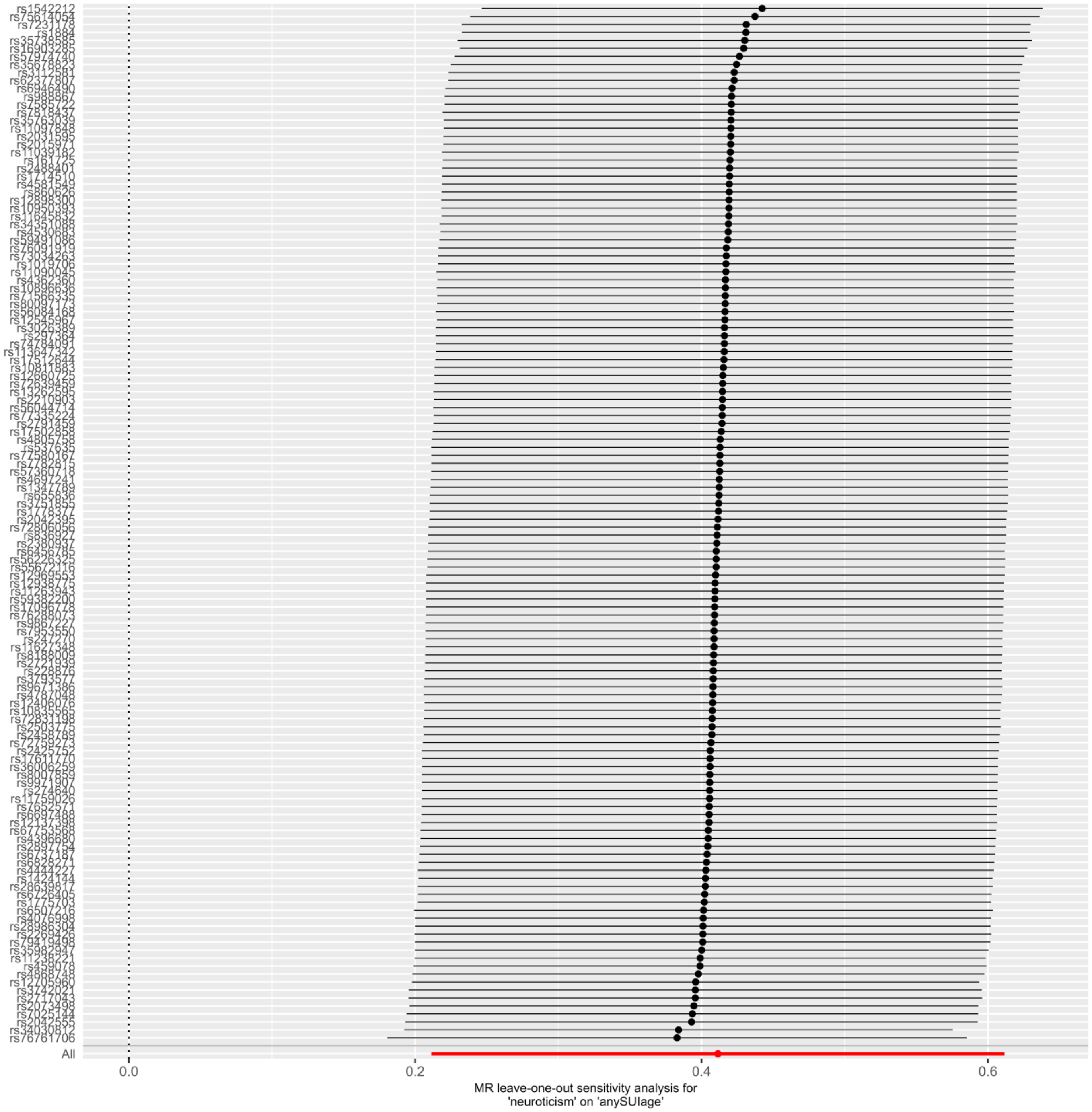

All
0.0
0.2
0.4
0.6
MR leave-one-out sensitivity analysis for
'neuroticism' on 'anySUIage'

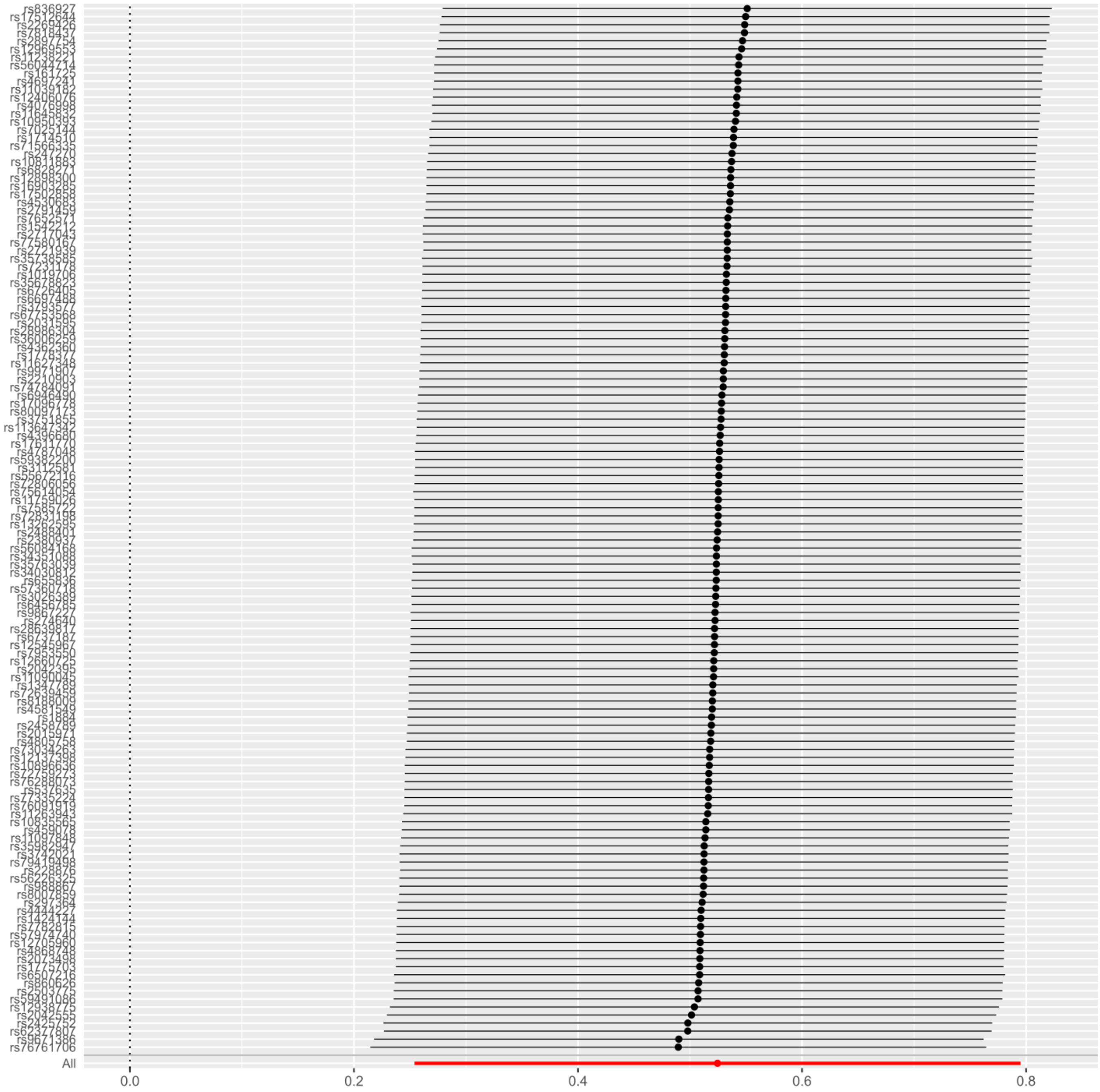

MR leave-one-out sensitivity analysis for
'neuroticism' on 'anyUUIage'

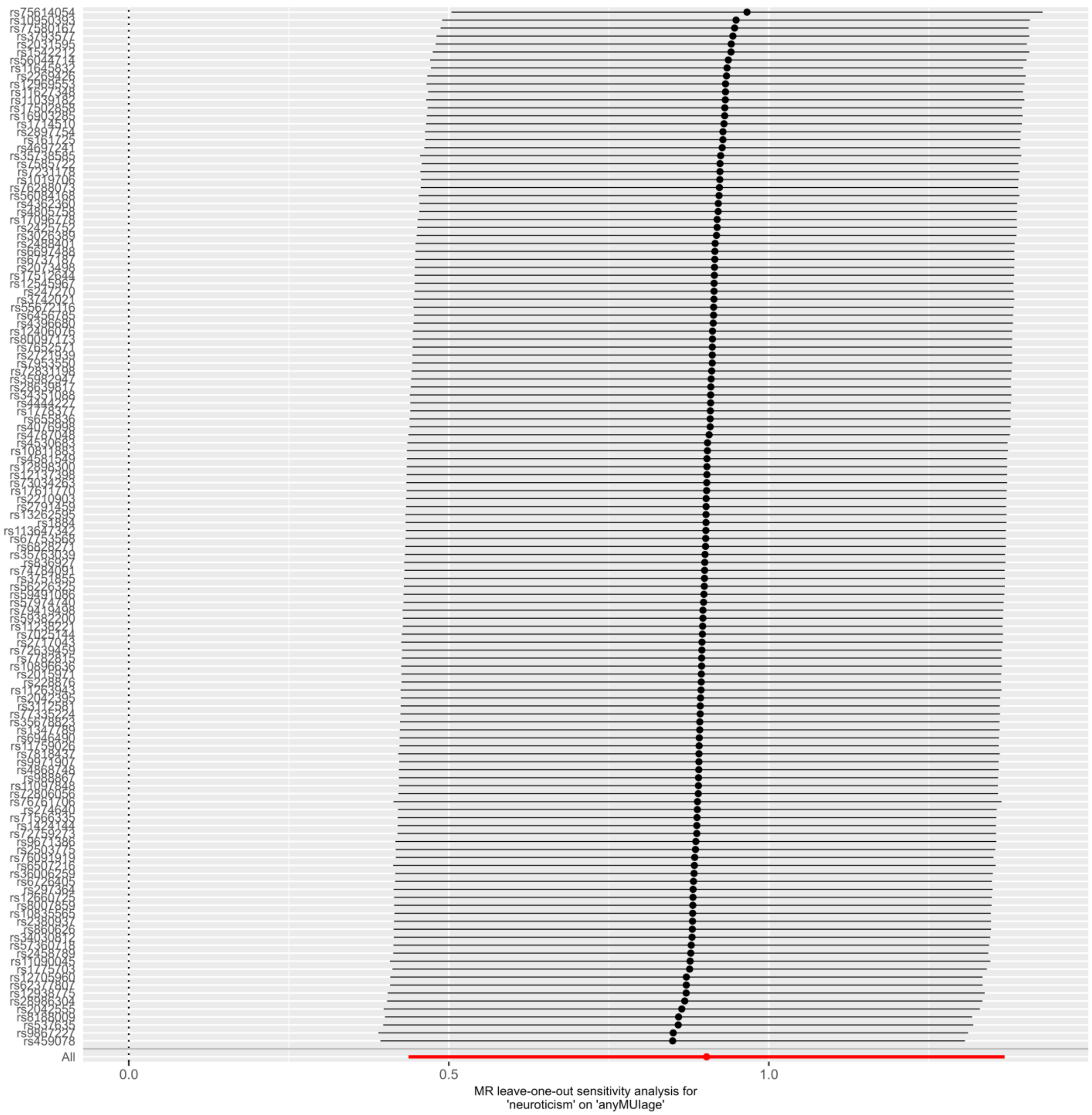

rs75614054
rs459078
All
0.0
0.5
1.0
MR leave-one-out sensitivity analysis for
'neuroticism' on 'anyMUIage'
